# A novel protein signature from plasma extracellular vesicles for non-invasive differential diagnosis of idiopathic pulmonary fibrosis

**DOI:** 10.1101/2021.05.07.21256811

**Authors:** Raju SR Adduri, Kai Cai, Karen V Alzate, Ravikiran Vasireddy, Jeffrey W Miller, Sergio Poli de Frías, Fernando Poli de Frías, Yasushi Horimasu, Hiroshi Iwamoto, Noboru Hattori, Yingze Zhang, Kevin F. Gibson, Anoop K Pal, Daniela Nicastro, Lily Li, Sujith Cherian, Lynette M Sholl, David A Schwartz, Daniel J Kass, Ivan O Rosas, Nagarjun V Konduru

**Author notes:** **Corresponding author:** Nagarjun V Konduru, Assistant Professor, Department of Cellular and Molecular Biology, University of Texas Health Science Center at Tyler, Tyler, TX75708, USA.

## Abstract

**Background:** Idiopathic pulmonary fibrosis (IPF) is a fibrosing interstitial pneumonia of unknown etiology often leading to respiratory failure. Over half of IPF patients present with discordant features of usual interstitial pneumonia on high-resolution computed tomography at diagnosis which warrants surgical lung biopsy to exclude the possibility of other interstitial lung diseases (ILDs). Therefore, there is a need for non-invasive biomarkers for expediting the differential diagnosis of IPF.

**Methods:** Using mass spectrometry, we performed proteomic analysis of plasma extracellular vesicles (EVs) in a cohort of subjects with IPF, chronic hypersensitivity pneumonitis, nonspecific interstitial pneumonitis, and healthy subjects (HS). A five-protein signature was identified by lasso regression and was validated in an independent cohort using ELISA. We evaluated the concordance between plasma EV proteome and the lung transcriptome data. Lastly, we compared the molecular pathways overrepresented in IPF by differentially expressed proteins and transcripts from EVs and lung tissues, respectively.

**Results:** The five-protein signature derived from mass spectrometry data showed area under the receiver operating characteristic curve of 0.915 (95%CI: 0.819-1.011) and 0.958 (95%CI: 0.882-1.034) for differentiating IPF from other ILDs and from HS, respectively. We also found that the EV protein expression profiles mirrored their corresponding mRNA expressions in IPF lungs. Further, we observed an overlap in the EV proteome- and lung mRNA-associated molecular pathways.

**Conclusions:** We discovered a plasma EV-based protein signature for differential diagnosis of IPF and validated this signature in an independent cohort. The signature needs to be tested in large prospective cohorts to establish its clinical utility.

## Introduction

Idiopathic pulmonary fibrosis (IPF) is a chronic, progressively fibrosing interstitial pneumonia of unknown etiology that often leads to respiratory failure^1^. With a median survival of 3.8 years, IPF appears to be more lethal than many cancer types^1^. Diagnosis at early stage and therapeutic intervention before the lung function is severely impaired could potentially improve treatment response and thereby prolong survival^2^.

A clear morphologic pattern of usual interstitial pneumonia (UIP) on high-resolution computed tomography (HRCT) with the absence of features that support alternate diagnosis confirms IPF, while discordant UIP features (detected in over 50% patients) ^3^ warrants histopathological analysis of surgical lung biopsies for definitive diagnosis^3^. However, surgical lung biopsy is not recommended for patients who are prone to high risk complications owing to their advanced age and poor fitness, and therefore the contemplation thwarts a confident diagnosis^4^. Additionally, UIP is not synonymous with IPF as other interstitial lung diseases (ILDs) including chronic hypersensitivity pneumonitis (CHP), and nonspecific interstitial pneumonia (NSIP), etc., may also exhibit a similar pattern^1^. Acquisition of adequate and faithfully representative biopsy sample is essential for accurate diagnosis of IPF and exclusion of other ILDs. Confirmation of IPF often requires a multidisciplinary approach involving pulmonologists, radiologists and pathologists with extensive expertise in the evaluation of patients with ILDs. Whilst the key features of IPF on HRCT or a multidisciplinary team consultation are important for accurate diagnosis of IPF, it needs to be acknowledged that access to these key elements of care in resource-constrained settings is a common challenge. Therefore, in some cases of IPF, a significant diagnostic delay is reported that is mainly attributable to the patients, general practitioners, and community hospitals^4^. Given that IPF diagnosed at early stage has better prognosis, a simple and quick diagnostic work up for IPF is expected to improve the outcome of the disease. Therefore, a novel non-invasive biomarker or a biomarker panel that can be tested in a routine laboratory is needed to expedite the differential diagnosis of IPF.

Several previous studies have reported biomarkers for differential diagnosis of IPF^5-9^. The majority of the studies evaluated selected genes/proteins with a putative role in the pathology of the ILDs for their efficacy as diagnostic markers, but their diagnostic efficacy has been unsatisfactory. As the molecular changes in IPF pathogenesis are complex, biomarker discovery using high throughput ‘omics’ approaches may yield robust biomarkers.

Extracellular vesicles (EVs), involved in intercellular signaling, contain different biomolecules (DNA, RNA, proteins, and metabolites) and their compositions reflect the molecular and physiological status of the parental cell. Due to their abundance in accessible body fluids^10^, in the past decade, EVs have gained prominence as a potential matrix for discovery of biomarkers of disease^10^. However, most of the studies examining lung diseases have focused on the miRNA cargo within EVs for identifying biomarkers, while proteins have been less explored^11^.

In the present study, we aimed to identify a protein signature in EVs that distinguishes IPF from other non-IPF ILDs and healthy subjects. Accordingly, we analyzed blood plasma EV protein profiles and performed a systematic biomarker discovery and validation.

## Materials and methods

### Patient cohorts and plasma collection

The study was conducted on a total of 118 samples including 44 IPF, 23 CHP, 19 NSIP and 32 healthy subjects (HS) from three different ILD centers. The samples were divided into two cohorts. Cohort-I consisted of 20 IPF, 11 CHP and 8 NSIP and 20 HS from University of Pittsburgh, Pittsburgh, USA and Brigham and Women’s hospital, Boston, USA, and have been extensively used for prior studies^12-17^. Cohort-II consisted of 24 IPF, 12 CHP, 11 NSIP and 12 HS from Hiroshima University, Hiroshima, Japan and Brigham and Women’s hospital, Boston, USA. The study was approved by institutional review boards of University of Pittsburgh (IRB# STUDY19040326 and STUDY20030223), Brigham and Women’s hospital (IRB#2012P000840), Hiroshima University (IRB#M326) and University of Texas Health Science Center at Tyler (IRB #20-019 & #0000370). Peripheral blood was collected from participants in EDTA or Citrated tubes after informed consent: and centrifuged at 1100xg for 10 minutes at room temperature.

### Preparation of extracellular vesicle samples

Size exclusion chromatography (SEC) was performed for isolating EVs using qEV original 35 nm pore size columns and automated fraction collector V1 setup (Izon Science US Ltd, MA, USA), from 150 µL of plasma, as per manufacturer’s instructions. Plasma was clarified by centrifugation at 2500xg for 15 minutes and subsequently at 10000xg for 10 min at 4°C. SEC columns were equilibrated with phosphate buffered saline and clarified plasma was loaded onto the column. After discarding 3 mL of void volume, 2 ml of EVs were collected and concentrated using 300 KDa centrifugal filters (Pall corp, NY, USA) at 3500xg at 4°C.

### Liquid chromatography–mass spectrometry/ mass spectrometry analysis

Samples were digested overnight with trypsin (Pierce) following reduction and alkylation with DTT and iodoacetamide (Sigma–Aldrich). The samples were then cleaned up using an Oasis HLB plate (Waters) and subsequently dried and reconstituted to approximately 0.5 mg/mL of 2% acetonitrile, 0.1% trifluoroacetic acid. 2 µL sample was injected onto a QExactive HF mass spectrometer coupled to an Ultimate 3000 RSLC-Nano liquid chromatography system. Samples were injected onto a 75 μm i.d., 15-cm long EasySpray column (Thermo) and eluted with a gradient from 0-28% buffer B over 90 min with a flow rate of 250 nL/min. Buffer-A contained 2% (v/v) acetonitrile and 0.1% formic acid, and buffer-B contained 80% (v/v) acetonitrile, 10% (v/v) trifluoroethanol, and 0.1% formic acid in water. The mass spectrometer operated in positive ion mode with a source voltage of 2.2 kV and an ion transfer tube temperature of 275°C. Mass spectrometry (MS) scans were acquired at 120,000 resolution in the Orbitrap and up to 20 MS/MS spectra were obtained for each full spectrum acquired using higher-energy collisional dissociation for ions with charges 2-8. Dynamic exclusion was set for 20s after an ion was selected for fragmentation.

Raw MS data files were analyzed using Proteome Discoverer v2.4 (Thermo), with peptide identification performed using Sequest HT searching against the human protein database from UniProt. Fragment and precursor tolerances of 10 ppm and 0.02 Da were specified, and three missed cleavages were allowed. Carbamidomethylation of cysteine was set as a fixed modification, with oxidation of methionine set as a variable modification. The false-discovery rate (FDR) cutoff was 1% for all peptides.

### Supplementary methods

Description of methods related to molecular analysis and statistics are detailed in supplementary document 1.

## Results

### Clinical features of IPF, non-IPF and healthy subjects

Clinical characteristics of participants diagnosed with IPF, non-IPF (CHP and NSIP), and HS from cohorts I and II are summarized in table 1 and 2, respectively. The majority of participants in cohorts I and II are non-Hispanic whites (USA) and East Asians (Japan), respectively, and are smokers over 50 years of age. Diagnosis of IPF or non-IPF ILDs was made according to consensus criteria^18^.

**Table 1.** Demographic information and lung function parameters of subjects in cohort-I.

|  |  | IPF | Non-IPF |  | HS |
| --- | --- | --- | --- | --- | --- |
|  |  |  | CHP | NSIP |  |
| **Total (N)** |  | 20 | 11 | 8 | 20 |
| **Demographic information** |  |  |  |  |  |
| Age (Years) |  | 68.5 (55-79) | 57.9 (32-70) | 53.3 (27-77) | 63.3 (47-80) |
| Gender (N) | Male | 16 (80%) | 6 (55%) | 2 (50%) | 12 (60%) |
|  | Female | 4 (20%) | 5 (45%) | 2 (50%) | 8 (40%) |
| Smoking (N) | Ever | 9 | 3 | 0 | 13 (65%) |
|  | Never | 6 | 3 | 1 | 7 (35) |
|  | NA | 5 | 5 | 3 | 0 (0%) |
| **Lung function parameters** |  |  |  |  |  |
| FEV1% |  | 87.0 (54.7-129.0)* | 79.7 (45.9-108.3)# | 63.7 (34.5-78.2) | - |
| FVC% |  | 73.0 (49.2-99.6)* | 70.8 (37.4-110.7)# | 57.9 (30.9-85.1) | - |
| FEV1/FVC |  | 81.6 (73.4-89.3)* | 82.9 (71.3-97.6)# | 82.6 (69.4-91.2) | - |

**Table 2.** Demographic information and lung function parameters of subjects in cohort -II.

|  |  | IPF | Non-IPF |  | HS |
| --- | --- | --- | --- | --- | --- |
|  |  |  | CHP | NSIP |  |
| **Total (N)** |  | 24 | 12 | 11 | 12 |
| **Demographic information** |  |  |  |  |  |
| Age (Years) |  | 66.1 (53-79) | 66.4 (54-81) | 70.5 (54-82) | 62.6 (42-75) |
| Gender (N) | Male | 18 (75%) | 8 (66.7%) | 5 (45.5%) | 7 (58.3%) |
|  | Female | 6 (25%) | 4 (33.3%) | 6 (54.5%) | 5 (41.7%) |
| Smoking (N) | Ever | 10 (41.7%) | 4 (33.3%) | 3 (27.3%) | 2 (16.7%) |
|  | Never | 5 (20.8%) | 4 (33.3%) | 4 (36.4%) | 3 (25%) |
|  | NA | 9 (37.5%) | 4 (33.3%) | 4 (36.4%) | 7 (58.3%) |
| **Lung function parameters** |  |  |  |  |  |
| FEV1% |  | 71.7 (36.5-104.2) | 76 (37-117.8) | 86.67 (56.9-116.6) | - |
| FVC% |  | 65.5 (32.5-99.2) | 71 (37-123.2) | 79.58 (52.9-100.3) | - |
| FEV1/FVC |  | 96.8 (77.8-120) | 94.34 (79.8-116.3) | 92.21 (69.4-120.7) | - |

### Characterization and proteomic landscape of plasma extracellular vesicles

Extracellular vesicles were isolated from blood plasma in cohort-I consisting of 20 IPF, 19 non-IPF (11 CHP and 8 NSIP) and 20 HS. Cryo-electron microscopy imaging of plasma EV preparations revealed the presence of intact spherical vesicles consisting a lipid bi-layer and were on average 100 nm in diameter (Figure 1A). Nanoparticle tracking analysis of EV preparations further confirmed that the majority EVs were in the range of 50-200 nm in diameter (Figure 1B). In addition, the tetraspanin protein CD9, and other exosomal proteins such as HSP70 and ALIX were detected while Calnexin, an endoplasmic reticulum membrane protein, was absent (Figure 1C). Altogether, these results demonstrate the qualitative characteristics of EVs and suggest that the plasma EV preparations were enriched with exosomes.

**Figure 1.**
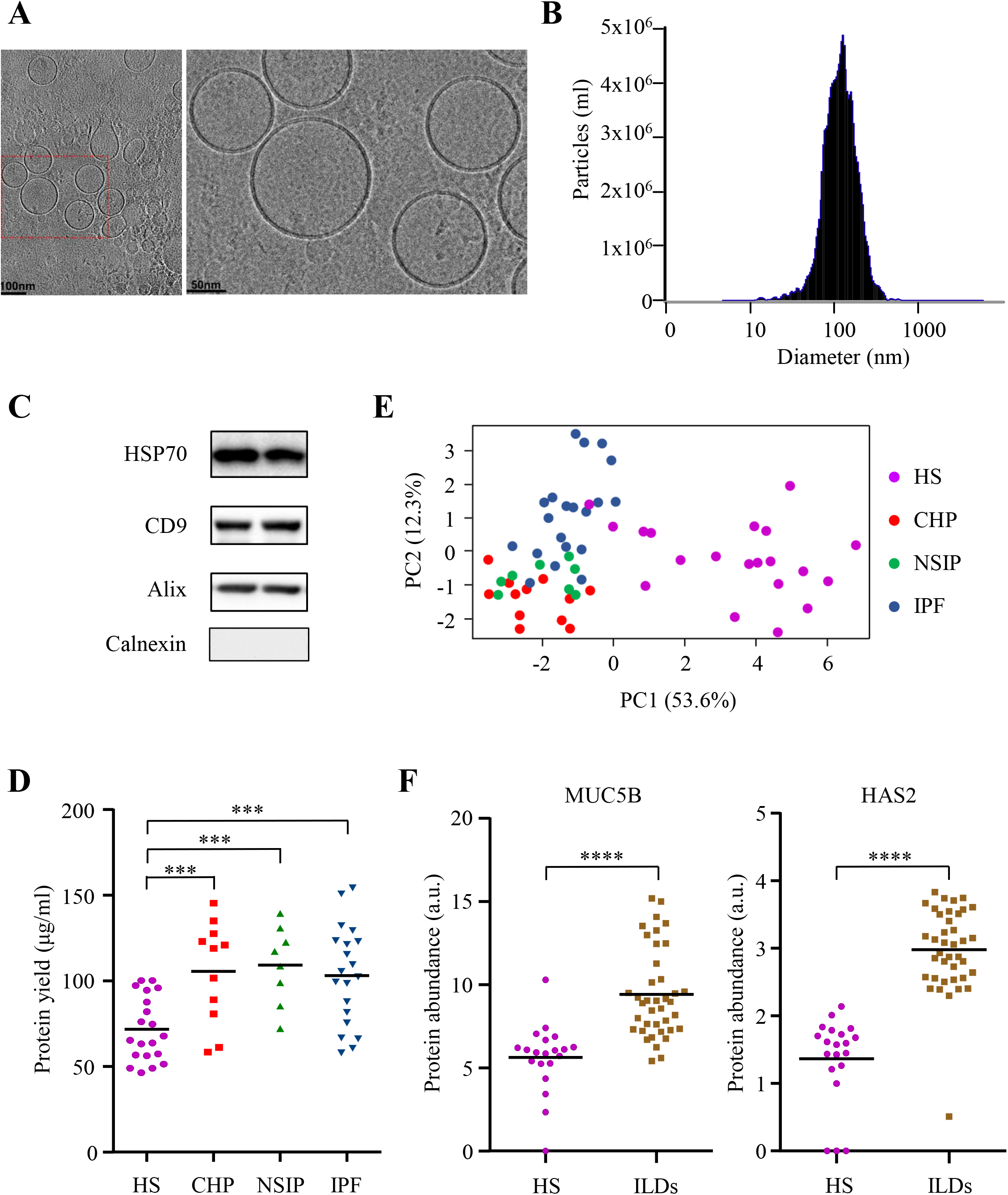
EV characterization, protein measurements and principle component analysis of EV proteomes. **A)** Representative cryo-electron micrograph of EVs isolated from peripheral blood plasma of patients in the study. **B)** Size and quantification of plasma EVs measured by Nanoparticle Tracking Analysis against 0.1 μm fluorescent polystyrene beads standard. **C)** Detection of common EV markers using Western blot for two representative plasma samples. Presence of EV membrane marker (CD9), internal markers (HSP70 and Alix), and absence of endoplasmic reticulum marker (Calnexin) are shown. **D)** Protein yield from plasma EV lysates of healthy subjects (HS) and ILD patients measured using Bicinchoninic Acid assay. Horizontal bar represents median expression value in the group. The asterisk *** denotes p<0.001. **E)** Principle component analysis of proteomic profiles of HS, non-IPF (CHP and NSIP) and IPF. Two principle components explaining highest variation were plotted on X and Y axis respectively. **F)** Expression levels of MUC5B and HAS2 in HS and ILDs determined by mass spectrometry. Horizontal bar represents mean expression value in the group. The asterisk **** denotes p<0.0001.

The total protein within the EVs in samples varied from 48.9 µg to 154.2 µg per ml of plasma. Interestingly, the EV protein yield was higher among patients with ILDs than in controls (p<0.001) (Figure 1D) suggesting that a diseased state could potentially increase EV secretion with altered protein composition. However, the protein yields from plasma EVs of IPF and non-IPF were not significantly different (Figure 1D). Mass spectrometry based proteomic profiling of EVs from 20 IPF, 19 non-IPF (11 CHP and 8 NSIP) and 20 HS quantified the expression of 520 proteins. Principle component analysis of the 250 proteins with highest variance revealed that the protein profiles of EVs from healthy subjects, CHP, NSIP and IPF clustered according to the lung conditions (Figure 1E). HS samples appeared distinct from all the ILDs. While the three ILDs clustered separately, they exhibited major overlap, underscoring the similarities in the proteome profiles. We next compared proteomic profiles of all ILDs (CHP, NSIP and IPF) together with HS and identified 196 and 46 up- and downregulated proteins in ILDs, respectively (Figure S1). As expected, several upregulated proteins enriched in EVs have also been reported to be overexpressed in fibrotic lungs. For example, Mucin 5B (MUC5B), a mucin often found over expressed in pulmonary fibrotic foci^19^ and Hyaluronan synthase 2 (HAS2) that promotes invasive phenotype in fibroblasts from lung fibrosis^20^ were overexpressed in EVs from IPF as well as non-IPF compared to healthy subjects (Figure 1F).

### EV protein biomarkers for non-invasive differential diagnosis of IPF

A schematic of biomarker discovery and validation pipeline is shown in Figure 2A. To identify biomarkers that distinguish IPF from non-IPF, we performed differential expression analysis on the proteome profiles of EVs isolated from plasma of 20 IPF and a total of 19 non-IPF patients. Non-IPF patients included 11 CHP and 8 NSIP samples exhibiting fibrosis that served as ILD controls to identify proteins specific to IPF. A total of 30 differentially expressed genes were identified at FDR and |log2 fold change| cut offs of 0.05 and 0.585, respectively (Table S1). Based on the premise that upregulated proteins as biomarkers are more suitable for diagnostic use in clinical settings and with the aim to develop an easy-to-adopt assay, we prioritized upregulated proteins for further analysis. We applied Least Absolute Shrinkage and Selection Operator (LASSO) on mass-spectrometry protein abundances of upregulated proteins, coupled with 5 -fold cross-validation to minimize prediction error to identify robust biomarker panel. At a minimum value of λ (0.000837), a five-protein signature comprising High mobility group box protein 1 (HMGB1), surfactant protein B (SFTPB), Aldolase A (ALDOA), calmodulin like 5 (CALML5) and Talin-1 (TLN1) discriminated IPF from other ILDs with minimum cross validation error (Figure 2B).

**Figure 2.**
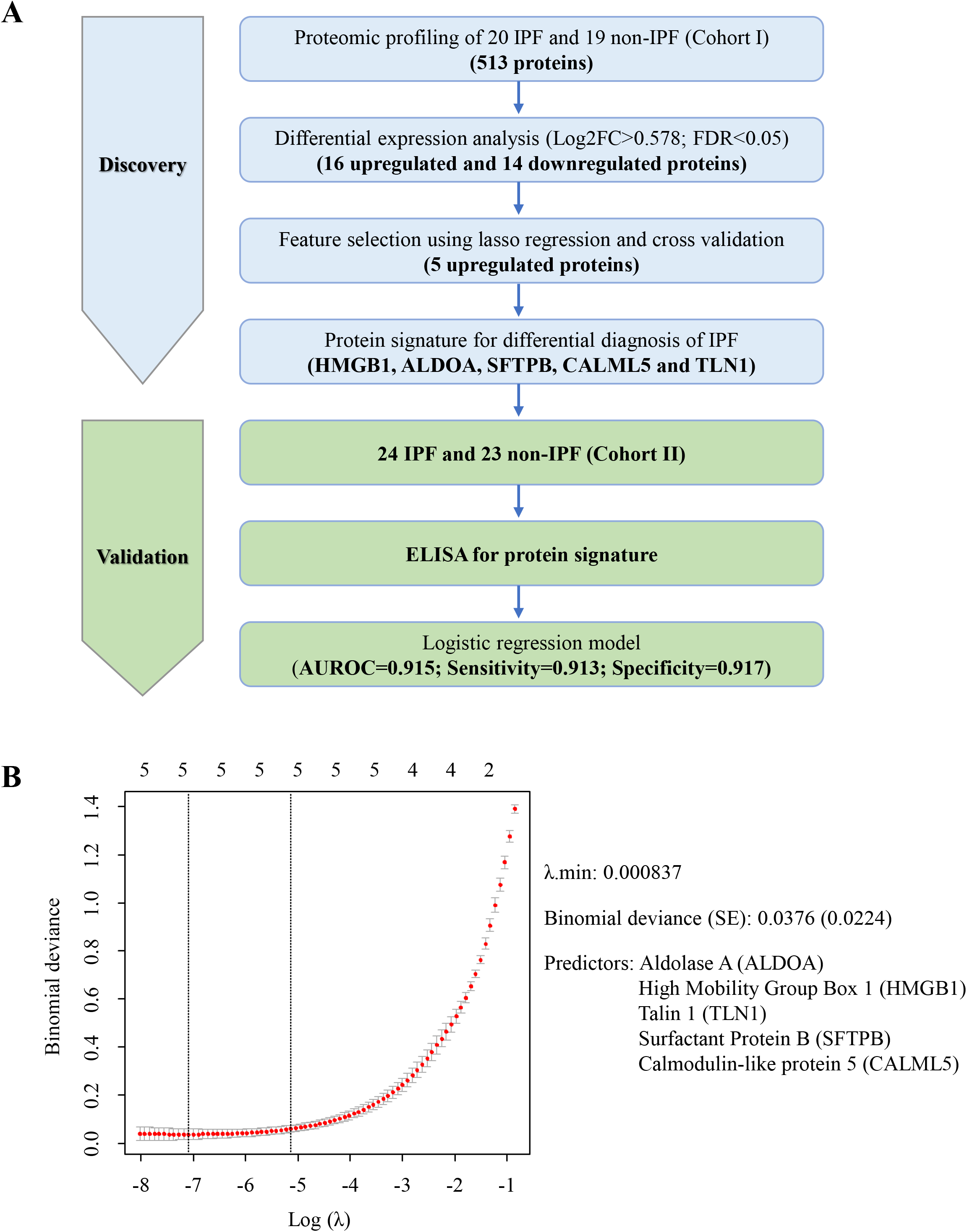
Discovery of EV protein biomarker signature for differential diagnosis of IPF from other ILDs. **A)** Study design of systematic discovery of EV protein biomarker signature is depicted in a flow chart. **B)** Plot showing cross validation error at different values of tuning parameter (λ) in lasso regression in cohort-I. Red dotted curve shows the 5-fold cross-validation curve while the whiskers represent upper and lower standard deviations, respectively. The vertical dotted lines show the locations of λ.min and λ.1se. The numbers across the top show the number of nonzero coefficient estimates. SE, Standard error.

Encouraged by our findings in the discovery phase, we estimated the levels of our protein biomarkers in plasma EVs of 24 IPF, 23 non-IPF and 12 healthy subjects from an independent validation cohort (cohort II) using ELISA. The expression levels of HMGB1, ALDOA and CALML5 were different between IPF and non-IPF in an independent cohort (Figure 3A). To evaluate the efficacy of the five EV proteins for differential diagnosis of IPF, we performed logistic regression and constructed ROC curves (Figure 3B). Our protein signature classified IPF and non-IPF with excellent efficacy in cohort II (AUROC=0.915, 95% CI: 0.819-1.011; Figure 3B). To check whether the number of proteins in the classifier could be minimized, we applied backward stepwise elimination approach and selected a sparse model comprising CALML5, HMGB1 and TLN1 which exhibited an AUROC of 0.902 (95% CI:0.805-1) (Figure 3B). Taken together, these results suggest that the five-protein EV biomarker panel has superior efficacy in differential diagnosis of IPF. Further, five out of four proteins, namely CALML5, HMGB1, CALML5 and SFTPB exhibited significantly high expression in IPF compared to HS (Figure S2A). The five-protein biomarker signature could distinguish IPF from healthy controls with similar efficiency (AUROC=0.958) (Figure S2B). Backward stepwise elimination approach yielded a model comprising CALML5 and HMGB1which exhibited AUROC of 0.951 (95% CI:0.876-1.026) (Figure S2B). Taken together, these results suggest that the EV biomarkers have excellent efficacy in discriminating IPF from HS.

**Figure 3.**
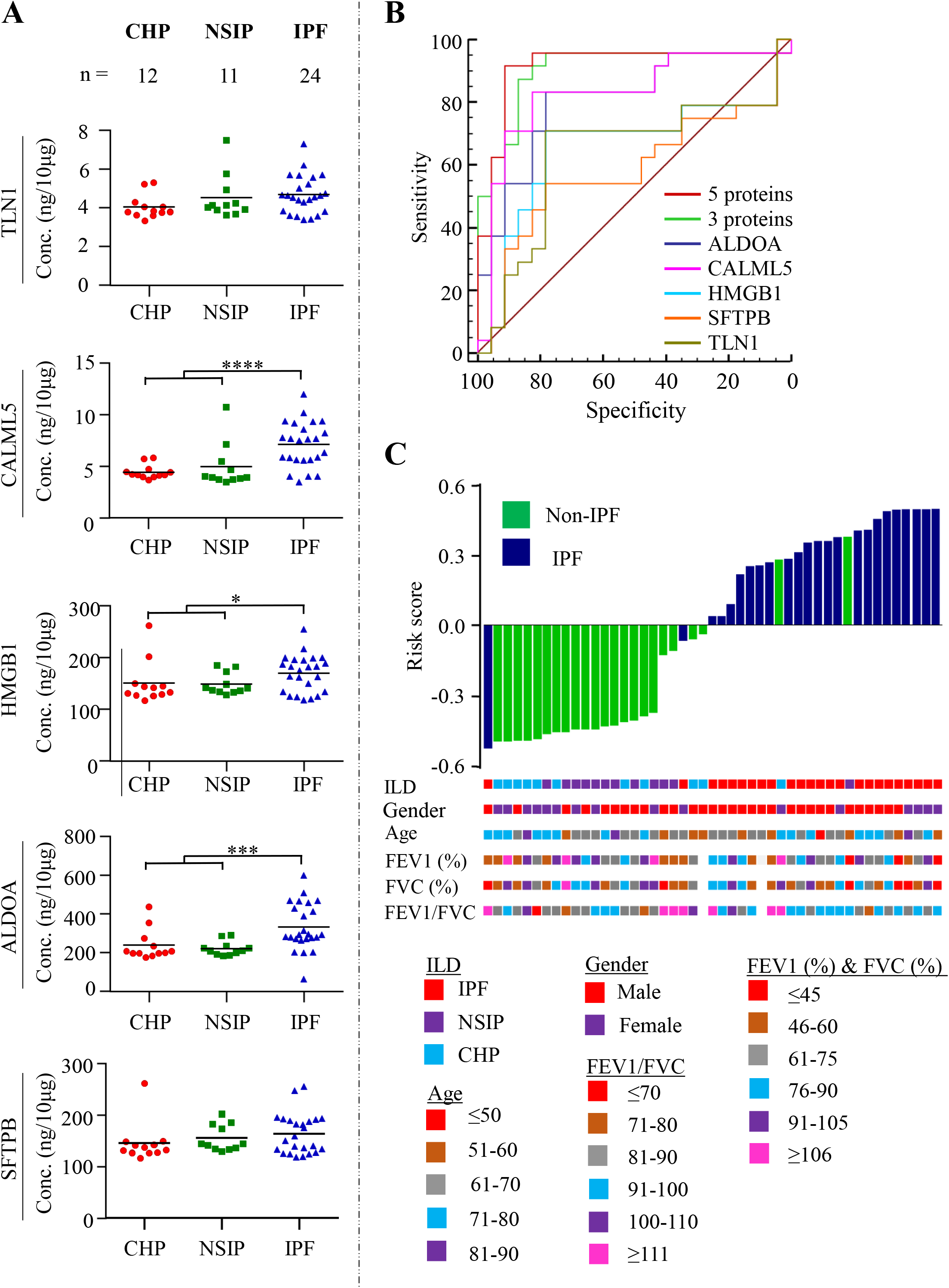
Quantification of five-protein signature in an independent cohort of IPF and non-IPF patients. **A)** Expression of five-protein signature in plasma EVs of 24 IPF and 23 non-IPF was performed using sandwich ELISA with purified recombinant human protein as standard. The asterisks *, ** * and **** denote p<0.05, p<0.001 and p<0.0001, respectively. **B)** Area under receiver operating characteristic (AUROC) curves for logistic regression model generated for five proteins on 24 IPF and 23 non-IPF patients. **C)** Risk scores calculated from predictive probabilities from logistic regression model for each patient and are shown in the water fall plot in ascending order.

### EV proteome identifies deregulated signaling pathways in IPF

Comprehensive analysis of transcriptome profiles has been explored previously to identify potential pathways involved in the pathogenesis of IPF^21,22^. We determined whether the EV protein levels followed the expression blueprint of their corresponding transcripts in the IPF lung tissue. To this end, we first performed differential expression analysis for IPF and healthy subjects. The EV proteome of IPF patients revealed 154 upregulated and 32 downregulated proteins that were identified at an FDR<0.05 and |log_2_ fold change| >0.585 (Figure 4A, Table S2). Four (HMGB1, SFTPB, ALDOA and CALML5) of five proteins were also found to be upregulated in IPF in comparison to HS. Further, we performed differential expression analysis for transcriptomes of the lung tissues from 103 IPF and 103 HS^21^. We identified 1299 upregulated genes and 2359 downregulated genes from IPF lung tissue (Figure 4A, Table S3). Notably, of the 186 proteins identified to be differentially expressed in EVs, the expression profiles of 36 differentially expressed proteins were found to be concordant with their corresponding mRNA expression profiles in the lung tissue (Table S4). We next elucidated whether the EV protein profiles could also identify the potential pathways involved in the pathogenesis of IPF. The differentially expressed proteins and genes were subjected to overrepresentation analysis independently using the gProfiler package^23^. Extracellular matrix and collagen deposition were commonly identified by EV proteome and lung transcriptome. Interestingly, EV proteome identified key IPF molecular events such as neutrophil and platelet degranulation, and surfactant protein dysfunction while transcriptome did not (Figure 4B, Table S5A). However, lung transcriptome identified several other pathways including G-protein couple receptor signaling, chemokine signaling, and mucin dysregulation (Figure 4B, Tables S5B).

**Figure 4.**
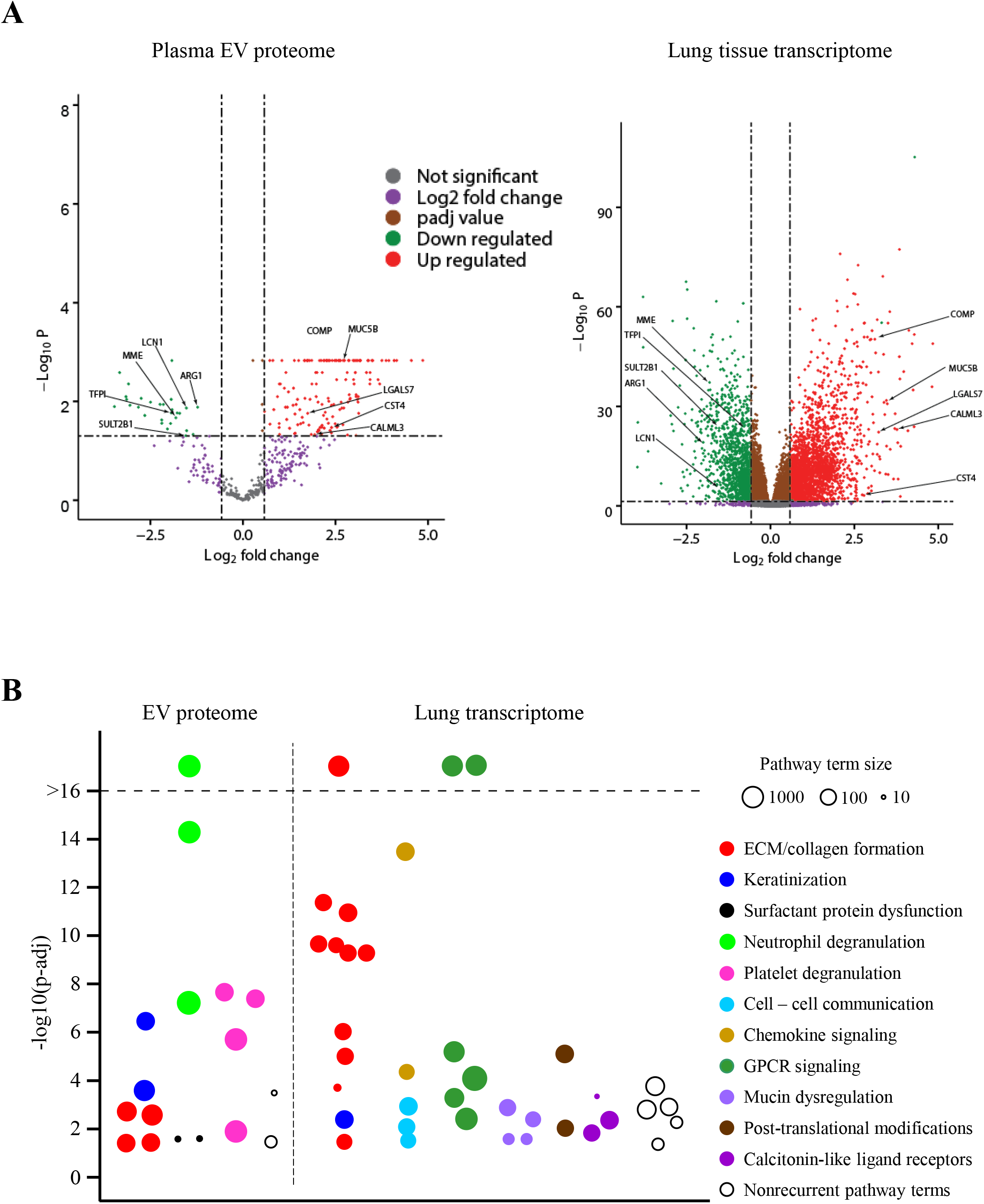
Comparison of EV proteome and lung tissue transcriptome in IPF. **A)** Differentially expressed proteins and transcripts in IPF were identified at FDR<0.05 and |log2 fold change| >0.585 when compared with healthy subjects. Top five upregulated and five downregulated genes/proteins common between proteome and transcriptome are labeled. **B)** Manhattan plot showing Reactome pathway terms associated with plasma EV proteome and lung tissue transcriptome in IPF. Pathways showing similar intersection genes/proteins are grouped and color coded to aid interpretation. Size of the circle indicates the number of gene entries belonging to the pathway in Reactome database. Y-axis represents -log10 value of adjusted p-value and was capped at 16. ECM: Extracellular matrix, GPCR: G-protein coupled receptor.

We also sought to understand whether the EV proteomic profiles could identify different biological functions pertaining to different ILDs. Ingenuity pathway analysis of differentially expressed genes in IPF and CHP revealed the enrichment of distinct biological functions in the two ILDs. The EV proteome in IPF exhibited enrichment of functions such as fibrogenesis, interaction of fibroblasts, binding of connective tissue cells, differentiation of epithelial tissue, and formation of focal adhesions (Table 4A). Contrastingly, EVs in CHP exhibited enrichment of functions involving angiogenesis, complement activation, endothelial cell development, biosynthesis of amide and eicosanoids as well as opsonization of cells (Table 4B).

**Table 3.**
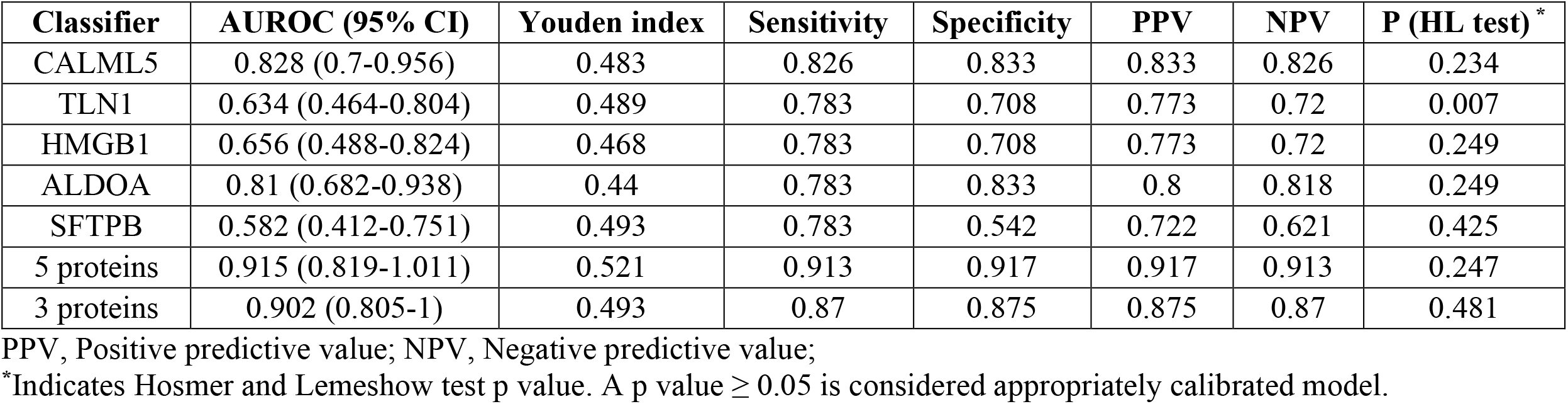
Summary of the logistic regression model generated for different classifiers for differential diagnosis of IPF.

| **Classifier** | **AUROC (95% CI)** | **Youden index** | **Sensitivity** | **Specificity** | **PPV** | **NPV** | **P (HL test) \*** |
| --- | --- | --- | --- | --- | --- | --- | --- |
| CALML5 | 0.828 (0.7-0.956) | 0.483 | 0.826 | 0.833 | 0.833 | 0.826 | 0.234 |
| TLN1 | 0.634 (0.464-0.804) | 0.489 | 0.783 | 0.708 | 0.773 | 0.72 | 0.007 |
| HMGB1 | 0.656 (0.488-0.824) | 0.468 | 0.783 | 0.708 | 0.773 | 0.72 | 0.249 |
| ALDOA | 0.81 (0.682-0.938) | 0.44 | 0.783 | 0.833 | 0.8 | 0.818 | 0.249 |
| SFTPB | 0.582 (0.412-0.751) | 0.493 | 0.783 | 0.542 | 0.722 | 0.621 | 0.425 |
| 5 proteins | 0.915 (0.819-1.011) | 0.521 | 0.913 | 0.917 | 0.917 | 0.913 | 0.247 |
| 3 proteins | 0.902 (0.805-1) | 0.493 | 0.87 | 0.875 | 0.875 | 0.87 | 0.481 |

PPV, Positive predictive value; NPV, Negative predictive value;
\*Indicates Hosmer and Lemeshow test p value. A p value $\geq 0.05$ is considered appropriately calibrated model.

**Table 4.** Diseases and function terms enriched in IPF (A) and CHP (B) based on plasma EV proteome analysis.

| **Diseases or Functions Annotation** | **p value** |
| --- | --- |
| Adhesion of immune cells | 6.77E-24 |
| Activation of myeloid cells | 3.42E-12 |
| Fibrogenesis | 1.69E-11 |
| Formation of filaments | 9.89E-11 |
| Adhesion of epithelial cells | 9.29E-10 |
| Activation of antigen presenting cells | 1.51E-08 |
| Systemic autoimmune syndrome | 3.63E-08 |
| Binding of connective tissue cells | 3.99E-08 |
| Differentiation of epithelial tissue | 1.12E-07 |
| Formation of focal adhesions | 1.16E-07 |
| Accumulation of myeloid cells | 1.28E-07 |
| Damage of endothelial tissue | 5.76E-07 |
| Adhesion of connective tissue cells | 5.80E-07 |

B)
| **Diseases or Functions Annotation** | **p value** |
| --- | --- |
| Complement activation | 1.29E-06 |
| Angiogenesis | 9.87E-11 |
| Development of vasculature | 1.60E-10 |
| Vasculogenesis | 2.80E-09 |
| Formation of cellular protrusions | 8.74E-09 |
| Development of epithelial tissue | 9.29E-08 |
| Endothelial cell development | 6.67E-07 |
| Movement of vascular endothelial cells | 4.53E-06 |
| Biosynthesis of amide | 6.35E-06 |
| Secretion of mucus | 4.73E-05 |
| Synthesis of eicosanoid | 5.61E-05 |

## Discussion

There is an expansive literature on the clinical utility of EV biomarkers in the diagnosis and evaluation of clinical progression of a wide range of diseases. EV fingerprinting is being used to characterize the molecular basis of human diseases as well as for exploring biomarkers to determine surrogate clinical end points^24,25^. Most cell types in the body typically release around 6000-15,000 EVs per day^26-29^. Based on this estimate, a single cell sheds about 1/1000^th^ of its mass as EVs daily that could serve as surrogate nanobiopsy material reflecting the continuum of physiological status of the cell/tissue of origin. The EVs are a ‘multi-omic’ rich matrix (comprising transcriptome, proteome, lipidome, and metabolome).

The encoding molecular information within the EVs could provide a window into the salient biological networks underlying the pathobiological stages of disease (subclinical, preclinical and clinical disease). Hitherto, non-invasive EV based biomarker discovery for ILDs has largely focused on microRNA signatures whereas studies on EV protein biomarkers have been scanty. In this study, we determined whether EV protein cargo varied among the different types of ILDs. Interestingly, analysis of the EV proteome could divulge information about the disease-perturbed molecular networks constituting ILDs. Further, EV biomarkers organized into networks could serve an additional objective of revealing novel targets for pharmacological interventions for disease treatment, especially to treat fibrosing diseases where there is current dearth of therapeutic options.

Diagnosis of IPF is challenging; and IPF is unique among all other fibrotic lung diseases in that its etiological agents have not yet been discerned. IPF exhibits extensive molecular heterogeneity which is also associated with clinical, radiological patterns of the disease and prognosis^22,30,31^. Few other fibrotic lung diseases also exhibit pathological features similar to IPF posing challenges to the clinicians in making a definitive diagnosis. A vast majority of studies investigated the diagnostic efficacy of individual proteins for differential diagnosis, which showed poor sensitivity and specificity^5,7,9,12^. Given the etiological complexity of IPF, it is predicted that differential diagnosis relying on a single biomarker is doomed to fail. Notably, the current ATS/ERS/JRS/ALAT clinical practice guidelines strongly recommend against use of serum/plasma biomarkers such as MMP9, SFTPD, CCL18 and KL-6 for the purpose of distinguishing IPF from other ILDs, owing mainly to their poor efficacy^3^. In this study, we identified a five-protein signature, none of them overlap with the above-mentioned biomarkers, for differential diagnosis of IPF from other ILDs with greater specificity and sensitivity. This study describes a pipeline for mass spectrometry and ELISA based systematic discovery and development of plasma EV protein biomarkers that can be applied for other disease conditions as well. Though mass spectroscopy methods purposed for biomarker discovery currently identify a limited number of proteins per sample and generally quantify high abundant proteins, technical improvements in future may help in detecting very low abundant proteins as well.

Our protein signature comprised of SFTPB, ALDOA, HMGB1, CALML5 and TLN1. SFTPB levels in serum of IPF patients are known to be elevated^32^ and are associated with poor survival^33^. Similarly, proteomic analysis of broncho-alveolar lavage fluid (BALF) revealed upregulation of ALDOA in IPF patients presented with acute exacerbations^34^. In IPF patients, HMGB1 protein was elevated in inflammatory cells and in hyperplasic epithelial cells^35^. Serum HMGB1 levels have been reported to be high in IPF patients than in healthy subjects^36^. In addition, higher levels of HMGB1 in IPF are associated with acute exacerbations and poor survival^36^. We found that the five-protein panel in combination with lung function parameters achieved excellent sensitivity and specificity. However, as the study focused on the discovery and development of biomarkers enriched in plasma EVs for the differential diagnosis of IPF, the diagnostic power of the biomarkers based on their general expression profile in the plasma needs to be verified.

Next, we sought to understand the concordance between the EV protein expression profiles with the corresponding transcriptome profiles in IPF. We found that the expression profiles of 36 differentially expressed proteins (∼20% of total differentially expressed EV proteins) were in agreement with their corresponding mRNA expressions in IPF lung tissue. However, at this time, the cellular origin of the EV proteins could not be ascertained. We compared the signaling pathways associated with plasma EV proteins and tissue transcriptome profiles. The comparative analysis of EV proteomes and lung tissue transcriptomes discovered common pathways reflective of the fibrotic features of the disease suggesting that plasma EV proteome profiling could be useful in identifying dysregulated pathways. Importantly, the EV proteome corresponded with the transcriptome profiles in the gene expression patterns known to be dysregulated in IPF. Although several enriched pathways were simultaneously present in the EVs and lung tissue, there was a contrast in some of the biological information identified by the overrepresentation analysis between EV proteome and lung tissue transcriptome. For example, the analysis of EV proteins identified platelet degranulation pathway, a classic pathway in IPF pathogenesis promoting the development of fibrosis through the release of platelet derived growth factor^37^ and transforming growth factor-beta^38^. On the other hand, the transcriptome profiles demonstrated an enrichment of G protein-coupled receptor signaling which is known to promote IPF through profibrotic fibroblast activation. However, we speculate that the partial dissimilarity in the identified signaling pathways between EV proteome versus lung tissue transcriptome could be attributed to technicalities related to the substantially smaller number of differentially expressed EV proteins (274 proteins) and a relatively large transcriptome dataset (3432 transcripts) uploaded into overrepresentation analysis. As there was a significant overlap in the plasma EV proteins between IPF and non-IPF conditions, we set out to inquire whether the EV proteins signified different biological functions between IPF and CHP conditions. Based on the IPA analysis, the differentially expressed EV proteins in IPF showed cellular functions and pathways associated with adhesion of immune cells, fibrogenesis, binding of connective tissue cells, differentiation of epithelial tissue, and formation of focal adhesions. Contrarily, the analysis of EV proteins in CHP revealed cellular functions related to movement of granulocytes, angiogenesis, endothelial cell development, biosynthesis of amide and eicosanoids. These results suggest that despite significant overlap in the EV proteome between the two ILD conditions they could potentially provide distinct insights and guide toward a new appreciation of pathways differentially expressed in diseases.

## Conclusions

The diagnostic work up of IPF and other ILDs is still evolving and there is risk of misdiagnosis. To the best of our knowledge, this is the first EV based noninvasive protein signature for IPF diagnosis. In addition, the protein signature was discovered and validated in cohorts from two different geographic locations. However, we also acknowledge that the two retrospective cohorts used in this study are of small sample size. Therefore, the efficacy of the protein signature needs to be validated in large prospective cohorts.

## Supporting information

Supplementary methods and fifures

## Data Availability

All the relevant data was included in the supplementary data

## Abbreviations

ALAT: Latin Americ an Thoracic Society
ATS: American Thoracic Society
AUROC: area under the receiver operating characteristic curve
BALF: broncho-alveolar lavage fluid
CHP: chronic hypersensitivity pneumonitis
EDTA: Ethylenediaminetetraacetic acid
ELISA: enzyme-linked immunosorbent assay
ERS: European respiratory society
EVs: Extracellular vesicles
FDR: False discovery rate
HRCT: high-resolution computed tomography
HS: healthy subjects
ILDs: interstitial lung diseases
IPA: Ingenuity Pathway Analysis
IPF: Idiopathic pulmonary fibrosis
JRS: Japanese Respiratory Society
LASSO: Least Absolute Shrinkage and Selection Operator
NSIP: nonspecific interstitial pneumonia
UIP: usual interstitial pneumonia

## Acknowledgements

We thank Dr. Krishna Vankayalapati and his laboratory personnel at UTHSCT, Tyler for helping with material and equipment support and Dr. Daria Filanov, Alpha Nano Tech LLC, Durham, NC, USA for helping with EV size analysis. RSRA is considered as guarantor of the study and takes responsibility for the contents of the manuscript including data and analysis. Mass spectrometry analysis of EV samples was performed by Dr. Andrew Lemoff at Proteomics Core at UT Southwestern Medical center, Dallas, TX. We thank Maria Planchart, Brigham and Woman’s Hospital, Boston, MA for helping in collecting patient’s clinical data. This work was supported by UT System Rising STARs award and core funds from UTHSCT, Tyler, Texas, to NVK. DN received funding from NIH (R01 GM083122). Cryo-TEM is supported by Cancer Prevention and Research Institute of Texas grant RR140082 to DN. We thank Dr. Priyanka Sharma, MD Anderson Cancer center, Houston for critical reading and suggestions during manuscript preparation.

## Author contributions

NVK conceived the study. NVK and RSRA designed the experiments with inputs from KC, KVA, RV, AKP, SPdF, HI, YZ, DAS, LL, SC, DJK. RSRA, KC, KVA, RV and AKP performed experiments and analyzed data together with NVK, DN. RSRA and JWM performed statistical analysis. SPdF, FPdF, YH, HI, NH, YZ, KFG, LMS, DJK, IOR provided specimens and collected patient data. NVK and RSRA wrote original draft. All authors contributed in writing, reviewing and editing. NVK, IOR and DAS provided resources. NVK acquired funding and supervised the study.

