## Supplementary methods and fifures for "A novel protein signature from plasma extracellular vesicles for non-invasive differential diagnosis of idiopathic pulmonary fibrosis"

**Cryo-Electron microscopy of extracellular vesicles**

Four microliters of the EV solution (1:20 dilution) was added to Lacey carbon grids (200-mesh; Electron Microscopy Sciences) that were negatively glow-discharged for 30s at 30 mA. Excess sample was removed by blotting once ~2 seconds with Whatman filter paper and then the grid was plunge-frozen in liquid ethane cooled by liquid nitrogen using a homemade plunge-freezer.

The vitrified vesicle samples were imaged using a Titan Krios 300 kV transmission electron microscope (FEI, Hillsboro, OR, USA) equipped with a post-column Gatan imaging filter (Gatan Inc, Pleasanton, CA, USA), and a Volta Phase Plate (FEI). The SerialEM software was used to collect 2D images and tilt series under low-dose conditions.

Medium magnification maps (MMMs) and High magnification maps (HMMs) were recorded at 2250x magnification and 42000x magnification, respectively, without using dose fractionation. Tilt series were recorded at 42000x magnification on a K3 Summit direct electron detector (Gatan Inc, Pleasanton CA, USA) with an effective pixel size of 0.99 Å in dose fractionation mode. For each image, 20 frames were recorded over 1s exposure time at a dose rate of 26.03 electrons/pixel/s. The defocus was set to −0.5 μm (with phase plate) and the energy filter was in zero-loss mode with a slit width of 20 eV. The movie frames were aligned using MotionCorr2 (2).

**Nanoparticle tracking analysis of extracellular vesicles**

To measure the concentrations and the size distributions of EV preparations obtained from plasma samples, EVs were diluted 1:50 with 1X PBS, pH 7.4 and analyzed on Nanosight NS300 instrument (Malvern, MA, USA), which was equipped with a 488 nm laser and a sCMOS camera. Three videos of 60s each were recorded for each sample at 25°C to estimate mean values of particle concentration.

**Lysis of extracellular vesicles, protein estimation and western blot**

The EV fractions from SEC were concentrated down to 50 µL in vacuum at 30°C, lysed in RIPA buffer (Sigma Aldrich, USA) at 95°C for 10 min and sonicated in ultrasonic sonicator bath (Branson Ultrasonics Corp, CT, USA) for 2 min. The supernatants containing the EV proteins were collected after centrifugation at 30000 g for 15 min at 4°C. The protein concentration was estimated using Pierce BCA protein assay kit (Thermo Fisher Scientific, MA, USA) using bovine serum albumin as standard. Western blot was performed following SDS PAGE using standard protocols involving ALIX (Cat# 2171) and CD9 (Cat#131714) antibodies from Cell Signaling Technologies, MA, USA; HSP70 (Cat# SC-24) and Calnexin (Cat# SC-23954) antibodies from Santa Cruz Biotechnology, CA, USA; Proteins were detected by chemiluminiscence using Clarity Max ECL substrate (Bio-Rad Laboratories, CA, USA) on Chemidoc-MP Gel imaging system (Bio-Rad Laboratories).

**Biomarker discovery:**

Protein abundance values obtained from mass spectrometry are suitable for determining relative protein amounts between samples^1-3^. The protein abundance values were log_2_(n+1) transformed prior to differential expression analysis. Proteins detected in less than 10% of the samples were removed from further analysis. A t-test was used for identifying differentially expressed genes between different classes of samples, using an FDR cut off value of 0.05 to correct for multiple hypothesis testing. Proteins were considered differentially expressed if both (A) the t-test indicates significant differential expression and (B) |log_2_ fold change| ≥0.585 (equivalent to 50% increase or decrease in expression). Glmnet package^4^ was used for performing LASSO regression. Five-fold cross validation was performed using cv.glmnet function to estimate cross validation error at varying values of tuning parameter (λ). Features included in the binomial model at λ_min_ were selected as protein signature.

**Biomarker validation:**

Biomarker validation was performed using Enzyme-linked immunosorbent assay (ELISA). ELISA was performed on standard sandwich ELISA kits (Aviva Systems Biology, CA, USA) as per manufacturer’s instructions and the microplates were read on Synergy microplate reader (BioTek Instruments Ltd, VT, USA). SPSS package was used for constructing binomial logistic regression models and ROC curves. Hosmer and Lemeshow (HL) test was used for logistic regression model calibration. A p-value greater than 0.05 was considered as appropriately calibrated model. Individual risk scores were calculated for each subject in the logistic regression model by subtracting Youden index value of the regression model from predicted probability value of the subject.

**Transcriptome analysis**

For comparing transcriptomic changes in lung tissue of CHP and IPF with healthy subjects, we used raw read counts of a large RNA sequencing data from a previous study (GSE150910) (Furasuwa, Schwartz 2020). We analyzed 103 IPF samples and 103 healthy subjects. Raw read counts of transcripts were collapsed to gene IDs using tximport package. Data transformations, normalization and differential expression analysis were carried out using DESeq2. Genes showing a minimum log2 fold change of 1 at FDR<0.01 were considered differentially expressed. Pathway analysis was performed on differentially expressed genes using Ingenuity pathway analysis module (Qiagen, MD, USA) and g:profiler package^5^.

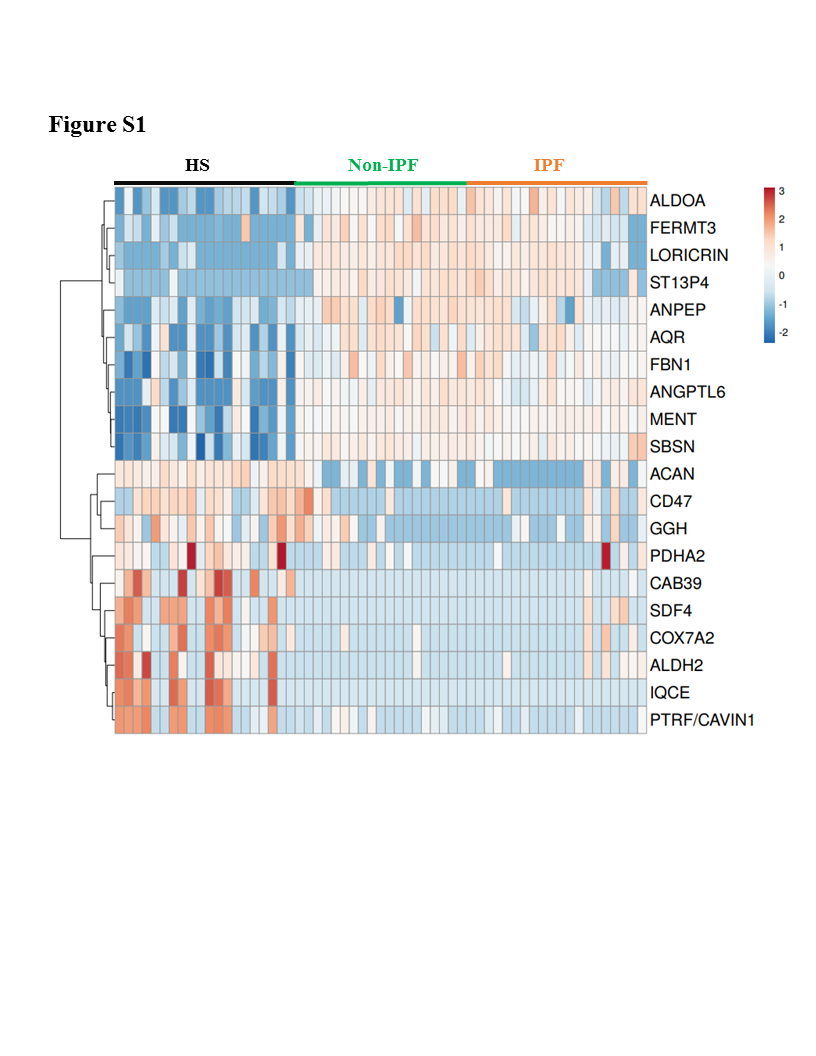

**Figure S1.** Heatmap showing top ten up- and downregulated proteins in a comparison between healthy subjects against all ILDs.

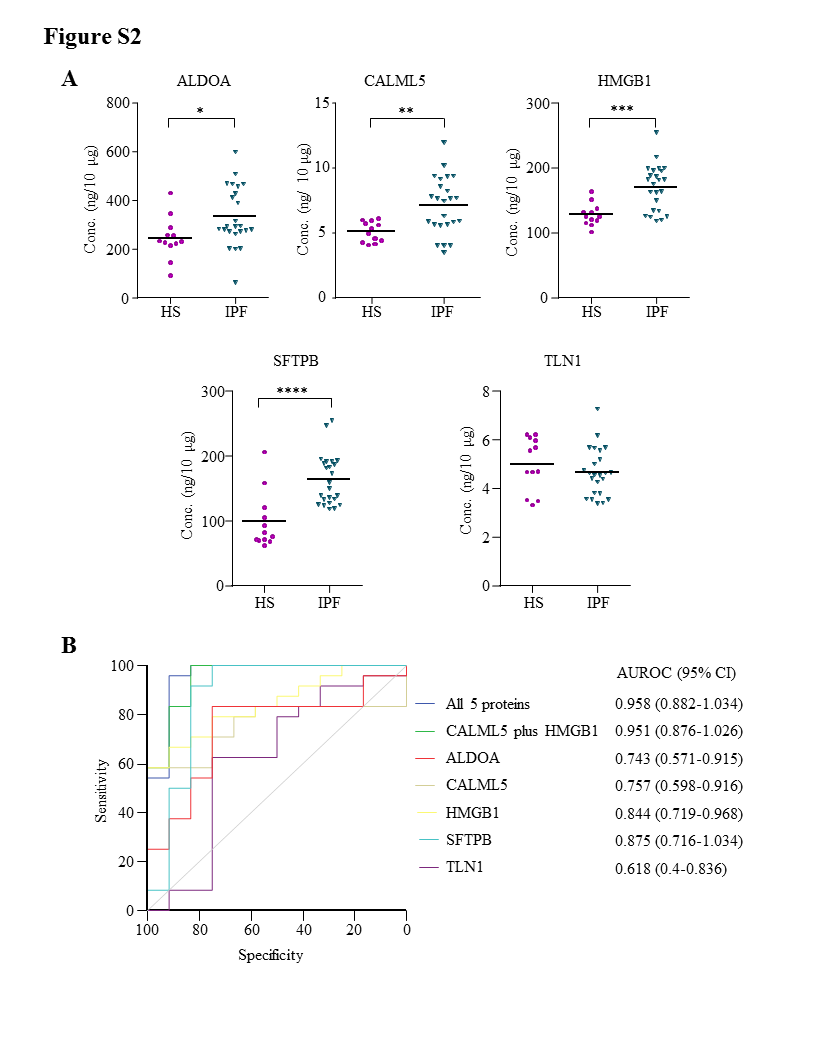

**Figure S2. A)** The levels of five proteins in plasma EVs of 24 IPF and 12 HS was assesed using sandwich ELISA with purified recombinant human protein as standard. The asterisks *, **, *** and **** denote p<0.05, p<0.01, p<0.001 and p<0.0001, respectively. **B)** AUROC curves for logistic regression model generated for five proteins on 24 IPF patients and 12 healthy subjects.
