## Supplementary tables for "A novel protein signature from plasma extracellular vesicles for non-invasive differential diagnosis of idiopathic pulmonary fibrosis"

**Table S1. Differentially expressed proteins identified in comparision of proteomic profiles of plasma EVs from IPF and non-IPF**

| **Protein symbol** | **FDR(BH)** | **Log2FC** |
| --- | --- | --- |
| CEP290 | 0.031835012 | -3.67489 |
| PTPRG | 0.013379933 | -2.42574 |
| DCDC1/DCDC5 | 0.006411218 | -2.28237 |
| UTP14C | 0.047131114 | -1.78836 |
| CRTAC1 | 0.006411218 | -1.12103 |
| EMD | 0.006411218 | -1.07538 |
| ATRN | 0.034193161 | -0.92917 |
| SDF4 | 0.006411218 | -0.77978 |
| MT-CO2 | 0.006411218 | -0.59182 |
| FLG2 | 0.01079784 | 0.650379 |
| TXN | 0.044878524 | 0.748756 |
| MASP1 | 0.029308424 | 0.780045 |
| VWF | 0.039708187 | 0.810987 |
| DSG1 | 0.01079784 | 0.819757 |
| CSTA | 0.047131114 | 0.835759 |
| ALDOA | 0.036208758 | 1.122451 |
| SFTPD | 0.018996201 | 1.146431 |
| A2ML1 | 0.046627038 | 1.228911 |
| LCN1 | 0.013379933 | 1.440185 |
| ARG1 | 0.006411218 | 1.643993 |
| HMGB1 | 0.006411218 | 1.714167 |
| LAMA2 | 0.018996201 | 1.78414 |
| SDR9C7 | 0.017096581 | 1.985864 |
| ALOX12B | 0.047131114 | 2.036869 |
| CALML5 | 0.01079784 | 2.070101 |
| TLN1 | 0.048590282 | 2.342485 |
| SERF2 | 0.006411218 | 2.564957 |
| TPP1 | 0.018996201 | 2.575143 |
| SFTPB | 0.006411218 | 3.83857 |
| PRRC2C | 0.006411218 | 3.916179 |

Log2FC denotes log2 fold change in expression of proteins in IPF over non-IPF

**Table S2. Differentially expressed proteins identified in comparision of proteomic profiles of plasma EVs from IPF and HS**

| **Protein symbol** | **FDR (BH)** | **Log2FC** |
| --- | --- | --- |
| PTRF/CAVIN1 | 0.003620452 | -5.666933957 |
| CAB39 | 0.001486659 | -4.381413748 |
| IQCE | 0.001486659 | -4.373319927 |
| ACAN | 0.001486659 | -4.119451163 |
| PDHA2 | 0.012723037 | -3.473594016 |
| GGH | 0.002596949 | -3.33544014 |
| COX7A2 | 0.007967339 | -3.164063015 |
| ANXA5 | 0.00907783 | -3.147676362 |
| CD47 | 0.004412021 | -3.095307394 |
| SDF4 | 0.012021033 | -3.05301955 |
| TMX2 | 0.013111663 | -2.823236916 |
| EZR | 0.008628181 | -2.752554205 |
| ALDH2 | 0.01914817 | -2.653780446 |
| LGALS3 | 0.01034415 | -2.498942007 |
| PYCARD | 0.011488902 | -2.242476365 |
| DSC3 | 0.023672189 | -2.194342238 |
| RAC2 | 0.027741621 | -2.194085271 |
| ACOX1 | 0.011488902 | -2.149811998 |
| PPIB | 0.036135955 | -2.040563177 |
| DSTN | 0.028034145 | -1.956803993 |
| TFPI | 0.017096581 | -1.938021551 |
| C12orf5/TIGAR | 0.00907783 | -1.937430867 |
| GNAI2 | 0.001486659 | -1.921477572 |
| SERPINB7 | 0.014654212 | -1.918516255 |
| MME | 0.021454533 | -1.811411479 |
| PABPC1 | 0.017096581 | -1.777446767 |
| SSR4 | 0.01744549 | -1.711124382 |
| SULT2B1 | 0.049429492 | -1.543917032 |
| LCN1 | 0.013576696 | -1.53072172 |
| PTPRZ1 | 0.039104886 | -1.522787413 |
| PRKCSH | 0.046627038 | -1.348275827 |
| ARG1 | 0.013111663 | -1.224671576 |
| INHBE | 0.001486659 | 0.725132102 |
| CD5L | 0.028772294 | 0.72728206 |
| PRTN3 | 0.001486659 | 0.806001118 |
| LGALS3BP | 0.014654212 | 0.830213263 |
| PODXL | 0.007967339 | 0.928297062 |
| SERPINF2 | 0.033797865 | 0.972978392 |
| HSPB1 | 0.028034145 | 0.978060655 |
| COLEC10 | 0.004412021 | 0.979694661 |
| CAMP | 0.046676379 | 0.985650991 |
| PRG4 | 0.004412021 | 1.009141731 |
| ATRN | 0.001486659 | 1.048123896 |
| DSC1 | 0.00539892 | 1.116570549 |
| PRDX2 | 0.013111663 | 1.144240558 |
| ITIH2 | 0.002596949 | 1.158366788 |
| SHBG | 0.023163109 | 1.18049439 |
| FCN1 | 0.013111663 | 1.199905377 |
| MBL2 | 0.006411218 | 1.228179912 |
| TTR | 0.035170109 | 1.257762323 |
| LBP | 0.012021033 | 1.350378468 |
| PFKL | 0.001486659 | 1.386518849 |
| TTN | 0.045467663 | 1.390696537 |
| AP1B1 | 0.01802072 | 1.405068049 |
| GGCT | 0.044042279 | 1.409217427 |
| PROL1/OPRPN | 0.011488902 | 1.43484314 |
| FETUB | 0.00907783 | 1.474320737 |
| SERF2 | 0.029661538 | 1.477267705 |
| HEG1 | 0.001486659 | 1.501683756 |
| ADIPOQ | 0.014654212 | 1.535035386 |
| FBLN1 | 0.028319489 | 1.557436188 |
| PFKP | 0.020920816 | 1.596222092 |
| ZNF511-PRAP1 | 0.026788273 | 1.627256386 |
| JUP | 0.001486659 | 1.657782473 |
| SFTPD | 0.008628181 | 1.673492131 |
| FLG2 | 0.001486659 | 1.673914277 |
| HAS2 | 0.001486659 | 1.727245524 |
| PCYOX1 | 0.001486659 | 1.744199863 |
| LGALS7/LGALS7B | 0.01744549 | 1.744686081 |
| MUC7 | 0.001486659 | 1.753862109 |
| CARHSP1 | 0.030098172 | 1.779325031 |
| DSP | 0.001486659 | 1.799033411 |
| SFN | 0.00907783 | 1.810123878 |
| SSC5D | 0.046627038 | 1.844957099 |
| LTF | 0.001486659 | 1.853745453 |
| RELN | 0.004412021 | 1.858939498 |
| CALML3 | 0.046970606 | 1.933699484 |
| DCDC1/DCDC5 | 0.002596949 | 1.962198005 |
| SLC4A1 | 0.007967339 | 1.975159453 |
| LAMC1 | 0.002596949 | 1.999065628 |
| CALR | 0.041144518 | 2.007967374 |
| PFN1 | 0.01034415 | 2.025607516 |
| CDSN | 0.037670432 | 2.039504612 |
| ATP5F1A | 0.039104886 | 2.048283187 |
| AMPD3 | 0.001486659 | 2.06651307 |
| GGT2 | 0.033797865 | 2.081791819 |
| PKP1 | 0.001486659 | 2.11119167 |
| CD226 | 0.046676379 | 2.119430167 |
| PTBP1 | 0.03318748 | 2.126686803 |
| LAP3 | 0.011488902 | 2.134309946 |
| TNC | 0.030529608 | 2.137049461 |
| ARHGAP1 | 0.002596949 | 2.13995453 |
| ITGA2B | 0.01914817 | 2.176526477 |
| BIN2 | 0.001486659 | 2.180326705 |
| NAMPT | 0.034782699 | 2.194424942 |
| FCN2 | 0.001486659 | 2.198053938 |
| SERPINA1 | 0.016614987 | 2.219879066 |
| SVEP1 | 0.001486659 | 2.245130212 |
| YWHAH | 0.022647419 | 2.268076851 |
| CSTA | 0.001486659 | 2.268788253 |
| SPARCL1 | 0.001486659 | 2.279597295 |
| CREG1 | 0.008628181 | 2.299255033 |
| EVPL | 0.027741621 | 2.307517025 |
| GSN | 0.00989802 | 2.316179365 |
| RAP1B | 0.001486659 | 2.316363311 |
| BLMH | 0.001486659 | 2.345994708 |
| AMY1A | 0.038611379 | 2.351644796 |
| GC | 0.012021033 | 2.387626892 |
| CCDC73 | 0.043161859 | 2.388609701 |
| CST4 | 0.033797865 | 2.393102066 |
| NCCRP1 | 0.001486659 | 2.411709151 |
| SELP | 0.02038003 | 2.418575182 |
| CALM3/CALM2/CALM1 | 0.011488902 | 2.451200005 |
| BCHE | 0.001486659 | 2.478956457 |
| SH3BGRL3 | 0.028319489 | 2.487311257 |
| MARCO | 0.001486659 | 2.492467085 |
| ACTN1 | 0.003620452 | 2.527390604 |
| CFP | 0.001486659 | 2.53133354 |
| LDHB | 0.03318748 | 2.541302887 |
| SNRPD3 | 0.001486659 | 2.582340095 |
| VTN | 0.001486659 | 2.595270266 |
| DMBT1 | 0.014654212 | 2.615476584 |
| COMP | 0.001486659 | 2.627172139 |
| MUC5B | 0.001486659 | 2.630800453 |
| GM2A | 0.01034415 | 2.650262963 |
| SFTPA1 | 0.013576696 | 2.673683105 |
| ZCCHC12 | 0.029661538 | 2.700839471 |
| CALML5 | 0.001486659 | 2.705746557 |
| YWHAE | 0.013576696 | 2.721711229 |
| CASP14 | 0.001486659 | 2.741079033 |
| IL36G | 0.003620452 | 2.773611853 |
| GAPDH | 0.002596949 | 2.815185614 |
| SELPLG | 0.047798817 | 2.822414615 |
| PTPRJ | 0.01034415 | 2.830497785 |
| CFL1 | 0.001486659 | 2.847777762 |
| FLG | 0.001486659 | 2.86687653 |
| ASAH1 | 0.007327106 | 2.869175918 |
| DDX60L | 0.00907783 | 2.885247358 |
| IVL | 0.008628181 | 2.89779604 |
| CSNK1A1L | 0.001486659 | 2.976093196 |
| TNXB | 0.003620452 | 2.985342212 |
| CAPN1 | 0.004412021 | 3.000917228 |
| DHTKD1 | 0.001486659 | 3.001083751 |
| ZYX | 0.002596949 | 3.030698927 |
| STAT3 | 0.002596949 | 3.033865849 |
| PRR4 | 0.001486659 | 3.044957398 |
| SPRR3 | 0.049152669 | 3.051918908 |
| CAPG | 0.007967339 | 3.053724171 |
| CALD1 | 0.007327106 | 3.054779469 |
| CHL1 | 0.001486659 | 3.065000969 |
| CSTB | 0.00907783 | 3.093301651 |
| ALOX12B | 0.001486659 | 3.105806079 |
| ACTN4 | 0.01034415 | 3.10816556 |
| LCAT | 0.007967339 | 3.114615482 |
| CAP1 | 0.01744549 | 3.122920986 |
| C1QTNF3-AMACR | 0.001486659 | 3.155674131 |
| TMCO1 | 0.001486659 | 3.164268926 |
| TGM1 | 0.001486659 | 3.164845819 |
| TGM3 | 0.001486659 | 3.176465477 |
| HSPG2 | 0.002596949 | 3.335028031 |
| RAB15 | 0.001486659 | 3.371794877 |
| HMGB1 | 0.001486659 | 3.392365043 |
| SLC2A2 | 0.004412021 | 3.428547041 |
| HMCN1 | 0.003620452 | 3.430668097 |
| CRNN | 0.002596949 | 3.434458377 |
| ABCC2 | 0.001486659 | 3.48056511 |
| VIM | 0.001486659 | 3.508576381 |
| CARD9 | 0.003620452 | 3.649198867 |
| APMAP | 0.004412021 | 3.691991418 |
| HRNR | 0.001486659 | 3.752622398 |
| SFTPB | 0.001486659 | 3.863922068 |
| VCL | 0.001486659 | 3.939590394 |
| CKMT1A | 0.001486659 | 4.084069522 |
| PNP | 0.001486659 | 4.084367923 |
| NR0B1 | 0.001486659 | 4.150251825 |
| GPR126/ADGRG6 | 0.001486659 | 4.548325365 |
| FBN1 | 0.001486659 | 4.855105716 |
| SBSN | 0.001486659 | 5.211991792 |
| ANPEP | 0.001486659 | 5.24179039 |
| AQR | 0.001486659 | 5.387815584 |
| ANGPTL6 | 0.001486659 | 5.424441222 |
| MENT | 0.001486659 | 5.437532738 |
| FERMT3 | 0.001486659 | 6.113355725 |
| ST13P4 | 0.001486659 | 6.281156031 |
| LORICRIN | 0.001486659 | 7.251469458 |
| ALDOA | 0.001486659 | 9.172324908 |

Log2FC denotes log2 fold change in expression of proteins in IPF over HS

**Table S3. Differentially expressed genes identified in comparison of transcriptome profiles of lung tissues from IPF and HS**

| **Gene name** | **padj** | **Log2FC** |
| --- | --- | --- |
| CT45A1 | -4.898522489 | 1.59E-09 |
| CT45A3 | -4.881880783 | 8.68E-12 |
| CT45A10 | -4.401202976 | 8.42E-10 |
| DEFA1B | -3.967471644 | 2.00E-12 |
| DEFA4 | -3.957564485 | 7.04E-26 |
| SLCO1A2 | -3.79849276 | 1.10E-63 |
| ITLN2 | -3.794258668 | 1.65E-48 |
| DEFA1 | -3.646815384 | 3.83E-17 |
| CT45A5 | -3.262834768 | 1.79E-07 |
| HELT | -2.977760851 | 6.04E-28 |
| HMGCS2 | -2.939426836 | 2.45E-25 |
| FAM167A | -2.916030121 | 1.86E-56 |
| APOA5 | -2.891035567 | 2.67E-23 |
| RTKN2 | -2.890913842 | 4.13E-42 |
| CCK | -2.752283623 | 2.04E-30 |
| GRM8 | -2.701519726 | 6.06E-37 |
| APOC3 | -2.669612199 | 4.60E-06 |
| SLC6A4 | -2.63781308 | 2.79E-19 |
| PRG3 | -2.62443513 | 3.63E-05 |
| TRIM10 | -2.61637637 | 6.12E-11 |
| FAM71A | -2.579027192 | 2.01E-09 |
| HTR3C | -2.565261705 | 2.33E-25 |
| CA4 | -2.540370361 | 1.47E-28 |
| MS4A15 | -2.527829702 | 1.07E-44 |
| KLRG2 | -2.516731849 | 2.71E-68 |
| LAMP3 | -2.492335911 | 1.36E-11 |
| ADRA1A | -2.484932405 | 4.09E-57 |
| MAP3K15 | -2.481688261 | 6.72E-66 |
| IL17A | -2.469894705 | 6.63E-07 |
| KLK5 | -2.394945912 | 2.19E-10 |
| SLC14A1 | -2.346118092 | 1.49E-26 |
| ADCY8 | -2.336271042 | 8.51E-30 |
| CSF3 | -2.30373321 | 3.53E-13 |
| SPINT3 | -2.30261583 | 0.001492864 |
| AL096711.2 | -2.288865157 | 3.13E-54 |
| LIM2 | -2.269807389 | 3.49E-17 |
| RGS9BP | -2.267340822 | 1.35E-47 |
| AGER | -2.259481414 | 1.08E-42 |
| TRIML1 | -2.259191696 | 2.51E-11 |
| LY6G6F | -2.235660333 | 3.28E-15 |
| RNASE3 | -2.234116272 | 3.45E-20 |
| FCN3 | -2.221586681 | 1.03E-25 |
| SEMA5A | -2.201692499 | 2.73E-21 |
| PRX | -2.200996994 | 6.03E-40 |
| MPO | -2.173489532 | 1.06E-24 |
| OLAH | -2.169065094 | 4.64E-26 |
| PNMT | -2.167135103 | 5.85E-25 |
| UNC5D | -2.154127721 | 1.56E-20 |
| CD177 | -2.1280835 | 4.37E-06 |
| MYZAP | -2.104218737 | 3.80E-29 |
| BTNL9 | -2.101161967 | 3.00E-29 |
| MYRF | -2.098116931 | 1.33E-41 |
| ALPP | -2.091865759 | 4.95E-23 |
| FABP12 | -2.089366644 | 2.91E-08 |
| CACNA1S | -2.07821514 | 9.22E-24 |
| KIR3DL1 | -2.061176412 | 3.16E-26 |
| VIPR1 | -2.060597053 | 1.07E-22 |
| LY6G6E | -2.058120947 | 1.76E-13 |
| GGTLC1 | -2.055864957 | 8.19E-33 |
| KLRF1 | -2.0451008 | 6.17E-28 |
| HBZ | -2.044178627 | 4.91E-18 |
| LRIT1 | -2.028039986 | 7.01E-07 |
| ARG1 | -2.006920952 | 7.54E-20 |
| LY6G6F-LY6G6D | -2.005629802 | 1.53E-16 |
| MGAM2 | -2.000794883 | 1.53E-14 |
| BTNL8 | -1.992861458 | 5.11E-46 |
| RS1 | -1.983057106 | 7.12E-30 |
| GHRHR | -1.966976608 | 1.97E-17 |
| RBP2 | -1.964343546 | 1.05E-26 |
| IL1RL1 | -1.954160799 | 1.26E-15 |
| AC025263.2 | -1.93498163 | 0.008442756 |
| POPDC3 | -1.924742062 | 5.85E-24 |
| CABP5 | -1.919703274 | 1.50E-11 |
| FAM19A4 | -1.917939633 | 0.000801614 |
| NKD1 | -1.90685471 | 3.40E-27 |
| SH2D1B | -1.903931537 | 1.26E-11 |
| RXFP1 | -1.901879823 | 1.04E-30 |
| C4orf51 | -1.882818854 | 2.30E-09 |
| SNX22 | -1.88082508 | 6.24E-28 |
| C2orf91 | -1.873162319 | 1.16E-24 |
| ASPG | -1.871634137 | 5.48E-26 |
| HSD17B6 | -1.869093157 | 8.45E-43 |
| BTNL3 | -1.867549449 | 2.23E-08 |
| CAMP | -1.864698836 | 6.87E-22 |
| QRFPR | -1.861688306 | 1.95E-06 |
| GNLY | -1.854956675 | 1.21E-27 |
| WFDC12 | -1.852121735 | 1.40E-35 |
| TAS2R40 | -1.851960171 | 9.66E-07 |
| KRT73 | -1.849556662 | 2.31E-16 |
| BRINP1 | -1.844767734 | 0.001322963 |
| OR5P3 | -1.839163634 | 4.00E-08 |
| PTPRQ | -1.838865441 | 4.85E-26 |
| ZAR1 | -1.836163528 | 1.35E-11 |
| TRIM15 | -1.830391927 | 0.002426892 |
| SLC13A2 | -1.81618606 | 6.47E-10 |
| STXBP6 | -1.815792269 | 6.31E-38 |
| CAV3 | -1.812695553 | 3.98E-28 |
| GRID2 | -1.812446492 | 4.69E-14 |
| ACADL | -1.810915274 | 1.06E-26 |
| GRIA1 | -1.806296312 | 1.85E-47 |
| GCOM1 | -1.800557007 | 9.83E-32 |
| LRRN3 | -1.79970154 | 1.10E-35 |
| C8B | -1.794489202 | 1.13E-12 |
| CRTAC1 | -1.783253173 | 6.13E-30 |
| NECAB1 | -1.776858725 | 5.50E-31 |
| CALCR | -1.775140002 | 1.33E-07 |
| ANKRD1 | -1.773590448 | 1.78E-10 |
| PRSS57 | -1.770391022 | 8.00E-24 |
| RNF182 | -1.766603211 | 1.48E-51 |
| AGBL1 | -1.763226786 | 1.36E-19 |
| CHRM2 | -1.762184825 | 6.56E-16 |
| CSAG3 | -1.758362199 | 1.60E-10 |
| FXYD4 | -1.757924398 | 2.49E-15 |
| KCNS1 | -1.756448472 | 1.02E-23 |
| NCKAP5 | -1.752503629 | 7.34E-28 |
| NCR1 | -1.74600921 | 8.55E-33 |
| TINCR | -1.745908539 | 3.19E-47 |
| ODAM | -1.743268905 | 9.15E-25 |
| MME | -1.735272366 | 4.66E-34 |
| NRG3 | -1.728224933 | 2.87E-22 |
| GP9 | -1.725861094 | 7.87E-22 |
| AFF3 | -1.725171849 | 2.29E-52 |
| OTULINL | -1.720527684 | 3.43E-49 |
| KRT72 | -1.719981773 | 3.99E-14 |
| CEACAM8 | -1.715620722 | 3.26E-05 |
| KIR2DL3 | -1.713748373 | 3.76E-21 |
| NOS2 | -1.713473866 | 7.43E-22 |
| SP6 | -1.711275837 | 4.14E-17 |
| HIF3A | -1.705348316 | 1.33E-26 |
| TRIM40 | -1.696825996 | 2.85E-10 |
| NDRG4 | -1.694208196 | 1.44E-31 |
| TMEM100 | -1.690054233 | 4.34E-14 |
| AGRP | -1.687896767 | 3.81E-22 |
| FAM107A | -1.683431182 | 3.02E-17 |
| CA1 | -1.68243083 | 6.67E-15 |
| PLPPR5 | -1.681017087 | 4.45E-05 |
| ZDHHC19 | -1.669816677 | 5.45E-26 |
| NXF3 | -1.66879047 | 1.07E-22 |
| TUBB1 | -1.661749434 | 2.28E-20 |
| ARC | -1.65712428 | 4.61E-08 |
| C19orf84 | -1.653989773 | 8.62E-05 |
| SGCG | -1.650929611 | 6.80E-26 |
| GALNT13 | -1.644552438 | 8.73E-17 |
| PLA2G12B | -1.64116026 | 4.15E-16 |
| F11 | -1.634228324 | 1.31E-16 |
| CD300LG | -1.628027513 | 1.61E-21 |
| RNF128 | -1.625919531 | 2.07E-13 |
| ZNF385B | -1.623259954 | 3.34E-24 |
| VEGFD | -1.616745378 | 7.19E-07 |
| SLC5A9 | -1.614691707 | 1.48E-30 |
| DPP6 | -1.613982859 | 1.78E-26 |
| C13orf42 | -1.613377341 | 1.65E-15 |
| EPB41L5 | -1.612000772 | 2.21E-62 |
| FIBIN | -1.607883092 | 1.92E-32 |
| CNTN6 | -1.607603355 | 3.48E-29 |
| MATN3 | -1.605731851 | 1.32E-13 |
| CTNND2 | -1.602663832 | 2.43E-28 |
| NFE2 | -1.60064263 | 9.27E-20 |
| WNT7A | -1.598866408 | 8.47E-33 |
| SH3GL3 | -1.597132934 | 3.09E-20 |
| CSAG2 | -1.595285644 | 3.15E-07 |
| FAM189A1 | -1.594301353 | 6.80E-31 |
| ST8SIA6 | -1.593020773 | 9.66E-28 |
| LHX9 | -1.592575448 | 4.37E-07 |
| FO681492.1 | -1.586679581 | 1.53E-34 |
| AL662899.1 | -1.585961885 | 1.50E-11 |
| CLRN2 | -1.585890195 | 0.006210619 |
| PF4 | -1.585272135 | 2.75E-14 |
| TNNC1 | -1.581063066 | 1.38E-26 |
| KIAA0408 | -1.571887859 | 7.14E-32 |
| ESM1 | -1.569650043 | 2.27E-08 |
| AMER2 | -1.568164758 | 1.82E-13 |
| PRSS41 | -1.564431669 | 2.38E-08 |
| DAPK2 | -1.564043792 | 4.30E-45 |
| ADRB1 | -1.560471519 | 1.15E-26 |
| SOSTDC1 | -1.555069725 | 5.60E-14 |
| SYN2 | -1.552250482 | 6.40E-29 |
| KCNMB4 | -1.549835531 | 1.28E-16 |
| KIR2DL1 | -1.547868555 | 0.000229626 |
| PSKH2 | -1.544498426 | 0.010214969 |
| APOA1 | -1.542238337 | 4.62E-31 |
| KLK3 | -1.537176116 | 0.006718528 |
| LTK | -1.534449583 | 2.25E-23 |
| MYL1 | -1.533090223 | 3.92E-05 |
| CA2 | -1.529728792 | 9.14E-17 |
| CASKIN2 | -1.528683641 | 3.74E-24 |
| HHIP | -1.521056573 | 2.13E-17 |
| CHRM1 | -1.519283818 | 4.46E-05 |
| MS4A3 | -1.517298846 | 2.12E-12 |
| HBA2 | -1.509419083 | 7.61E-08 |
| HPCAL4 | -1.496471121 | 4.31E-14 |
| ERICH4 | -1.496469615 | 9.08E-19 |
| FATE1 | -1.494618338 | 3.96E-12 |
| FOLR3 | -1.493119087 | 1.38E-21 |
| EFCC1 | -1.492506343 | 9.12E-42 |
| GPR158 | -1.492259316 | 1.29E-14 |
| LYZL4 | -1.49071076 | 0.001305727 |
| DPRX | -1.490079776 | 4.93E-07 |
| PCSK9 | -1.48979396 | 3.49E-20 |
| EDNRB | -1.484142679 | 3.78E-44 |
| C18orf63 | -1.48174826 | 6.51E-10 |
| SCN1A | -1.48148013 | 2.75E-19 |
| TRPC3 | -1.479935887 | 5.71E-12 |
| DYTN | -1.476678849 | 0.00016131 |
| CYP4A22 | -1.475414021 | 5.22E-10 |
| PRLH | -1.464058479 | 0.001518276 |
| CHRM3 | -1.45994329 | 3.61E-31 |
| MYMK | -1.457199051 | 4.19E-06 |
| AC067752.1 | -1.456171616 | 1.50E-12 |
| ADAM29 | -1.456166233 | 1.82E-18 |
| TTPA | -1.454180933 | 1.25E-33 |
| LRP1B | -1.451779252 | 1.43E-10 |
| UPB1 | -1.451477152 | 4.09E-33 |
| GPM6A | -1.448228724 | 1.21E-35 |
| SLC19A3 | -1.44242473 | 4.40E-24 |
| TNNT1 | -1.439463188 | 2.71E-15 |
| AATK | -1.439081201 | 5.30E-36 |
| MUCL3 | -1.438447996 | 1.18E-13 |
| CDH13 | -1.435672764 | 8.90E-33 |
| CLIC3 | -1.43122107 | 8.01E-25 |
| ANKRD29 | -1.429034366 | 2.80E-34 |
| INSC | -1.42882105 | 9.49E-18 |
| ARHGEF26 | -1.426389764 | 4.68E-42 |
| RNASE8 | -1.425014008 | 0.00270824 |
| COL4A3 | -1.42447193 | 5.63E-18 |
| WWC2 | -1.420978999 | 6.81E-51 |
| GPRIN2 | -1.419609711 | 1.19E-31 |
| GSG1L | -1.415645044 | 0.000210425 |
| OLFML2A | -1.415602992 | 1.79E-24 |
| KCTD16 | -1.415468125 | 3.23E-19 |
| WFDC5 | -1.414868192 | 1.21E-07 |
| OR5P2 | -1.411472186 | 3.13E-06 |
| ANKRD34B | -1.411042279 | 0.030632512 |
| C16orf78 | -1.408170186 | 6.06E-09 |
| LAMC3 | -1.407798364 | 6.47E-16 |
| LRRC52 | -1.406939249 | 1.39E-08 |
| S100A8 | -1.406135305 | 4.65E-11 |
| S1PR5 | -1.403977854 | 2.44E-20 |
| PLA2G1B | -1.403187236 | 3.28E-10 |
| SPOCK2 | -1.397398651 | 2.08E-24 |
| GPAM | -1.392005226 | 3.49E-09 |
| SH2D3C | -1.388385444 | 1.42E-41 |
| KLRC3 | -1.388320625 | 5.00E-11 |
| OR52K2 | -1.387922973 | 0.000441625 |
| HEMGN | -1.386099541 | 1.70E-10 |
| SYCP2L | -1.38411659 | 1.35E-17 |
| CACNA2D2 | -1.383656304 | 9.71E-20 |
| CADM2 | -1.382016923 | 1.09E-14 |
| ANXA3 | -1.381050911 | 9.42E-30 |
| IBA57 | -1.380674309 | 1.92E-25 |
| HTR1D | -1.380430714 | 3.22E-22 |
| SLFNL1 | -1.380019215 | 7.02E-24 |
| CX3CR1 | -1.379998837 | 5.84E-13 |
| OR6N1 | -1.379511847 | 2.26E-07 |
| LRRC3B | -1.378938846 | 2.77E-08 |
| SLC39A8 | -1.364629157 | 3.90E-34 |
| F2RL3 | -1.364066741 | 1.39E-08 |
| LILRA1 | -1.360360578 | 6.64E-22 |
| TKTL1 | -1.358301287 | 7.92E-13 |
| OVCH2 | -1.357147077 | 2.40E-14 |
| PCDH12 | -1.355736107 | 7.94E-32 |
| CYP3A5 | -1.355500298 | 3.00E-22 |
| ARPP21 | -1.354607593 | 0.000813936 |
| JCAD | -1.354492852 | 1.27E-13 |
| C13orf46 | -1.350451786 | 1.76E-21 |
| OR6N2 | -1.349445185 | 0.001661205 |
| TMIE | -1.347909331 | 2.52E-30 |
| PTPRB | -1.346801921 | 1.03E-33 |
| ALAS2 | -1.34078627 | 5.18E-07 |
| SEC14L4 | -1.338932856 | 2.87E-22 |
| EDN3 | -1.33688377 | 1.63E-08 |
| PEBP4 | -1.334180919 | 1.14E-12 |
| LCN1 | -1.333886181 | 9.28E-06 |
| CAV1 | -1.332699334 | 4.41E-37 |
| CCDC54 | -1.331677443 | 1.50E-15 |
| C2orf72 | -1.331046773 | 8.34E-22 |
| AFF2 | -1.329863847 | 1.34E-25 |
| PLPPR1 | -1.327411044 | 5.42E-12 |
| CCDC68 | -1.325676397 | 6.14E-22 |
| MAG | -1.323191311 | 1.32E-05 |
| HBA1 | -1.319277361 | 4.37E-07 |
| SOGA3 | -1.317591868 | 1.46E-18 |
| FMO5 | -1.317573682 | 7.76E-21 |
| NKG7 | -1.314639178 | 2.29E-19 |
| GBP4 | -1.312850653 | 6.00E-15 |
| IL1R2 | -1.310040311 | 9.01E-07 |
| KIR3DL2 | -1.309927214 | 3.14E-15 |
| IZUMO1 | -1.308997438 | 7.82E-14 |
| SLC9A3R2 | -1.30713878 | 7.86E-40 |
| PRKG2 | -1.305521619 | 7.28E-21 |
| P3H2 | -1.302702022 | 1.95E-30 |
| PCARE | -1.297625685 | 0.011136179 |
| NPR3 | -1.297366141 | 3.15E-25 |
| INMT | -1.293324913 | 4.23E-26 |
| ADRB2 | -1.292857464 | 5.00E-45 |
| PCDH9 | -1.292369949 | 8.34E-18 |
| LIFR | -1.285879355 | 3.31E-29 |
| TCF15 | -1.285149179 | 6.43E-17 |
| CBSL | -1.283542675 | 4.64E-06 |
| PLA2G4F | -1.283373194 | 1.76E-11 |
| LRRC36 | -1.283335834 | 1.63E-14 |
| CLIC5 | -1.282741028 | 8.83E-07 |
| HEY1 | -1.282482324 | 3.13E-20 |
| SEMA6A | -1.28230139 | 1.51E-36 |
| RXFP2 | -1.28204534 | 1.46E-08 |
| PPARGC1B | -1.28187535 | 3.12E-20 |
| ADGRE1 | -1.279036644 | 9.33E-14 |
| G6PC2 | -1.278637635 | 4.47E-10 |
| MPIG6B | -1.278304781 | 3.45E-24 |
| RND1 | -1.272804585 | 6.16E-09 |
| KL | -1.272722313 | 6.19E-15 |
| INMT-MINDY4 | -1.268710332 | 3.53E-21 |
| B3GALNT1 | -1.268502 | 2.07E-31 |
| GYPB | -1.268422291 | 2.64E-08 |
| CD5L | -1.265615476 | 8.22E-05 |
| KLRD1 | -1.264134667 | 3.44E-24 |
| RD3L | -1.25804646 | 0.001819142 |
| SYNPO2L | -1.256061251 | 2.07E-09 |
| SLCO4A1 | -1.256029285 | 2.43E-10 |
| FOSB | -1.255695217 | 4.02E-05 |
| PTCRA | -1.255525126 | 5.82E-18 |
| STC2 | -1.251690191 | 1.39E-12 |
| ART1 | -1.250813104 | 4.49E-07 |
| LRRC32 | -1.25040865 | 1.35E-19 |
| BPI | -1.246806772 | 0.000543277 |
| STX11 | -1.24620176 | 3.44E-34 |
| ID1 | -1.243780229 | 1.98E-22 |
| CLEC6A | -1.243150002 | 1.79E-08 |
| ACKR4 | -1.242023794 | 1.90E-31 |
| SLIT2 | -1.241699168 | 2.44E-29 |
| ACVRL1 | -1.239640059 | 4.83E-30 |
| ANO3 | -1.237987446 | 4.26E-09 |
| MSN | -1.23767041 | 6.22E-29 |
| CECR2 | -1.237048489 | 1.07E-31 |
| SEMA3G | -1.23667792 | 6.81E-15 |
| KCNH7 | -1.235884013 | 4.79E-10 |
| SLC17A3 | -1.234156188 | 4.11E-16 |
| AJAP1 | -1.231439572 | 1.29E-12 |
| EPAS1 | -1.228642431 | 9.82E-19 |
| S100A3 | -1.22542095 | 1.96E-19 |
| MMP8 | -1.223491084 | 0.003137958 |
| NUTM2F | -1.222815047 | 1.22E-07 |
| UPK3B | -1.221815179 | 1.28E-08 |
| SCEL | -1.22170948 | 7.62E-14 |
| MGAT3 | -1.220939332 | 9.47E-19 |
| HYAL1 | -1.220226612 | 1.80E-33 |
| ADRA1B | -1.219893217 | 1.97E-05 |
| IL18R1 | -1.218972368 | 3.60E-18 |
| CDH19 | -1.217952963 | 1.15E-28 |
| NIM1K | -1.217652961 | 5.84E-38 |
| CDO1 | -1.217144133 | 5.45E-16 |
| WFDC10B | -1.216894135 | 1.57E-14 |
| ORM1 | -1.216110096 | 5.09E-09 |
| C6orf223 | -1.2151692 | 1.05E-12 |
| FBN3 | -1.214982544 | 1.94E-09 |
| MYO7B | -1.214837745 | 1.08E-17 |
| MGAM | -1.213462214 | 5.67E-09 |
| FUT1 | -1.211666318 | 1.37E-35 |
| PRF1 | -1.208503779 | 2.99E-21 |
| AC020636.2 | -1.205657048 | 0.032164113 |
| LY6G6D | -1.20529612 | 7.00E-06 |
| CDH5 | -1.204921189 | 2.40E-27 |
| TAL1 | -1.203338098 | 1.06E-26 |
| CYS1 | -1.202510249 | 2.48E-15 |
| USHBP1 | -1.202390013 | 1.20E-26 |
| ACE | -1.200989307 | 8.02E-16 |
| RAMP2 | -1.198885019 | 3.65E-22 |
| LRFN2 | -1.196266347 | 9.04E-06 |
| CCDC141 | -1.195614903 | 7.66E-12 |
| MVB12B | -1.1950516 | 2.62E-33 |
| MRGPRX2 | -1.194978837 | 0.00069723 |
| HPCAL1 | -1.194786223 | 9.53E-52 |
| PCDH10 | -1.194751101 | 6.73E-10 |
| PRKAR2B | -1.193220716 | 0.036855544 |
| GATA3 | -1.19150774 | 1.98E-05 |
| TMEM139 | -1.191475083 | 6.35E-23 |
| LGR5 | -1.191401278 | 1.11E-08 |
| PLCXD2 | -1.189893194 | 5.06E-19 |
| RLN3 | -1.185604246 | 6.67E-06 |
| SH2D4B | -1.185508776 | 1.54E-15 |
| PDYN | -1.1826608 | 0.004490992 |
| LRRN4 | -1.181942981 | 3.29E-13 |
| GGTLC2 | -1.181786685 | 1.37E-11 |
| SULT2B1 | -1.180363312 | 2.93E-22 |
| GYPA | -1.179766694 | 1.75E-07 |
| ANKRD20A3 | -1.178820479 | 2.39E-18 |
| SEC14L6 | -1.176492691 | 8.95E-23 |
| ESAM | -1.175615449 | 7.93E-41 |
| SCN7A | -1.174182675 | 6.98E-22 |
| DISP1 | -1.173376975 | 1.73E-30 |
| CSNK1A1L | -1.172297692 | 2.74E-05 |
| FGFR4 | -1.172031814 | 2.72E-20 |
| CLEC1B | -1.170252193 | 4.15E-16 |
| LEFTY2 | -1.169874298 | 2.70E-13 |
| KANK3 | -1.169402946 | 1.32E-40 |
| NOTCH4 | -1.169109226 | 8.89E-35 |
| CLDN5 | -1.16904764 | 2.69E-19 |
| NR5A1 | -1.168529282 | 0.000492647 |
| CSF3R | -1.168397384 | 4.59E-18 |
| UGT2B4 | -1.168288805 | 3.69E-07 |
| PDZD2 | -1.168148842 | 4.50E-36 |
| RSPO2 | -1.164535938 | 3.36E-14 |
| ACOXL | -1.164138949 | 1.78E-19 |
| ITGA2B | -1.164006109 | 2.16E-13 |
| CDH10 | -1.163288446 | 6.47E-08 |
| NR1H4 | -1.161654911 | 7.95E-08 |
| VIP | -1.159117822 | 1.17E-06 |
| SLC22A8 | -1.15705 | 0.002451069 |
| IL1A | -1.156573508 | 2.43E-07 |
| TSPAN12 | -1.153989793 | 1.57E-32 |
| RGCC | -1.15314949 | 1.38E-14 |
| GPX3 | -1.150800146 | 1.32E-19 |
| AC018755.2 | -1.150690663 | 1.07E-06 |
| RIMS4 | -1.149923369 | 1.52E-17 |
| MFAP3L | -1.147476286 | 2.99E-21 |
| ZC3H12C | -1.147455771 | 5.13E-06 |
| TMEM26 | -1.143659469 | 4.47E-17 |
| EFNB2 | -1.143154466 | 1.42E-33 |
| MAP2 | -1.141904852 | 3.58E-34 |
| RIPPLY3 | -1.141481319 | 4.11E-13 |
| RAMP3 | -1.141177743 | 1.75E-18 |
| AL162171.1 | -1.138749981 | 9.73E-47 |
| RETN | -1.137386108 | 0.000327418 |
| SPX | -1.136969886 | 7.11E-15 |
| PRKACG | -1.133204726 | 0.006922082 |
| SFTA2 | -1.132310857 | 1.91E-15 |
| NOVA2 | -1.131647861 | 4.10E-21 |
| PAPSS2 | -1.1303432 | 1.57E-32 |
| LAMA3 | -1.1297749 | 1.10E-32 |
| ELAVL3 | -1.127787807 | 0.047841692 |
| MAP3K21 | -1.127251122 | 6.05E-37 |
| CD36 | -1.12534245 | 2.12E-22 |
| SPTA1 | -1.124972449 | 2.40E-07 |
| KANK4 | -1.123598751 | 1.94E-10 |
| PPARG | -1.12313871 | 2.67E-11 |
| DACH1 | -1.123054625 | 5.61E-10 |
| SMAD6 | -1.122174403 | 5.60E-10 |
| TEK | -1.121836195 | 1.08E-24 |
| THSD1 | -1.121372131 | 3.32E-23 |
| CLEC12B | -1.120283086 | 4.54E-13 |
| ZNF705A | -1.118875979 | 1.31E-05 |
| DTL | -1.118619583 | 4.32E-08 |
| PLAG1 | -1.118121969 | 4.28E-09 |
| OR2W3 | -1.118009103 | 3.25E-09 |
| FAXDC2 | -1.117876776 | 9.02E-25 |
| SEMA6D | -1.117130965 | 5.92E-22 |
| SSTR4 | -1.114980358 | 2.85E-06 |
| RAB40A | -1.113930214 | 1.52E-26 |
| ZNF541 | -1.112910933 | 3.53E-20 |
| PTPRR | -1.112226327 | 1.54E-11 |
| OR2S2 | -1.111530612 | 0.003124917 |
| LOXHD1 | -1.108195156 | 1.97E-13 |
| ABHD5 | -1.107244536 | 8.38E-11 |
| KLF4 | -1.107164077 | 8.86E-15 |
| CASP5 | -1.106752807 | 8.48E-13 |
| RCSD1 | -1.103669842 | 1.31E-23 |
| LRRC19 | -1.102485351 | 4.28E-12 |
| GFI1B | -1.101954557 | 1.92E-19 |
| CCBE1 | -1.100979817 | 9.99E-14 |
| TSPAN16 | -1.100119639 | 3.13E-08 |
| GPRIN3 | -1.0969136 | 0.001513928 |
| RGR | -1.096367317 | 8.61E-06 |
| MPP3 | -1.091243273 | 1.98E-18 |
| NTRK2 | -1.091185229 | 1.89E-10 |
| ADGRL2 | -1.090570616 | 5.72E-28 |
| CACNA2D3 | -1.090209747 | 1.33E-26 |
| GABRB2 | -1.090035898 | 9.26E-15 |
| HPGD | -1.08926192 | 1.22E-11 |
| PIGA | -1.085424926 | 2.67E-17 |
| HIGD1B | -1.084412315 | 3.32E-16 |
| SLC22A10 | -1.082209056 | 5.89E-09 |
| C19orf81 | -1.082082265 | 3.40E-12 |
| ICAM1 | -1.080471912 | 2.87E-09 |
| SHH | -1.079785656 | 1.45E-11 |
| FGL1 | -1.077617269 | 2.80E-06 |
| CLDN24 | -1.07729165 | 3.57E-09 |
| KCNK16 | -1.073603933 | 1.58E-05 |
| NLGN4X | -1.073527337 | 4.71E-11 |
| PIP5K1B | -1.072702279 | 1.14E-39 |
| EXOC3L1 | -1.070822213 | 5.54E-24 |
| C1orf198 | -1.067897972 | 1.57E-47 |
| NEBL | -1.067167946 | 1.25E-38 |
| CCM2L | -1.06610495 | 6.15E-23 |
| ZNF358 | -1.065033625 | 1.24E-06 |
| CLEC14A | -1.064857866 | 8.48E-24 |
| CLDN18 | -1.063874744 | 1.08E-06 |
| KCNS3 | -1.06300896 | 5.04E-13 |
| SLC2A12 | -1.061637889 | 1.01E-19 |
| LIMCH1 | -1.061038694 | 4.15E-36 |
| MYBPC1 | -1.060860455 | 7.97E-06 |
| PADI4 | -1.060822974 | 6.12E-06 |
| LPL | -1.059925017 | 1.86E-07 |
| BATF2 | -1.059852059 | 2.32E-11 |
| PRSS21 | -1.058959621 | 1.66E-07 |
| DIPK1B | -1.057047264 | 2.75E-17 |
| LMO7 | -1.056973432 | 1.57E-24 |
| JPH1 | -1.05665841 | 4.01E-07 |
| DLC1 | -1.056255595 | 4.39E-15 |
| TMIGD2 | -1.056076826 | 5.38E-17 |
| LINGO2 | -1.054102807 | 3.04E-07 |
| JUN | -1.05377076 | 4.19E-15 |
| AC068775.1 | -1.053546409 | 9.37E-07 |
| CPAMD8 | -1.052726668 | 3.72E-29 |
| RSPO1 | -1.051023408 | 5.41E-15 |
| GGT1 | -1.050252232 | 2.29E-22 |
| TBX3 | -1.050008751 | 5.55E-15 |
| PCDH11X | -1.049499611 | 4.59E-11 |
| FFAR4 | -1.049439941 | 8.88E-09 |
| EMP2 | -1.049095123 | 1.70E-17 |
| C1orf167 | -1.049072282 | 6.57E-17 |
| ADRA1D | -1.04900741 | 7.91E-14 |
| SMIM9 | -1.048517188 | 0.002628406 |
| ST6GALNAC5 | -1.047633934 | 3.91E-18 |
| FIBCD1 | -1.046076017 | 5.10E-12 |
| SLC46A2 | -1.044751887 | 1.82E-07 |
| CHRM4 | -1.044482861 | 1.37E-06 |
| TOX2 | -1.044399381 | 2.05E-23 |
| PGC | -1.044190711 | 1.80E-05 |
| DENND3 | -1.042664473 | 3.04E-33 |
| FRMD1 | -1.04207399 | 1.29E-06 |
| MAP1LC3C | -1.041881698 | 9.35E-11 |
| ARHGAP6 | -1.041337022 | 4.97E-43 |
| DLL4 | -1.040105833 | 4.71E-19 |
| GPBAR1 | -1.039614145 | 3.87E-21 |
| DUSP8 | -1.039007719 | 7.08E-13 |
| MAOA | -1.038619537 | 3.96E-31 |
| KCNIP1 | -1.037595069 | 1.89E-13 |
| TEF | -1.036123025 | 1.13E-11 |
| HBB | -1.034139275 | 6.19E-05 |
| GYPE | -1.033650346 | 6.96E-05 |
| AC008537.1 | -1.032780757 | 0.043276149 |
| CCL26 | -1.031715203 | 3.60E-07 |
| NTNG1 | -1.03148859 | 3.55E-16 |
| TAS2R60 | -1.031024252 | 0.001507663 |
| TBX20 | -1.029687329 | 0.00067184 |
| TBX21 | -1.029409734 | 2.91E-06 |
| Z82206.1 | -1.029326978 | 1.26E-11 |
| SMIM41 | -1.029320076 | 6.67E-19 |
| ADIRF | -1.028949849 | 1.46E-29 |
| SIRPB1 | -1.027651934 | 1.07E-10 |
| HIST1H2BO | -1.02721452 | 0.005372154 |
| PRKCE | -1.027088188 | 5.35E-15 |
| IFIT1B | -1.027005078 | 4.42E-05 |
| GGTLC3 | -1.026880108 | 7.82E-07 |
| CHI3L2 | -1.025816968 | 3.42E-09 |
| GPR17 | -1.024896024 | 4.34E-09 |
| SULT1C4 | -1.024509238 | 4.52E-24 |
| P2RY14 | -1.024125087 | 1.05E-12 |
| SPRYD7 | -1.023928053 | 2.64E-25 |
| CD302 | -1.022878988 | 1.31E-16 |
| LIN7A | -1.021895354 | 1.61E-38 |
| METTL21C | -1.021474547 | 0.000156491 |
| CD244 | -1.020796047 | 1.35E-07 |
| MMEL1 | -1.020459965 | 3.50E-17 |
| IL18RAP | -1.017645815 | 3.29E-10 |
| STC1 | -1.017165767 | 0.010194949 |
| AL132671.2 | -1.016328245 | 2.37E-05 |
| TPPP2 | -1.015938651 | 5.65E-08 |
| ACRBP | -1.01563953 | 1.50E-16 |
| C5orf38 | -1.014538055 | 4.10E-20 |
| MEFV | -1.012980448 | 8.91E-12 |
| CLEC4E | -1.012565499 | 9.86E-10 |
| CRYAB | -1.012294659 | 4.24E-10 |
| SLC17A1 | -1.012056569 | 0.001210987 |
| OR6K3 | -1.011960279 | 1.21E-10 |
| SMCO3 | -1.011394681 | 2.36E-13 |
| ODF3L2 | -1.0091082 | 1.46E-16 |
| MYH2 | -1.008638192 | 0.000175759 |
| KRT85 | -1.006627805 | 2.17E-05 |
| IL7R | -1.002454231 | 1.98E-11 |
| PTGS2 | -1.001019757 | 0.000191026 |
| ADH1B | -0.999823342 | 1.04E-16 |
| GATA2 | -0.998405031 | 2.34E-18 |
| RAPGEF5 | -0.998230859 | 5.23E-22 |
| PPP4R4 | -0.99742802 | 2.38E-08 |
| SYT15 | -0.996622491 | 6.16E-28 |
| HIST1H2AD | -0.996530487 | 0.04605143 |
| NPR1 | -0.99511734 | 1.38E-14 |
| USP44 | -0.995060086 | 1.10E-11 |
| TCF21 | -0.994364711 | 1.43E-22 |
| OCLN | -0.992129178 | 8.46E-34 |
| VSIG10 | -0.991777747 | 9.74E-37 |
| LIMS2 | -0.986871256 | 6.11E-21 |
| MXD1 | -0.985917239 | 7.80E-07 |
| RAD21L1 | -0.985062385 | 0.002726369 |
| PTF1A | -0.984289178 | 0.046727158 |
| PKNOX2 | -0.983553928 | 2.56E-20 |
| CHIA | -0.983383588 | 1.23E-05 |
| TPRG1 | -0.981891312 | 2.89E-40 |
| SULT1A2 | -0.980069488 | 8.85E-22 |
| TRPM1 | -0.979212498 | 0.000643742 |
| TCAP | -0.978358385 | 5.91E-14 |
| CDKL2 | -0.977487207 | 3.36E-17 |
| NINJ2 | -0.975927822 | 2.65E-28 |
| CEACAM4 | -0.975656613 | 2.04E-09 |
| ARRB1 | -0.975527388 | 2.60E-56 |
| ROBO4 | -0.974728224 | 1.11E-18 |
| SIGLEC5 | -0.973419567 | 1.57E-08 |
| ZNF560 | -0.972406164 | 0.00015451 |
| HOPX | -0.972185791 | 5.40E-15 |
| REG4 | -0.970709958 | 0.007543176 |
| ABCA3 | -0.967722741 | 2.81E-08 |
| EGFL7 | -0.963997227 | 1.13E-21 |
| RNF144B | -0.963404845 | 6.71E-45 |
| CRKL | -0.962583615 | 0.007991987 |
| SDR16C5 | -0.961579814 | 7.30E-11 |
| LAMB4 | -0.958712423 | 1.42E-09 |
| NRGN | -0.95835456 | 2.69E-19 |
| QRICH2 | -0.957311422 | 1.04E-19 |
| CALN1 | -0.957256092 | 0.000124903 |
| SPTBN1 | -0.956951301 | 1.61E-20 |
| RGS6 | -0.956941369 | 3.13E-14 |
| PTPN5 | -0.956225051 | 5.75E-07 |
| CIB4 | -0.954719918 | 0.003450429 |
| RADIL | -0.953894216 | 1.45E-28 |
| SFTPA1 | -0.951103906 | 5.06E-08 |
| LILRA2 | -0.950813176 | 8.36E-15 |
| CCL4 | -0.950605089 | 2.48E-07 |
| FRY | -0.949928873 | 1.14E-33 |
| MGAT5B | -0.949887328 | 3.18E-13 |
| KCNH6 | -0.948540586 | 2.75E-10 |
| DUXA | -0.94819977 | 2.81E-07 |
| DSCAM | -0.947240714 | 0.000703388 |
| HIST2H2AA3 | -0.945630328 | 0.025274503 |
| HYAL2 | -0.94416172 | 1.98E-18 |
| IFI27 | -0.943373568 | 3.86E-20 |
| SYN3 | -0.942651429 | 1.39E-22 |
| MANSC4 | -0.941755699 | 6.34E-09 |
| SYDE2 | -0.936971307 | 2.95E-12 |
| HAL | -0.936970719 | 3.84E-09 |
| KCNG3 | -0.936724483 | 0.000175714 |
| GLT1D1 | -0.935994807 | 9.68E-07 |
| AGTR1 | -0.933691866 | 1.40E-17 |
| INHBA | -0.932050576 | 7.48E-05 |
| EXD1 | -0.931344959 | 3.66E-07 |
| MYOZ1 | -0.929971142 | 1.61E-13 |
| ANKRD62 | -0.929170096 | 3.06E-08 |
| ARHGEF10 | -0.928954105 | 1.35E-29 |
| MCEMP1 | -0.928130187 | 1.90E-07 |
| OLIG1 | -0.926712554 | 2.11E-05 |
| FAM189A2 | -0.925923308 | 6.47E-08 |
| SIRPD | -0.924863169 | 4.23E-07 |
| CAMK1G | -0.924554709 | 0.034428998 |
| B3GALT2 | -0.92058622 | 1.62E-10 |
| B3GNT8 | -0.920443663 | 4.44E-17 |
| TMEM130 | -0.919521982 | 1.20E-09 |
| GPD1 | -0.918752642 | 8.43E-07 |
| CLDN34 | -0.918593054 | 1.69E-06 |
| LHFPL3 | -0.918004466 | 0.014493763 |
| LGALSL | -0.917762132 | 1.46E-17 |
| MMP25 | -0.91686821 | 2.54E-06 |
| CLEC1A | -0.914151096 | 2.46E-22 |
| RASIP1 | -0.9130574 | 1.76E-16 |
| OASL | -0.912417272 | 3.01E-09 |
| CGNL1 | -0.912156103 | 1.10E-29 |
| ADAMTS7 | -0.911787418 | 5.77E-11 |
| UNC13B | -0.910986841 | 1.94E-32 |
| REP15 | -0.910729775 | 1.16E-19 |
| GIMAP6 | -0.910220605 | 6.10E-23 |
| AZU1 | -0.908642647 | 6.75E-10 |
| MT1A | -0.908297405 | 0.004195649 |
| PPFIBP1 | -0.907326549 | 1.45E-29 |
| SLAIN1 | -0.906188194 | 1.09E-39 |
| ATF3 | -0.904921202 | 4.41E-05 |
| VGLL3 | -0.904189978 | 4.50E-08 |
| GRK5 | -0.902859101 | 7.35E-32 |
| GRASP | -0.902612747 | 3.59E-14 |
| ANP32D | -0.902447706 | 0.018102541 |
| SFTPA2 | -0.901659037 | 3.40E-07 |
| KLRC2 | -0.900368156 | 8.74E-07 |
| KLF17 | -0.898668643 | 1.00E-13 |
| NPNT | -0.897981071 | 7.33E-22 |
| RGS18 | -0.895643357 | 1.04E-07 |
| OVCH1 | -0.895614281 | 8.70E-06 |
| PADI6 | -0.89486509 | 0.005078385 |
| CLEC9A | -0.894514978 | 8.42E-08 |
| SULT1A1 | -0.894465532 | 5.35E-18 |
| AL136531.2 | -0.893265941 | 5.97E-11 |
| RAB11FIP1 | -0.892644293 | 3.86E-18 |
| KIR2DL4 | -0.892138669 | 6.47E-09 |
| KCNK17 | -0.890233338 | 5.98E-07 |
| DAO | -0.889074606 | 0.000328452 |
| AC124312.1 | -0.888571777 | 3.12E-15 |
| SHISA2 | -0.886712176 | 1.47E-09 |
| KLRB1 | -0.886489519 | 4.54E-12 |
| CDKN2B | -0.886259016 | 9.15E-10 |
| IP6K3 | -0.885962283 | 1.24E-05 |
| MYO1A | -0.885926644 | 6.78E-06 |
| CXCR2 | -0.88592049 | 0.002846258 |
| RANBP3L | -0.885636806 | 5.53E-09 |
| S1PR1 | -0.884845129 | 1.50E-17 |
| ADGRF5 | -0.882602288 | 1.50E-23 |
| HSD3B1 | -0.882265358 | 2.02E-08 |
| OLR1 | -0.880679338 | 1.76E-05 |
| SUSD2 | -0.880520119 | 1.05E-06 |
| TIMP3 | -0.87928509 | 4.40E-18 |
| ATP13A4 | -0.878176781 | 6.42E-11 |
| PADI2 | -0.877133276 | 1.95E-06 |
| GALNT18 | -0.876058536 | 1.66E-06 |
| GRPEL2 | -0.875254239 | 1.21E-09 |
| CSNK2A3 | -0.874885973 | 9.70E-08 |
| TTN | -0.874372888 | 2.09E-13 |
| LURAP1L | -0.873559248 | 7.47E-12 |
| KRT79 | -0.873443864 | 0.00023147 |
| ECHDC3 | -0.873384864 | 7.35E-12 |
| APOL3 | -0.87212206 | 5.76E-24 |
| SPRY4 | -0.872038646 | 7.47E-14 |
| SPAAR | -0.871047107 | 1.37E-23 |
| ANKRD33 | -0.869863859 | 0.000287544 |
| OR8H2 | -0.869676396 | 0.019059137 |
| ADGRD1 | -0.869523112 | 2.01E-22 |
| HMGCS1 | -0.868573466 | 1.26E-23 |
| ACSM5 | -0.867230655 | 1.18E-11 |
| MOGAT1 | -0.867227253 | 0.000189547 |
| GPM6B | -0.867086457 | 1.45E-26 |
| ATP8A1 | -0.866340724 | 1.18E-23 |
| C9orf153 | -0.865764744 | 4.80E-09 |
| NLRP12 | -0.864533575 | 2.29E-10 |
| HLA-E | -0.862908223 | 2.70E-17 |
| SFTPC | -0.86220732 | 3.02E-05 |
| PAK4 | -0.86048088 | 1.33E-30 |
| SFTPD | -0.860328586 | 9.20E-08 |
| CFAP161 | -0.858978077 | 8.94E-07 |
| DESI2 | -0.858849669 | 1.98E-11 |
| FCRL6 | -0.858285221 | 8.10E-11 |
| CTSW | -0.85753121 | 2.23E-09 |
| AL049634.2 | -0.857365089 | 1.76E-05 |
| SLC5A4 | -0.856824581 | 1.76E-12 |
| ACR | -0.856561733 | 6.11E-10 |
| AKAP2 | -0.856131059 | 5.38E-17 |
| LRP2 | -0.855551546 | 9.41E-05 |
| CORO6 | -0.855115193 | 1.43E-11 |
| MCHR2 | -0.854284699 | 0.002386501 |
| GPER1 | -0.853975245 | 8.66E-09 |
| HES7 | -0.853554406 | 3.18E-05 |
| C20orf202 | -0.853387839 | 6.10E-12 |
| FAM187B | -0.853197042 | 2.52E-05 |
| APOH | -0.853106419 | 0.000383674 |
| ZP2 | -0.852670321 | 6.02E-08 |
| RAB17 | -0.852341743 | 1.08E-15 |
| SH3RF1 | -0.852040438 | 0.004842107 |
| RGS9 | -0.850842326 | 2.44E-23 |
| LRRK2 | -0.849061515 | 1.09E-08 |
| UBASH3B | -0.848819129 | 4.07E-07 |
| KIAA0040 | -0.847995834 | 2.43E-18 |
| EDA | -0.847330882 | 4.87E-26 |
| KHDRBS3 | -0.845762563 | 1.73E-23 |
| TRHDE | -0.845367093 | 9.71E-14 |
| KLRC4 | -0.84392912 | 1.41E-06 |
| SLC22A24 | -0.842863494 | 0.007389953 |
| AMOTL2 | -0.842831886 | 8.90E-25 |
| FRMD3 | -0.842643995 | 5.60E-17 |
| CASS4 | -0.842338887 | 1.43E-14 |
| FCGR3B | -0.841185229 | 5.72E-05 |
| MAP4K2 | -0.840199864 | 9.19E-29 |
| SELENOP | -0.839338141 | 1.39E-07 |
| FOS | -0.838753626 | 6.18E-05 |
| SLC15A5 | -0.838421905 | 0.003716763 |
| CCNB3 | -0.838234319 | 2.93E-11 |
| CITED2 | -0.838209009 | 6.54E-19 |
| ZNF534 | -0.838011474 | 1.77E-05 |
| OTUD1 | -0.837432557 | 1.98E-28 |
| HBEGF | -0.836456484 | 1.35E-05 |
| AOC3 | -0.835460179 | 1.11E-12 |
| NPB | -0.83518018 | 0.000139418 |
| FLT4 | -0.834937227 | 1.68E-14 |
| CELA2B | -0.834928383 | 4.64E-07 |
| HIP1 | -0.834679114 | 5.91E-18 |
| SORL1 | -0.834277684 | 2.57E-06 |
| CBS | -0.834273955 | 3.96E-09 |
| KIF17 | -0.834225373 | 3.96E-27 |
| RAB8B | -0.83324413 | 1.73E-06 |
| FZD8 | -0.832881355 | 5.42E-22 |
| FMO2 | -0.832173304 | 8.42E-07 |
| PDGFB | -0.831423752 | 6.43E-13 |
| GJB1 | -0.831049415 | 9.51E-06 |
| SLC30A8 | -0.830533321 | 0.000652053 |
| MAB21L2 | -0.830397777 | 2.30E-05 |
| FKBP1B | -0.828499222 | 5.39E-27 |
| CXCL3 | -0.827724341 | 3.45E-05 |
| MPL | -0.827583656 | 7.85E-13 |
| GPC3 | -0.82714856 | 2.74E-16 |
| TMEM97 | -0.826345823 | 7.11E-06 |
| CALCRL | -0.826131363 | 1.78E-11 |
| MAPT | -0.825792612 | 4.18E-08 |
| FGD5 | -0.82576383 | 1.50E-25 |
| BDNF | -0.825686081 | 9.91E-08 |
| GZMH | -0.824456172 | 3.15E-06 |
| ANKRD20A2 | -0.824292981 | 1.55E-06 |
| SIGLEC10 | -0.824164605 | 9.63E-08 |
| STARD8 | -0.823391091 | 1.68E-26 |
| LRRTM4 | -0.822732849 | 1.28E-08 |
| C11orf21 | -0.822478242 | 9.82E-12 |
| STARD9 | -0.82246191 | 2.92E-19 |
| HSPA12B | -0.822430305 | 0.005116957 |
| LILRA5 | -0.821812582 | 5.73E-07 |
| SLITRK2 | -0.821651497 | 4.31E-17 |
| CD274 | -0.821513329 | 5.58E-08 |
| PRRT1B | -0.819487287 | 1.95E-08 |
| SLC25A37 | -0.81935747 | 1.91E-09 |
| HNF1B | -0.8189364 | 5.43E-11 |
| GAB1 | -0.818110646 | 6.22E-28 |
| DOCK9 | -0.816614378 | 1.61E-26 |
| DCC | -0.815916738 | 1.25E-07 |
| OR2T10 | -0.815890932 | 0.00058333 |
| HMSD | -0.815738948 | 1.72E-05 |
| ATOH8 | -0.814954307 | 7.29E-15 |
| CLEC12A | -0.813808567 | 4.78E-09 |
| AGTPBP1 | -0.813370613 | 9.34E-62 |
| SNX25 | -0.812024234 | 1.21E-10 |
| TBC1D28 | -0.810497378 | 0.000104304 |
| NFKBIA | -0.810219333 | 7.00E-12 |
| CD160 | -0.809863675 | 3.26E-13 |
| CXCL2 | -0.80937509 | 9.59E-06 |
| SYT1 | -0.80861312 | 1.11E-06 |
| COL6A6 | -0.80845153 | 6.35E-08 |
| RAPGEF4 | -0.808042892 | 7.39E-22 |
| ICAM2 | -0.80798203 | 1.86E-20 |
| NTNG2 | -0.806977619 | 3.12E-10 |
| FAM72A | -0.806605634 | 1.04E-14 |
| LYPD6 | -0.805209854 | 0.001535973 |
| TJP1 | -0.804666981 | 1.17E-45 |
| AFAP1L1 | -0.804384349 | 2.77E-10 |
| LY6G6C | -0.801904492 | 0.013458242 |
| PITPNM2 | -0.801202565 | 2.13E-31 |
| GUCY1A2 | -0.801122938 | 8.35E-10 |
| PARD6B | -0.80103531 | 0.003979356 |
| ADORA2A | -0.799150329 | 6.92E-16 |
| SCAI | -0.797214415 | 1.38E-30 |
| HSPA1A | -0.797089525 | 0.000112437 |
| SLC40A1 | -0.796720058 | 5.31E-16 |
| DMRT2 | -0.796607554 | 1.96E-06 |
| ST6GALNAC3 | -0.795580453 | 1.27E-08 |
| SPDYC | -0.79543572 | 0.003022503 |
| TSPAN18 | -0.79410283 | 3.21E-15 |
| HPN | -0.792306778 | 7.19E-08 |
| KLF2 | -0.792047691 | 1.36E-10 |
| KLF10 | -0.792047503 | 1.59E-09 |
| VSIG2 | -0.792044767 | 5.38E-19 |
| GPR146 | -0.790689592 | 8.47E-12 |
| RBMS2 | -0.790564725 | 2.87E-09 |
| MID1IP1 | -0.79010751 | 3.76E-23 |
| TIE1 | -0.788874738 | 2.83E-15 |
| CD101 | -0.78863815 | 1.25E-07 |
| SIGLEC11 | -0.787231964 | 2.32E-09 |
| PDGFRA | -0.787107169 | 3.32E-07 |
| SMAD7 | -0.786596594 | 1.06E-24 |
| C1orf210 | -0.784728783 | 1.98E-13 |
| IRX2 | -0.784718803 | 9.96E-14 |
| DOCK4 | -0.784250551 | 2.19E-32 |
| FAM89A | -0.783640494 | 3.00E-08 |
| ADM | -0.783628658 | 9.92E-05 |
| BEX5 | -0.783560838 | 0.001794559 |
| COX4I2 | -0.782765427 | 8.25E-13 |
| HIST1H3H | -0.782451982 | 6.83E-10 |
| RXRG | -0.782219134 | 1.86E-05 |
| TMEM52B | -0.781350829 | 4.17E-06 |
| IFITM5 | -0.781240168 | 0.003330069 |
| AGO1 | -0.780724122 | 0.002857869 |
| ASRGL1 | -0.779571646 | 3.40E-16 |
| CFAP20 | -0.779457009 | 3.39E-16 |
| ARAP3 | -0.779394841 | 1.48E-17 |
| CCRL2 | -0.777644697 | 2.05E-11 |
| ASB11 | -0.777125617 | 8.56E-08 |
| BEND2 | -0.775801614 | 2.13E-06 |
| PRLR | -0.775650847 | 1.03E-06 |
| AC019257.8 | -0.775425406 | 0.002311336 |
| GIMAP4 | -0.774597894 | 4.20E-09 |
| ARHGAP31 | -0.774428228 | 7.94E-26 |
| KCNN2 | -0.774415547 | 1.89E-05 |
| TNNT2 | -0.773700251 | 5.97E-11 |
| ANKRD34C | -0.772441239 | 1.48E-06 |
| ZBTB16 | -0.772383439 | 0.000231945 |
| NOSTRIN | -0.771935074 | 3.31E-16 |
| CMPK2 | -0.77140657 | 4.66E-16 |
| ESYT3 | -0.770413252 | 2.17E-07 |
| LRP4 | -0.769808117 | 4.05E-10 |
| NFE4 | -0.768669881 | 0.00031345 |
| JDP2 | -0.768461587 | 2.41E-18 |
| MSMO1 | -0.767353654 | 3.87E-12 |
| F8 | -0.76619003 | 1.63E-10 |
| UNC13D | -0.76606535 | 4.75E-25 |
| KLRC1 | -0.765633305 | 6.62E-11 |
| BNIP3L | -0.764869725 | 2.31E-07 |
| KIF12 | -0.764591184 | 9.94E-08 |
| BFSP1 | -0.764345734 | 3.85E-20 |
| FXYD6 | -0.762791676 | 2.85E-18 |
| KLF15 | -0.762190319 | 0.002394523 |
| PRAM1 | -0.761782591 | 8.08E-10 |
| ADGRE3 | -0.761094392 | 2.87E-05 |
| TULP2 | -0.760418112 | 3.51E-05 |
| TMEM255A | -0.760305159 | 1.85E-08 |
| SIGLECL1 | -0.759207954 | 0.01928437 |
| TLR8 | -0.75828536 | 3.93E-07 |
| ADCYAP1R1 | -0.757819707 | 1.77E-05 |
| ZNF511-PRAP1 | -0.75775952 | 3.19E-06 |
| GADD45B | -0.756356769 | 5.39E-06 |
| TGFBR3 | -0.755458785 | 2.99E-11 |
| SHROOM4 | -0.75519597 | 5.19E-18 |
| DPEP2 | -0.755156077 | 2.47E-11 |
| ANKRD44 | -0.755143639 | 1.31E-07 |
| SFTA3 | -0.754856299 | 8.46E-10 |
| ANKRD33B | -0.754761568 | 0.012983489 |
| FGF12 | -0.754283072 | 1.43E-09 |
| SLC24A4 | -0.754072697 | 1.42E-11 |
| SHE | -0.754060629 | 3.09E-15 |
| ZNF683 | -0.753223034 | 0.000482782 |
| GPD1L | -0.752858653 | 2.35E-32 |
| KLF6 | -0.752775387 | 2.78E-05 |
| SEMA3B | -0.752548078 | 2.26E-18 |
| BCL6B | -0.749893899 | 5.60E-11 |
| RXFP4 | -0.748887736 | 6.65E-05 |
| PRKCZ | -0.748732401 | 2.63E-17 |
| NOTUM | -0.748619324 | 0.027646488 |
| PPP2R5A | -0.748002373 | 4.99E-10 |
| IGFALS | -0.747498353 | 1.09E-09 |
| VEPH1 | -0.746909544 | 6.60E-07 |
| PKDCC | -0.746378961 | 1.07E-13 |
| ETV1 | -0.746294423 | 2.90E-15 |
| PTGDR | -0.746043253 | 5.89E-09 |
| MRPL54 | -0.745461096 | 3.33E-10 |
| CMTM5 | -0.74541795 | 1.72E-09 |
| PCDH15 | -0.74521275 | 1.44E-07 |
| APOA2 | -0.743737761 | 0.004237585 |
| PQLC2L | -0.74346835 | 5.41E-08 |
| CDKN2D | -0.743303511 | 1.67E-15 |
| SLC12A1 | -0.742421164 | 0.011017553 |
| MYLIP | -0.74150704 | 1.02E-28 |
| REPS2 | -0.741117421 | 4.38E-26 |
| ZNF365 | -0.740122715 | 1.02E-05 |
| RRAS | -0.739943838 | 3.51E-14 |
| PRSS48 | -0.739585383 | 0.001465802 |
| GIMAP5 | -0.739206711 | 6.81E-16 |
| FXYD6-FXYD2 | -0.739007747 | 0.003277745 |
| LDB2 | -0.73830563 | 6.38E-17 |
| USP53 | -0.736158746 | 1.36E-16 |
| ZNF704 | -0.735831321 | 2.86E-19 |
| CCDC63 | -0.735774477 | 0.00712442 |
| FLRT3 | -0.735285639 | 2.74E-08 |
| PRTG | -0.734564309 | 1.70E-06 |
| PAG1 | -0.733256877 | 1.67E-15 |
| CTTNBP2NL | -0.732105999 | 2.00E-10 |
| BMPR2 | -0.731323509 | 2.58E-15 |
| SH3BP5 | -0.731126555 | 1.88E-08 |
| CCN3 | -0.730832226 | 0.035420324 |
| AC142391.1 | -0.730388789 | 0.008763863 |
| BST2 | -0.7298503 | 4.57E-07 |
| ITGA10 | -0.729784872 | 5.78E-08 |
| WFIKKN2 | -0.72966094 | 0.000188748 |
| RAPH1 | -0.729591376 | 1.45E-05 |
| TFPI | -0.729460635 | 2.95E-22 |
| ADAMTSL3 | -0.728559542 | 5.30E-15 |
| ZNF467 | -0.728121065 | 2.39E-18 |
| SELENBP1 | -0.727850222 | 1.98E-09 |
| PTN | -0.72765685 | 6.31E-09 |
| PNPLA3 | -0.727571523 | 3.99E-05 |
| PTPRM | -0.727475327 | 1.32E-33 |
| LRRC31 | -0.727439412 | 4.46E-05 |
| CTRB1 | -0.727410848 | 0.006495608 |
| MOCS1 | -0.727198716 | 1.03E-13 |
| SEL1L2 | -0.72655327 | 0.001668513 |
| SPECC1L-ADORA2A | -0.725583127 | 2.91E-05 |
| LONRF3 | -0.724010815 | 1.37E-09 |
| KRT71 | -0.723947876 | 0.040573306 |
| ARAP2 | -0.723603855 | 1.10E-11 |
| IQCN | -0.723552489 | 7.24E-07 |
| FOXA2 | -0.723552215 | 2.91E-11 |
| HSD3B2 | -0.722850754 | 2.04E-07 |
| PAQR5 | -0.72276495 | 1.08E-14 |
| RORC | -0.721797744 | 4.81E-08 |
| RTN1 | -0.721421991 | 3.50E-14 |
| PNMA6E | -0.721068656 | 0.011037428 |
| FGG | -0.720987353 | 0.034906616 |
| TMCO2 | -0.72054484 | 0.007969241 |
| MFSD2A | -0.719389474 | 3.67E-07 |
| EPDR1 | -0.718604634 | 4.80E-07 |
| TNFSF10 | -0.717528231 | 1.87E-08 |
| DKK2 | -0.715576271 | 0.021682628 |
| NEDD4L | -0.714866928 | 1.53E-16 |
| TNS1 | -0.714431703 | 2.75E-31 |
| GDF2 | -0.714089537 | 0.008735918 |
| CPNE2 | -0.713974026 | 5.83E-29 |
| QKI | -0.713424628 | 5.23E-22 |
| PROSER2 | -0.713333064 | 7.54E-17 |
| AC113554.1 | -0.713308055 | 0.000296571 |
| RIPOR2 | -0.713222507 | 5.52E-09 |
| TNNI1 | -0.713181127 | 3.92E-07 |
| ANKRD20A4 | -0.712714418 | 8.66E-09 |
| CTAGE15 | -0.712678571 | 0.002766837 |
| EPB41L2 | -0.712187204 | 1.22E-19 |
| IFIT2 | -0.711929525 | 1.04E-09 |
| SMLR1 | -0.711327564 | 0.001708253 |
| NTSR1 | -0.711166123 | 0.001411307 |
| ASGR2 | -0.710870085 | 1.02E-05 |
| FBXL16 | -0.710819508 | 9.34E-09 |
| SAMD14 | -0.710487958 | 8.42E-08 |
| PARP14 | -0.709546538 | 7.39E-09 |
| GAB2 | -0.709126702 | 2.35E-10 |
| CD300LF | -0.708599585 | 9.06E-10 |
| WSCD1 | -0.708431441 | 3.63E-07 |
| DUSP26 | -0.708346482 | 0.015517651 |
| FGR | -0.708203024 | 1.17E-11 |
| VSIR | -0.707384657 | 4.17E-27 |
| IFIT3 | -0.706844724 | 5.10E-08 |
| PROK2 | -0.706651461 | 0.010317534 |
| AHNAK | -0.704877299 | 4.53E-16 |
| BCAS2 | -0.704552972 | 3.45E-06 |
| GNG11 | -0.703710518 | 3.06E-14 |
| FBLN5 | -0.703230511 | 3.09E-15 |
| OR2A1 | -0.702986317 | 2.47E-06 |
| LGI3 | -0.702279014 | 0.012374417 |
| CABLES1 | -0.702131854 | 7.82E-17 |
| LMO2 | -0.701701042 | 1.47E-19 |
| CXCR1 | -0.701220981 | 0.004225308 |
| MAPK4 | -0.701064332 | 0.009442627 |
| IDO1 | -0.701018835 | 0.001032353 |
| POU6F2 | -0.700365515 | 0.000289704 |
| TSPAN7 | -0.699967073 | 7.69E-16 |
| EPOR | -0.699358408 | 6.60E-13 |
| MMP28 | -0.699107932 | 2.49E-12 |
| B3GNT7 | -0.697859359 | 0.00115601 |
| FREM3 | -0.697145591 | 0.001810165 |
| MACF1 | -0.697104947 | 5.59E-30 |
| LDHD | -0.696843818 | 2.30E-16 |
| TREML4 | -0.696670901 | 0.001466076 |
| SRGAP2C | -0.696414412 | 3.21E-26 |
| CHPT1 | -0.696327342 | 4.51E-31 |
| TJP2 | -0.69595375 | 3.59E-18 |
| PDE6A | -0.695432248 | 1.21E-09 |
| SLC27A3 | -0.695311674 | 2.51E-12 |
| ZFYVE9 | -0.694993538 | 7.12E-33 |
| PLEKHA1 | -0.694873614 | 2.60E-30 |
| NRN1 | -0.694496528 | 2.14E-13 |
| DOCK6 | -0.693770509 | 4.43E-15 |
| CLIC2 | -0.693697215 | 9.42E-09 |
| FASN | -0.693600789 | 1.88E-07 |
| PCDHA12 | -0.693436249 | 0.004191339 |
| CHP1 | -0.693431341 | 6.96E-11 |
| OSGIN2 | -0.69320159 | 5.88E-16 |
| CD247 | -0.693184098 | 4.20E-09 |
| MAL2 | -0.692810254 | 0.0011226 |
| PDE4C | -0.692809587 | 5.08E-07 |
| TAS2R50 | -0.692613453 | 0.002300709 |
| HIST2H2AC | -0.691564439 | 6.08E-06 |
| SP5 | -0.691080691 | 2.50E-08 |
| CYREN | -0.690807156 | 3.69E-14 |
| RASSF8 | -0.690750746 | 1.34E-17 |
| PHACTR1 | -0.690573422 | 2.24E-15 |
| CARD16 | -0.690203444 | 4.74E-13 |
| PDE1C | -0.689741338 | 7.51E-13 |
| PTPRG | -0.686545243 | 2.06E-06 |
| SLC27A6 | -0.686455461 | 0.000140487 |
| LMAN1L | -0.686360568 | 0.002735623 |
| PREX1 | -0.68611846 | 1.65E-13 |
| HEG1 | -0.684539589 | 8.53E-07 |
| PEAR1 | -0.684194678 | 1.29E-14 |
| KCTD19 | -0.683653674 | 1.87E-08 |
| CXCL11 | -0.683562583 | 0.004752511 |
| SSH1 | -0.683093682 | 5.27E-05 |
| CYP51A1 | -0.682922308 | 2.52E-06 |
| AOC2 | -0.682908975 | 7.91E-05 |
| STARD3 | -0.682330255 | 6.59E-19 |
| CAV2 | -0.682268734 | 1.52E-19 |
| DNAH17 | -0.681344857 | 1.21E-08 |
| EVA1A | -0.681276093 | 2.50E-10 |
| CX3CL1 | -0.680761387 | 0.000249709 |
| IMPG1 | -0.680738486 | 3.63E-06 |
| SNX18 | -0.680176421 | 6.41E-07 |
| AC007326.4 | -0.680155787 | 4.00E-05 |
| GRPR | -0.679761095 | 1.79E-07 |
| PRKCQ | -0.679124304 | 3.36E-13 |
| ITPRID2 | -0.678801573 | 1.28E-36 |
| VEGFA | -0.677934401 | 2.04E-11 |
| SHANK3 | -0.677699785 | 2.61E-14 |
| SLC30A3 | -0.676951342 | 9.38E-07 |
| CBX2 | -0.676007387 | 8.73E-11 |
| OBSCN | -0.67545825 | 2.68E-18 |
| NKX2-1 | -0.67538105 | 1.57E-08 |
| PHACTR2 | -0.675208156 | 2.07E-26 |
| WFS1 | -0.674702612 | 6.80E-23 |
| CALB1 | -0.674629006 | 1.88E-08 |
| TXK | -0.673600992 | 4.70E-07 |
| ADGRB3 | -0.672485061 | 4.97E-07 |
| FSD1 | -0.671844471 | 9.53E-06 |
| ACAT2 | -0.671747062 | 1.26E-16 |
| PNPLA1 | -0.670940664 | 1.69E-08 |
| RNF133 | -0.670756093 | 9.65E-06 |
| TDRD10 | -0.66997848 | 1.40E-10 |
| TRH | -0.669900426 | 0.039986119 |
| WIF1 | -0.667980426 | 0.00210635 |
| TMEM74B | -0.667886531 | 5.51E-09 |
| ERVMER34-1 | -0.667383866 | 3.59E-08 |
| SVEP1 | -0.665274754 | 8.70E-08 |
| GRAP | -0.665052451 | 2.17E-12 |
| ID3 | -0.664682755 | 3.35E-09 |
| KIF1C | -0.664439614 | 7.71E-25 |
| IL15RA | -0.66412423 | 3.43E-13 |
| L3MBTL4 | -0.663886452 | 9.75E-42 |
| KLF11 | -0.663814455 | 3.27E-25 |
| DDC | -0.663324721 | 0.001101455 |
| TNFRSF8 | -0.663303491 | 2.10E-08 |
| SNX10 | -0.66324478 | 2.17E-08 |
| FABP4 | -0.663029864 | 0.000273559 |
| ELMSAN1 | -0.662011571 | 2.32E-23 |
| CCL27 | -0.661702195 | 0.007149636 |
| NCR3 | -0.661197033 | 1.54E-05 |
| DNM3 | -0.661147604 | 2.39E-26 |
| WWC3 | -0.660829381 | 2.16E-29 |
| PPP1R15A | -0.660598305 | 3.70E-05 |
| CARHSP1 | -0.66037521 | 1.77E-26 |
| ZIM2 | -0.660345856 | 7.15E-05 |
| TDRD9 | -0.65969166 | 2.47E-06 |
| ACPP | -0.658670236 | 2.03E-08 |
| LEXM | -0.658409888 | 2.64E-06 |
| CASZ1 | -0.658309104 | 7.91E-11 |
| OR2A7 | -0.658084684 | 2.95E-07 |
| TMEM150B | -0.657949332 | 8.20E-07 |
| ARHGEF4 | -0.657932747 | 4.96E-09 |
| VSTM1 | -0.657929204 | 0.015381801 |
| ADAMTS8 | -0.657792658 | 9.02E-09 |
| SULT1B1 | -0.657447285 | 0.004643859 |
| AMBP | -0.6573314 | 0.001200933 |
| DLEU7 | -0.656922457 | 1.07E-07 |
| FRAS1 | -0.656188609 | 3.42E-05 |
| TMEM125 | -0.655225021 | 4.16E-08 |
| AL163636.2 | -0.65419001 | 1.15E-13 |
| FCAR | -0.653947295 | 0.025218193 |
| NEURL1 | -0.653932584 | 2.02E-10 |
| RGL4 | -0.653605884 | 4.11E-05 |
| TMEM64 | -0.653557855 | 4.07E-10 |
| FYB1 | -0.653400014 | 0.000216778 |
| HOGA1 | -0.653139565 | 9.91E-06 |
| PFKFB2 | -0.653007106 | 1.08E-10 |
| CACNB4 | -0.652808337 | 1.27E-12 |
| CD83 | -0.65221038 | 7.49E-06 |
| NRXN1 | -0.651953583 | 6.68E-05 |
| RDX | -0.65046487 | 3.99E-30 |
| MYLK4 | -0.650284346 | 2.00E-11 |
| C12orf49 | -0.650210771 | 9.01E-12 |
| RERGL | -0.649784366 | 0.000514718 |
| XIRP2 | -0.648833061 | 0.004171779 |
| DHRS2 | -0.648342415 | 1.46E-05 |
| INPP5K | -0.647860227 | 5.41E-23 |
| PID1 | -0.647781363 | 1.65E-16 |
| ZSCAN10 | -0.647765648 | 0.006926191 |
| C16orf90 | -0.646887183 | 0.005327692 |
| MTURN | -0.645383905 | 6.14E-17 |
| FGD3 | -0.645135253 | 1.90E-13 |
| SERPINB1 | -0.644617638 | 6.71E-05 |
| TM4SF4 | -0.644349571 | 0.000988587 |
| AC037459.1 | -0.644279832 | 1.79E-06 |
| PMM1 | -0.643185362 | 6.66E-09 |
| GRAPL | -0.642167598 | 6.12E-06 |
| ABCC8 | -0.640646071 | 0.012889809 |
| CCDC194 | -0.639084482 | 3.03E-06 |
| MARCO | -0.638353795 | 2.12E-05 |
| RAG1 | -0.638067283 | 3.35E-14 |
| FAM19A1 | -0.637065532 | 0.024576246 |
| TLR3 | -0.636833737 | 8.09E-14 |
| NUPR1 | -0.636405467 | 2.83E-16 |
| IFI44L | -0.63610496 | 2.75E-05 |
| DUOXA1 | -0.635878487 | 7.03E-06 |
| CEACAM3 | -0.634806722 | 0.000149083 |
| IFNGR1 | -0.63381183 | 1.09E-19 |
| FHDC1 | -0.633441698 | 5.57E-09 |
| CEBPE | -0.63323362 | 6.27E-05 |
| CMTM2 | -0.631980427 | 0.002773363 |
| ROR1 | -0.631574624 | 7.94E-13 |
| SKAP2 | -0.631500577 | 5.17E-05 |
| FCHO2 | -0.631285372 | 0.002868495 |
| TMX4 | -0.630581499 | 0.025143139 |
| CMTM8 | -0.630367888 | 4.64E-09 |
| AC131160.1 | -0.630231067 | 0.007037459 |
| FAM222A | -0.629992717 | 1.91E-05 |
| SLITRK5 | -0.629504389 | 0.000191626 |
| TGM1 | -0.629269545 | 1.67E-07 |
| FOXO4 | -0.62907411 | 7.33E-34 |
| FAM166A | -0.62891017 | 0.007378623 |
| LMCD1 | -0.628423579 | 4.18E-08 |
| KCNJ11 | -0.628010823 | 2.29E-05 |
| DGAT2 | -0.627498397 | 0.004169513 |
| CADM1 | -0.627115874 | 7.61E-11 |
| CAPN9 | -0.626730971 | 0.000100661 |
| HIRIP3 | -0.626132383 | 2.45E-25 |
| LDLR | -0.625962657 | 4.11E-06 |
| ARHGAP29 | -0.625755211 | 2.22E-13 |
| HAPLN1 | -0.624598634 | 0.010736426 |
| CIT | -0.624531453 | 0.000214752 |
| CEACAM19 | -0.62356012 | 1.33E-10 |
| CYP3A7 | -0.623033949 | 0.000134931 |
| ANXA1 | -0.622849957 | 0.000103431 |
| GCA | -0.62169371 | 1.89E-09 |
| SIRPB2 | -0.621337002 | 2.75E-07 |
| RAI2 | -0.621268393 | 2.72E-16 |
| CD226 | -0.621126259 | 5.29E-07 |
| ALOX5 | -0.621099586 | 9.16E-08 |
| NPY1R | -0.619469327 | 0.006243012 |
| KAT2B | -0.618960474 | 3.21E-05 |
| OR2A42 | -0.618846589 | 1.82E-08 |
| PEAK1 | -0.618306103 | 5.95E-24 |
| OSCAR | -0.616956425 | 8.23E-08 |
| AMIGO2 | -0.616679229 | 3.31E-15 |
| ABO | -0.61666982 | 0.019781933 |
| ARGLU1 | -0.61661272 | 2.81E-07 |
| IRF1 | -0.616580638 | 1.21E-05 |
| 43718 | -0.616216096 | 1.25E-32 |
| CBFA2T3 | -0.615866386 | 1.09E-15 |
| FCGR3A | -0.615776914 | 3.89E-06 |
| PTGER4 | -0.615514352 | 5.03E-05 |
| DOK2 | -0.615509004 | 1.24E-07 |
| FGF22 | -0.61530909 | 1.04E-06 |
| SLC11A1 | -0.615129609 | 3.07E-05 |
| PPM1D | -0.614834542 | 1.31E-22 |
| ART5 | -0.614789173 | 3.96E-05 |
| GIMAP1-GIMAP5 | -0.614611913 | 6.58E-10 |
| PLLP | -0.61433995 | 0.000137743 |
| GCH1 | -0.613967881 | 3.44E-07 |
| VSIG4 | -0.613227726 | 1.90E-05 |
| SECISBP2L | -0.612840005 | 4.17E-06 |
| MNDA | -0.61272483 | 0.000363328 |
| PCOLCE2 | -0.612393135 | 6.10E-05 |
| FFAR2 | -0.611869348 | 0.000302369 |
| TSPAN32 | -0.611460588 | 1.47E-08 |
| PREX2 | -0.609448729 | 4.98E-05 |
| TNFSF12 | -0.609434604 | 1.22E-18 |
| IL12A | -0.608582019 | 0.000720271 |
| ETV5 | -0.60816869 | 4.66E-09 |
| ABCC12 | -0.607795952 | 0.020637524 |
| MGLL | -0.607056084 | 3.20E-24 |
| ARHGEF2 | -0.606623083 | 1.28E-14 |
| SDCBP2 | -0.605700625 | 7.42E-06 |
| TBX2 | -0.605318362 | 1.99E-06 |
| NAMPT | -0.604888804 | 0.015089697 |
| VN1R2 | -0.60415034 | 0.00022519 |
| LSS | -0.603293922 | 1.08E-19 |
| GSAP | -0.603219037 | 4.40E-09 |
| MOAP1 | -0.603106408 | 6.54E-08 |
| LYSMD4 | -0.602963217 | 9.74E-24 |
| CNBD1 | -0.602058037 | 2.05E-07 |
| RHOBTB2 | -0.601862164 | 6.61E-09 |
| DGKE | -0.601646652 | 2.92E-07 |
| DAPK1 | -0.601617164 | 2.26E-14 |
| PCDHGB4 | -0.601126672 | 1.29E-16 |
| NFKBIZ | -0.600862456 | 1.60E-05 |
| PALM3 | -0.600411695 | 0.00046983 |
| EFNA1 | -0.599676463 | 3.63E-15 |
| MAGI3 | -0.59942838 | 3.84E-15 |
| MYBPC3 | -0.599332488 | 1.32E-05 |
| CFP | -0.599283033 | 7.91E-08 |
| PPM1K | -0.599132544 | 2.18E-08 |
| NECTIN3 | -0.599132208 | 3.53E-06 |
| GPSM1 | -0.598482816 | 2.97E-12 |
| HK3 | -0.59825427 | 1.15E-06 |
| PLA1A | -0.598167467 | 0.000369344 |
| ZNF765 | -0.597932698 | 8.71E-10 |
| SPIN4 | -0.597931742 | 6.12E-10 |
| CLCN4 | -0.597856407 | 6.30E-06 |
| NARS2 | -0.597493647 | 2.14E-11 |
| AARD | -0.597374646 | 0.000679098 |
| PELI1 | -0.597055318 | 0.000624725 |
| RGSL1 | -0.597004297 | 0.006245576 |
| FGD4 | -0.596822373 | 6.90E-14 |
| SUMO4 | -0.596584145 | 0.00712442 |
| ITGAL | -0.596161757 | 2.62E-08 |
| OPN3 | -0.595811868 | 6.89E-10 |
| SPN | -0.595676386 | 7.48E-07 |
| SLCO2A1 | -0.595403187 | 0.00022088 |
| TRIB1 | -0.594874251 | 0.004729933 |
| CEMIP2 | -0.594593731 | 8.69E-08 |
| RASGRP4 | -0.59458221 | 3.08E-08 |
| SLC26A9 | -0.594389618 | 0.00154434 |
| GHR | -0.594354179 | 6.38E-13 |
| TMEM235 | -0.594061151 | 0.027259106 |
| MYO1C | -0.593981515 | 5.34E-15 |
| SLC44A2 | -0.593901621 | 6.21E-24 |
| NTM | -0.593549368 | 0.0075416 |
| CHCHD4 | -0.592906989 | 7.96E-10 |
| HELZ | -0.592055491 | 4.51E-05 |
| ZDHHC22 | -0.591894589 | 0.009686019 |
| FOXN2 | -0.591852504 | 0.000230098 |
| RAPGEF2 | -0.591261855 | 2.28E-16 |
| MYO1F | -0.591141428 | 5.04E-10 |
| MYO10 | -0.590209477 | 4.35E-15 |
| LSAMP | -0.589856598 | 8.97E-09 |
| NKAPL | -0.589811429 | 3.34E-13 |
| PPP1R9A | -0.589029442 | 6.10E-13 |
| BEX4 | -0.589023911 | 2.27E-12 |
| ZYG11B | -0.588058601 | 0.0036517 |
| PLBD1 | -0.587667053 | 0.001524817 |
| SLC4A1 | -0.587397266 | 3.23E-07 |
| GDPD5 | -0.587361352 | 1.74E-11 |
| PIK3C2B | -0.587328603 | 3.02E-10 |
| PCDHGB3 | -0.586148806 | 0.046082508 |
| PCDH1 | -0.586122257 | 4.02E-11 |
| LINGO4 | -0.586079952 | 0.002744589 |
| REELD1 | -0.58602678 | 3.44E-05 |
| JAKMIP2 | -0.585721529 | 1.17E-05 |
| AK1 | -0.585540819 | 7.14E-14 |
| TMC7 | -0.585516055 | 1.78E-11 |
| SCRN1 | 0.585267519 | 0.020650594 |
| UAP1 | 0.585660828 | 9.82E-06 |
| SHISA9 | 0.585752538 | 0.00096986 |
| BYSL | 0.585804159 | 4.90E-07 |
| SEZ6L2 | 0.585866612 | 1.16E-07 |
| ESPNL | 0.585935556 | 0.001790336 |
| KY | 0.586192377 | 7.01E-06 |
| TRIM9 | 0.586295075 | 7.38E-06 |
| CTSL | 0.586660296 | 2.53E-05 |
| IGSF11 | 0.586865825 | 5.45E-05 |
| ATOH7 | 0.58714899 | 0.000761162 |
| PLA2G2C | 0.587213552 | 0.000597787 |
| HMCN1 | 0.587352244 | 5.90E-10 |
| GVQW2 | 0.587354834 | 1.18E-06 |
| HILPDA | 0.588281799 | 0.002856506 |
| S100B | 0.588309986 | 9.95E-05 |
| ZNF485 | 0.588323833 | 0.017337214 |
| UQCC3 | 0.588393147 | 0.002935991 |
| CENPW | 0.588514885 | 2.19E-12 |
| KDELR3 | 0.588620978 | 0.000237102 |
| PRRX1 | 0.588751527 | 6.52E-05 |
| SNED1 | 0.589258067 | 5.26E-09 |
| TMEM252 | 0.589760606 | 5.83E-06 |
| LRIG3 | 0.590022328 | 2.06E-15 |
| IGBP1 | 0.590288595 | 9.42E-07 |
| ZC2HC1B | 0.590482076 | 0.020522061 |
| BUB1B | 0.590805612 | 6.40E-05 |
| CCNE1 | 0.591572944 | 2.58E-07 |
| PTGS1 | 0.592117978 | 0.001114741 |
| CARMIL2 | 0.593307481 | 1.99E-06 |
| IFT81 | 0.593868632 | 9.29E-11 |
| RHOH | 0.594532176 | 2.61E-06 |
| RPL22L1 | 0.594554077 | 1.40E-07 |
| SLC7A1 | 0.594618823 | 0.027052127 |
| IFITM10 | 0.594751907 | 4.56E-08 |
| MIPEP | 0.594870877 | 1.52E-05 |
| GXYLT2 | 0.594873064 | 1.33E-08 |
| PHLDA1 | 0.59513037 | 1.44E-05 |
| RUNX1 | 0.595221617 | 6.52E-15 |
| AC139491.7 | 0.595237202 | 0.00096108 |
| BTNL2 | 0.595560287 | 0.002729494 |
| KCP | 0.595922699 | 5.02E-07 |
| PTGER3 | 0.595964338 | 0.000178038 |
| HSPA12A | 0.59647508 | 4.96E-09 |
| CDC25A | 0.596641847 | 5.78E-07 |
| C10orf55 | 0.596875116 | 1.70E-08 |
| HSD3B7 | 0.596930811 | 3.37E-06 |
| ICOSLG | 0.596945439 | 8.86E-09 |
| MAP9 | 0.597314518 | 0.000545891 |
| OVOL3 | 0.597338631 | 0.001907347 |
| HSPG2 | 0.59745081 | 4.52E-08 |
| CA3 | 0.597456521 | 0.016581536 |
| PCLAF | 0.597483163 | 3.01E-05 |
| BTC | 0.597886291 | 0.001432628 |
| RAB19 | 0.597989176 | 2.23E-07 |
| ABCB5 | 0.598263497 | 0.000116986 |
| VILL | 0.598680689 | 3.74E-15 |
| ZNF20 | 0.598746289 | 5.84E-07 |
| P3H4 | 0.599160553 | 2.83E-11 |
| MMRN1 | 0.599582628 | 2.39E-05 |
| DLG4 | 0.599608161 | 7.12E-08 |
| RAD54L | 0.60060064 | 1.83E-07 |
| CCR4 | 0.600931007 | 0.000107224 |
| RUBCNL | 0.600970828 | 2.98E-05 |
| NME1 | 0.601232642 | 1.06E-06 |
| CALML6 | 0.602431908 | 2.61E-06 |
| SRPX | 0.602464812 | 1.28E-05 |
| HSPBP1 | 0.602890134 | 4.34E-10 |
| TLR9 | 0.603144331 | 1.11E-05 |
| FDXR | 0.60355582 | 1.89E-09 |
| SLC30A10 | 0.603707976 | 0.003507281 |
| PCOLCE | 0.60398174 | 5.73E-09 |
| UGDH | 0.604384371 | 1.98E-05 |
| VSNL1 | 0.604636736 | 3.33E-07 |
| PKM | 0.604777077 | 3.61E-22 |
| RIN1 | 0.604890719 | 2.74E-08 |
| CATIP | 0.605676409 | 0.000127213 |
| C1R | 0.605721085 | 1.23E-07 |
| TTC24 | 0.605880973 | 0.001169566 |
| KCNH5 | 0.606828836 | 0.002066304 |
| CROT | 0.608850029 | 3.74E-21 |
| NEURL1B | 0.609087124 | 2.01E-09 |
| SEZ6L | 0.609380051 | 0.001568275 |
| BICDL2 | 0.609641027 | 3.02E-07 |
| SDSL | 0.609709765 | 2.97E-07 |
| NUCB2 | 0.609774942 | 1.97E-14 |
| AC137834.1 | 0.609893594 | 0.002574509 |
| SMIM5 | 0.609934544 | 5.17E-06 |
| GRAMD2B | 0.610212384 | 1.30E-11 |
| HSD11B2 | 0.61146067 | 0.000423773 |
| FBLN7 | 0.611494538 | 1.72E-08 |
| ADORA2B | 0.611572621 | 2.43E-09 |
| HAMP | 0.611695408 | 0.002179356 |
| SLC35F3 | 0.611826493 | 0.036883963 |
| RAET1G | 0.612013777 | 1.10E-06 |
| STMN3 | 0.612230343 | 0.000250376 |
| ZNF695 | 0.61264272 | 0.00035662 |
| LOXL2 | 0.613592786 | 2.39E-08 |
| E2F7 | 0.61412654 | 0.027956448 |
| SEC11C | 0.614290426 | 6.20E-18 |
| MROH2B | 0.61513723 | 0.006569757 |
| SCARA5 | 0.615560126 | 0.010702324 |
| CTSK | 0.615827091 | 1.79E-05 |
| FUT3 | 0.616149737 | 9.37E-06 |
| KLK10 | 0.616258428 | 0.000106703 |
| PPM1J | 0.616334899 | 1.21E-09 |
| PWWP3B | 0.617539778 | 0.000123577 |
| STYK1 | 0.617637112 | 8.05E-05 |
| MFAP1 | 0.617776478 | 0.018997929 |
| RGS20 | 0.617778504 | 1.48E-05 |
| TNFSF18 | 0.618793588 | 0.000459409 |
| AC093899.2 | 0.619239031 | 9.91E-06 |
| ANKRD53 | 0.619254813 | 4.92E-09 |
| MICALL2 | 0.61954154 | 4.84E-13 |
| TEDC2 | 0.619683975 | 8.24E-06 |
| GNL3 | 0.620071724 | 1.51E-11 |
| CPLX4 | 0.620346522 | 0.001666005 |
| HOXC9 | 0.620562853 | 0.043490661 |
| PLXNB1 | 0.620608743 | 2.65E-12 |
| UCK2 | 0.620621292 | 5.06E-08 |
| PDLIM7 | 0.621073818 | 4.38E-12 |
| PSD | 0.621729669 | 2.67E-25 |
| NR1I2 | 0.621774833 | 1.09E-07 |
| AQP7 | 0.621817436 | 9.27E-08 |
| HHAT | 0.622250069 | 3.02E-15 |
| RGPD1 | 0.622300572 | 0.001063174 |
| SPAG5 | 0.62232162 | 3.01E-08 |
| WNT10B | 0.622403197 | 0.027786802 |
| TUBB2A | 0.622662998 | 0.0001598 |
| FOSL1 | 0.623661473 | 0.040277563 |
| SH3PXD2B | 0.623912591 | 0.004340181 |
| VWDE | 0.624276937 | 3.54E-05 |
| KLHL6 | 0.624344294 | 1.98E-11 |
| FREM1 | 0.625255779 | 7.29E-09 |
| MPP2 | 0.626477477 | 7.02E-11 |
| BCL2L14 | 0.626972231 | 7.91E-09 |
| THBS3 | 0.627149795 | 1.52E-17 |
| FUT8 | 0.627620887 | 2.54E-32 |
| SLC52A3 | 0.627817288 | 0.014453608 |
| MEIKIN | 0.627860405 | 0.000194717 |
| MYADML2 | 0.627967793 | 0.002126132 |
| BBS5 | 0.628854373 | 5.68E-16 |
| TNIP3 | 0.629243227 | 0.000279919 |
| LRRK1 | 0.630288633 | 1.43E-29 |
| AL669918.1 | 0.630638258 | 0.000239633 |
| ARNT2 | 0.630838486 | 0.001742856 |
| SOX8 | 0.631125377 | 0.000257601 |
| MRTO4 | 0.631289843 | 1.42E-09 |
| EPS8L1 | 0.631548626 | 3.14E-12 |
| FZD3 | 0.631971975 | 8.18E-07 |
| FAM180A | 0.632598493 | 0.001233073 |
| MYLK | 0.632637163 | 8.10E-11 |
| SLC9A4 | 0.632686049 | 9.44E-05 |
| KIAA0319 | 0.632769703 | 0.001407359 |
| OLFML2B | 0.633665467 | 3.61E-08 |
| MAD2L1 | 0.633989414 | 5.57E-09 |
| PPIC | 0.634309124 | 1.83E-20 |
| EFNA5 | 0.634432615 | 1.40E-13 |
| CEP55 | 0.635006159 | 2.14E-05 |
| COLCA2 | 0.635015818 | 1.43E-08 |
| DGKB | 0.635383153 | 5.64E-05 |
| AC092718.3 | 0.635561022 | 7.72E-11 |
| CXCL17 | 0.635631532 | 3.13E-07 |
| MAGEE2 | 0.636034212 | 7.51E-07 |
| C16orf45 | 0.636324497 | 2.17E-14 |
| STAG3 | 0.636851477 | 2.40E-11 |
| RASGRF2 | 0.637242616 | 7.33E-07 |
| HOXB8 | 0.637291347 | 0.000323363 |
| IL1R1 | 0.637450236 | 1.26E-08 |
| GLI1 | 0.638447871 | 1.07E-06 |
| CHL1 | 0.63931514 | 3.58E-07 |
| CPB1 | 0.63943136 | 0.001699407 |
| RAB6D | 0.63944963 | 0.026502578 |
| EPS8L3 | 0.639718187 | 0.011297692 |
| SELP | 0.640569755 | 1.63E-10 |
| RASSF10 | 0.640946001 | 0.000104626 |
| LRRN2 | 0.640969468 | 2.85E-08 |
| CCNB1 | 0.641816019 | 4.15E-09 |
| TUBB2B | 0.641970372 | 0.006493248 |
| CENPS-CORT | 0.642161565 | 2.51E-05 |
| C21orf59-TCP10L | 0.642653146 | 0.004001362 |
| AC243967.1 | 0.642799707 | 0.00831041 |
| AC087289.1 | 0.643449217 | 2.78E-15 |
| NCAM1 | 0.644009383 | 6.56E-10 |
| TTLL13P | 0.644059348 | 3.34E-05 |
| CNTD1 | 0.644462378 | 1.97E-06 |
| TYMS | 0.644905012 | 7.24E-12 |
| PPIAL4E | 0.645275901 | 0.019964908 |
| TENM3 | 0.645503384 | 0.005111507 |
| FP565260.3 | 0.64696265 | 7.33E-06 |
| BLNK | 0.64718139 | 2.36E-12 |
| ADAD2 | 0.648012216 | 0.000757114 |
| CD70 | 0.648198175 | 1.52E-06 |
| STK32B | 0.648302523 | 9.49E-07 |
| PPP1R1A | 0.64833523 | 0.003524428 |
| TOX3 | 0.648901873 | 0.003123716 |
| NLRC3 | 0.649050024 | 7.57E-06 |
| ARHGEF33 | 0.64939311 | 0.000309397 |
| GPI | 0.649404109 | 6.33E-11 |
| EPHB3 | 0.649451874 | 0.000219525 |
| CEP41 | 0.650751176 | 9.02E-13 |
| SIT1 | 0.65077121 | 0.002331319 |
| SMCO2 | 0.651115964 | 2.79E-06 |
| CACNA1A | 0.651605008 | 9.08E-08 |
| FOXL1 | 0.652281815 | 6.15E-09 |
| CNN1 | 0.652784793 | 1.88E-05 |
| MOK | 0.652788517 | 8.42E-10 |
| LUM | 0.653424034 | 1.71E-08 |
| CNTD2 | 0.653549223 | 3.43E-05 |
| ANKRD30BL | 0.653961934 | 0.000456802 |
| SH3PXD2A | 0.654645926 | 6.08E-18 |
| DCX | 0.654769082 | 0.024515703 |
| PLIN4 | 0.65528219 | 4.55E-07 |
| ZNF154 | 0.655338656 | 1.18E-17 |
| PTP4A3 | 0.655772585 | 1.04E-12 |
| C1QTNF3 | 0.655938906 | 1.17E-07 |
| GLIS3 | 0.656693843 | 3.45E-09 |
| DNAH1 | 0.656788737 | 9.24E-10 |
| GPR78 | 0.656803945 | 0.00032551 |
| ANKMY1 | 0.657059549 | 6.15E-11 |
| TMEM176A | 0.657136536 | 3.43E-11 |
| CDH22 | 0.657180834 | 2.97E-05 |
| RPGRIP1L | 0.657458866 | 6.53E-13 |
| UGT2B11 | 0.657660669 | 0.035778382 |
| METTL24 | 0.657810393 | 6.89E-12 |
| CYP2W1 | 0.65794987 | 0.048750298 |
| KCNE4 | 0.658251027 | 5.69E-10 |
| KCTD4 | 0.65825576 | 0.014955311 |
| PRSS50 | 0.658292157 | 3.34E-06 |
| SHD | 0.658326531 | 0.024164758 |
| TMC3 | 0.658955485 | 8.16E-05 |
| CNTNAP2 | 0.659056108 | 6.62E-11 |
| SPACA9 | 0.65956628 | 7.61E-07 |
| AC007998.2 | 0.65975943 | 0.029989186 |
| OLFM1 | 0.660169694 | 1.30E-12 |
| ANGPTL2 | 0.66050422 | 2.52E-07 |
| IRF6 | 0.660943919 | 4.08E-07 |
| CRACR2B | 0.660958443 | 1.11E-09 |
| PAQR7 | 0.661248747 | 5.40E-32 |
| CENPF | 0.661248989 | 2.16E-08 |
| ISL2 | 0.662071948 | 0.016260794 |
| UBL4B | 0.662115652 | 0.001376921 |
| AC092042.3 | 0.662514642 | 2.74E-10 |
| TMED3 | 0.663186273 | 6.82E-12 |
| DCPS | 0.66339622 | 0.000550632 |
| SPAG16 | 0.663646005 | 1.03E-10 |
| MTRNR2L12 | 0.663651149 | 0.003659899 |
| PCDH20 | 0.663821892 | 5.10E-07 |
| PRIMA1 | 0.664405357 | 0.001065252 |
| CDCA5 | 0.66462372 | 0.000240306 |
| CCDC34 | 0.664828268 | 1.48E-08 |
| SMTNL2 | 0.664961298 | 5.43E-08 |
| MYOM1 | 0.665354779 | 6.80E-11 |
| BASP1 | 0.665398988 | 4.94E-07 |
| TRAF5 | 0.665593299 | 2.55E-13 |
| NPAS2 | 0.666122865 | 1.51E-07 |
| TMEM67 | 0.666428644 | 7.88E-08 |
| FAM81A | 0.666800763 | 6.28E-08 |
| RBP5 | 0.667388015 | 1.51E-13 |
| NTN3 | 0.667638217 | 7.58E-06 |
| CPEB1 | 0.66826566 | 5.54E-11 |
| SLC16A1 | 0.668337986 | 2.20E-09 |
| PSENEN | 0.668410546 | 6.60E-14 |
| THPO | 0.668886533 | 1.54E-11 |
| CLBA1 | 0.669232765 | 8.80E-09 |
| GGN | 0.670102337 | 8.68E-07 |
| C1QTNF1 | 0.670443207 | 2.18E-06 |
| GABRR1 | 0.670682767 | 0.048880444 |
| MAP1B | 0.670833322 | 0.049884737 |
| AGR3 | 0.670834828 | 4.09E-06 |
| ADAMTS4 | 0.671128552 | 0.04269817 |
| ENPP5 | 0.671375403 | 5.01E-11 |
| LRRC70 | 0.671528244 | 2.78E-07 |
| HIST1H2AM | 0.671692614 | 0.026061297 |
| VNN3 | 0.672194343 | 0.000560456 |
| DUSP27 | 0.672196041 | 0.000820867 |
| KLK11 | 0.672839498 | 4.25E-05 |
| LHPP | 0.672848739 | 1.15E-14 |
| PDE2A | 0.674148781 | 2.13E-09 |
| SMAD3 | 0.674151397 | 2.58E-06 |
| TJP3 | 0.674890457 | 6.93E-08 |
| TPSB2 | 0.675253759 | 5.73E-05 |
| GRM2 | 0.675636336 | 2.49E-07 |
| ZC3H12D | 0.675996407 | 1.30E-10 |
| BICC1 | 0.676311228 | 4.39E-16 |
| HP | 0.676325196 | 0.007808783 |
| PARD6G | 0.676421813 | 2.04E-09 |
| SPATA24 | 0.676777655 | 5.22E-14 |
| SLC18A2 | 0.67679363 | 1.64E-05 |
| PRKY | 0.67702633 | 0.03064443 |
| HS3ST2 | 0.677491317 | 8.38E-06 |
| ADCY5 | 0.678958563 | 4.90E-10 |
| TCTN1 | 0.68016337 | 3.66E-15 |
| NPY4R2 | 0.680815244 | 0.002552492 |
| HAPLN3 | 0.680893186 | 3.75E-06 |
| PLXDC1 | 0.680893966 | 7.70E-13 |
| FAM221B | 0.681068334 | 4.08E-05 |
| ANGPTL6 | 0.681071006 | 2.04E-09 |
| BTLA | 0.682047773 | 6.98E-05 |
| KCND1 | 0.683843916 | 2.62E-05 |
| KLK14 | 0.684043811 | 0.00053659 |
| KRT222 | 0.684186803 | 0.000152435 |
| AC004832.3 | 0.684229606 | 0.000743176 |
| FBLN1 | 0.684280729 | 1.87E-07 |
| CLEC11A | 0.684528188 | 0.027716957 |
| STX1A | 0.684585229 | 7.27E-11 |
| UHRF1 | 0.684651319 | 2.88E-08 |
| COL20A1 | 0.684663692 | 0.000375216 |
| TNNT3 | 0.684699712 | 0.000221431 |
| AC092073.1 | 0.685527593 | 0.000400254 |
| GRIP1 | 0.68559957 | 2.82E-07 |
| IQCK | 0.686015968 | 8.22E-11 |
| LYPD2 | 0.686031874 | 0.020671092 |
| PDZD3 | 0.686849385 | 0.000357724 |
| NUTM1 | 0.686942784 | 0.008483253 |
| ZNF157 | 0.68706485 | 9.44E-05 |
| AP3B2 | 0.687620833 | 2.14E-10 |
| NPR2 | 0.687642969 | 1.11E-08 |
| MYPN | 0.688193789 | 5.62E-06 |
| TP53INP1 | 0.68858223 | 1.82E-12 |
| NLRP4 | 0.688669378 | 0.043294008 |
| SLC22A14 | 0.689013752 | 2.90E-06 |
| HJV | 0.689046728 | 0.001801723 |
| ACRV1 | 0.689490179 | 7.90E-06 |
| RUVBL1 | 0.689746872 | 7.31E-19 |
| PDIA4 | 0.689778089 | 2.34E-12 |
| RFX3 | 0.690112066 | 4.47E-08 |
| DTX1 | 0.69014905 | 8.57E-07 |
| CYYR1 | 0.690230551 | 0.000154256 |
| PYCR1 | 0.691097085 | 3.01E-07 |
| OSBPL3 | 0.691262086 | 1.49E-12 |
| SLC45A3 | 0.691406657 | 3.34E-09 |
| MELK | 0.691489694 | 1.03E-06 |
| ZNF487 | 0.692378672 | 6.18E-07 |
| SLC24A2 | 0.692443584 | 2.36E-08 |
| DLGAP2 | 0.692502979 | 0.000203974 |
| SYNGR4 | 0.693204472 | 0.049824026 |
| STRIP2 | 0.693237556 | 0.001220264 |
| CHRD | 0.693272679 | 5.78E-13 |
| MUC3A | 0.693439393 | 0.000147397 |
| CHEK2 | 0.693640594 | 3.27E-30 |
| AL049697.1 | 0.693817461 | 5.29E-06 |
| BEAN1 | 0.693939743 | 5.96E-16 |
| ABCB1 | 0.694274874 | 2.13E-07 |
| FBXO31 | 0.694858883 | 1.41E-21 |
| ZNF440 | 0.69595348 | 3.43E-09 |
| TMC4 | 0.696229604 | 2.12E-14 |
| TENM2 | 0.696945465 | 1.47E-06 |
| STX1B | 0.697175301 | 0.005495962 |
| ZBP1 | 0.697449376 | 8.94E-10 |
| PIGR | 0.697652185 | 0.001001156 |
| TSGA10 | 0.698699724 | 4.20E-10 |
| C14orf39 | 0.698886499 | 0.016919346 |
| ANKRD18A | 0.699061212 | 7.69E-06 |
| KIF21A | 0.699597539 | 2.18E-07 |
| KCNMA1 | 0.70028659 | 7.17E-11 |
| OTOL1 | 0.700400169 | 0.002080561 |
| CLUAP1 | 0.700456248 | 1.91E-09 |
| TMEM117 | 0.701784942 | 2.70E-09 |
| GSDMA | 0.701805478 | 3.32E-05 |
| GALNT14 | 0.703927678 | 8.62E-07 |
| MSI2 | 0.704113485 | 2.68E-13 |
| TUB | 0.70418459 | 6.49E-13 |
| HRH1 | 0.704199411 | 4.54E-09 |
| MT1H | 0.704220431 | 0.028252551 |
| SPINK5 | 0.704946037 | 3.81E-08 |
| COL6A2 | 0.705125572 | 4.12E-13 |
| RPP40 | 0.705352622 | 4.11E-16 |
| TRIM31 | 0.705576957 | 2.90E-06 |
| GJB7 | 0.705639995 | 0.006554311 |
| AL583836.1 | 0.70657602 | 0.010320849 |
| TTYH1 | 0.706701856 | 1.42E-12 |
| SYTL2 | 0.706841569 | 7.18E-18 |
| FAAH2 | 0.70791607 | 1.23E-13 |
| ZNF80 | 0.70792263 | 1.80E-05 |
| NAT14 | 0.70886064 | 6.51E-10 |
| TNFRSF19 | 0.708863593 | 4.79E-15 |
| TPM2 | 0.708881358 | 6.24E-09 |
| MORN1 | 0.708941784 | 7.42E-13 |
| MAPK8IP1 | 0.7091697 | 7.94E-14 |
| PAPLN | 0.709683587 | 8.32E-20 |
| ZFP69B | 0.70987214 | 4.09E-13 |
| RGS5 | 0.710143321 | 2.52E-05 |
| IFT22 | 0.710144888 | 3.78E-12 |
| STK36 | 0.710251922 | 4.44E-13 |
| ROBO1 | 0.711771496 | 4.86E-15 |
| DTX2 | 0.712141383 | 3.43E-24 |
| TIGIT | 0.712600454 | 7.76E-05 |
| PCSK4 | 0.712608918 | 6.34E-09 |
| KREMEN2 | 0.712903617 | 1.02E-10 |
| FBXL13 | 0.713180507 | 1.06E-07 |
| AEN | 0.713199306 | 5.89E-09 |
| TSACC | 0.713449555 | 1.14E-10 |
| OMP | 0.713718698 | 0.006288923 |
| EHF | 0.71448302 | 5.30E-06 |
| GPR19 | 0.714755468 | 6.92E-09 |
| HSPB7 | 0.714760466 | 9.20E-07 |
| C9orf43 | 0.715056804 | 1.61E-09 |
| CYP11B1 | 0.715078185 | 0.044877025 |
| SV2B | 0.71540877 | 5.93E-08 |
| SLC16A14 | 0.715864265 | 3.01E-05 |
| C1orf61 | 0.715902954 | 1.98E-09 |
| VSIG8 | 0.716098907 | 2.19E-06 |
| NPIPB6 | 0.716956836 | 3.53E-07 |
| REM2 | 0.717584719 | 0.000420493 |
| WNT1 | 0.717809163 | 1.47E-06 |
| MIOX | 0.718506055 | 2.09E-07 |
| LOX | 0.718764621 | 2.24E-05 |
| LDLRAD3 | 0.719285469 | 0.000126932 |
| TRPM6 | 0.719893402 | 1.59E-08 |
| CDKL4 | 0.719974046 | 0.000449987 |
| ADGRF2 | 0.720322949 | 0.000478642 |
| TNFRSF9 | 0.720838024 | 7.24E-06 |
| GNG14 | 0.721317015 | 0.011448095 |
| PODN | 0.721625669 | 1.96E-08 |
| SLC25A12 | 0.721984136 | 1.09E-15 |
| IFT27 | 0.722367004 | 3.78E-11 |
| CCDC110 | 0.722995672 | 1.23E-07 |
| CC2D2A | 0.72319564 | 9.15E-12 |
| SLFN13 | 0.723283201 | 2.21E-12 |
| TTC30B | 0.723353243 | 3.78E-08 |
| MMP27 | 0.723452247 | 0.04866891 |
| TRO | 0.723857551 | 5.50E-13 |
| BX276092.9 | 0.725132728 | 0.005599618 |
| FBLIM1 | 0.725153816 | 2.20E-15 |
| ADH1C | 0.725745717 | 0.002169138 |
| TPSAB1 | 0.726293233 | 2.35E-06 |
| SEMA3C | 0.727004547 | 0.000806531 |
| CELSR2 | 0.7270062 | 2.62E-12 |
| ACTC1 | 0.727100282 | 0.01050758 |
| ZMAT3 | 0.727619829 | 2.82E-06 |
| SLC16A2 | 0.727813478 | 2.86E-12 |
| MYB | 0.728236166 | 1.74E-06 |
| IGSF1 | 0.728271596 | 1.11E-05 |
| LVRN | 0.728366439 | 9.82E-05 |
| PPP1R32 | 0.728517655 | 7.94E-08 |
| COL4A5 | 0.728715877 | 5.89E-15 |
| LTA | 0.728759622 | 1.41E-06 |
| AQP10 | 0.731465711 | 0.000133309 |
| POLR1E | 0.731541997 | 0.000171318 |
| LTB | 0.732542181 | 1.44E-06 |
| MFSD6L | 0.732842549 | 9.19E-11 |
| SPOCD1 | 0.733060659 | 1.65E-07 |
| SLC12A8 | 0.733254771 | 2.39E-14 |
| PTGIS | 0.73347352 | 0.026648603 |
| RUNX2 | 0.733622921 | 7.29E-20 |
| KRT8 | 0.733671024 | 1.66E-08 |
| MRPS31 | 0.73530984 | 4.84E-12 |
| RABL2B | 0.736009985 | 3.77E-11 |
| GCKR | 0.736220892 | 1.75E-13 |
| C15orf65 | 0.736642794 | 2.39E-08 |
| CTSG | 0.737222818 | 0.015121697 |
| TBX10 | 0.737263939 | 0.006691172 |
| DZIP3 | 0.737510149 | 1.36E-07 |
| LHX6 | 0.738051766 | 6.22E-09 |
| SPNS3 | 0.7386641 | 5.10E-11 |
| AC097637.1 | 0.739454847 | 1.25E-08 |
| HNF1A | 0.740239015 | 0.00075241 |
| LGR6 | 0.740819752 | 3.21E-08 |
| NCAPH | 0.741264152 | 1.88E-07 |
| CDKN3 | 0.741703206 | 9.21E-08 |
| SLC4A5 | 0.741923597 | 7.45E-13 |
| S100P | 0.742169515 | 0.000304952 |
| KLHDC9 | 0.742797911 | 4.42E-08 |
| CHPF | 0.742843499 | 1.59E-17 |
| ANKRD7 | 0.743033898 | 7.14E-08 |
| GMDS | 0.743448016 | 4.76E-22 |
| COL23A1 | 0.743764994 | 2.05E-11 |
| DZANK1 | 0.744696685 | 9.70E-10 |
| SMC1B | 0.744933373 | 0.005372154 |
| CDT1 | 0.744953646 | 0.000888274 |
| TEKT3 | 0.745360521 | 1.09E-06 |
| STK32A | 0.746017709 | 2.98E-08 |
| SHANK1 | 0.746079838 | 4.51E-12 |
| RBM20 | 0.746370239 | 4.43E-06 |
| ITGB3 | 0.746878942 | 6.52E-10 |
| STRC | 0.74706472 | 3.58E-08 |
| GMNC | 0.748384287 | 0.039589652 |
| YBX2 | 0.748683567 | 8.46E-06 |
| OR10A2 | 0.749129956 | 8.04E-05 |
| KIF9 | 0.750200377 | 8.38E-12 |
| SLC28A2 | 0.750316077 | 0.043365018 |
| TCN1 | 0.750768667 | 0.034828737 |
| MSH5-SAPCD1 | 0.752080014 | 5.08E-13 |
| EFCAB11 | 0.752570556 | 1.98E-18 |
| ABCG4 | 0.752872886 | 0.002324172 |
| PFKP | 0.753715353 | 6.97E-14 |
| GPR135 | 0.753759258 | 8.61E-15 |
| IFT172 | 0.754345888 | 1.95E-10 |
| TAGLN | 0.754414691 | 1.51E-10 |
| USP43 | 0.754483913 | 3.62E-07 |
| CA7 | 0.754524745 | 0.029962354 |
| C1S | 0.75486906 | 8.58E-18 |
| AC091167.2 | 0.754910463 | 8.20E-08 |
| ANTXRL | 0.754989487 | 6.14E-05 |
| AC112229.3 | 0.755308978 | 0.000174174 |
| ANXA8 | 0.75581155 | 2.59E-05 |
| CCL11 | 0.756557668 | 0.001855329 |
| FCGBP | 0.756596338 | 8.99E-06 |
| IGFBP7 | 0.757136077 | 1.15E-17 |
| RSPO3 | 0.757177252 | 1.52E-05 |
| LUZP2 | 0.757707835 | 0.000775266 |
| UGT1A7 | 0.758327528 | 1.15E-06 |
| SYN1 | 0.759002317 | 5.98E-06 |
| CADM3 | 0.759590846 | 1.90E-05 |
| EFHB | 0.760052081 | 3.50E-06 |
| KRT19 | 0.761152076 | 5.78E-11 |
| IQCA1L | 0.761415952 | 0.026661273 |
| HIST1H4J | 0.762018654 | 6.63E-16 |
| DNAJA4 | 0.762421085 | 3.81E-06 |
| PROB1 | 0.762434892 | 2.44E-20 |
| NLRP11 | 0.762705538 | 1.09E-05 |
| SLC6A8 | 0.762759895 | 1.26E-06 |
| TACR2 | 0.76345453 | 2.35E-09 |
| TMC5 | 0.76440103 | 9.42E-11 |
| FAM57A | 0.764431811 | 4.33E-14 |
| GASK1B | 0.764917842 | 1.78E-15 |
| TICRR | 0.764997845 | 6.77E-19 |
| IGSF5 | 0.76563376 | 1.51E-07 |
| LCA5 | 0.765646087 | 1.22E-05 |
| GLI2 | 0.766427139 | 1.82E-15 |
| DDIT4 | 0.767557023 | 9.02E-07 |
| GPT2 | 0.767615221 | 1.23E-09 |
| SMOX | 0.767655632 | 4.74E-08 |
| AC010531.1 | 0.767949064 | 1.98E-05 |
| UPP2 | 0.7684636 | 1.91E-05 |
| DLG2 | 0.769297662 | 1.23E-18 |
| ALDH1L1 | 0.769552647 | 0.00149336 |
| FAM78B | 0.770093988 | 6.14E-16 |
| MEIS3 | 0.770231351 | 6.19E-20 |
| GDPD2 | 0.770823144 | 4.93E-11 |
| SPARC | 0.770890649 | 3.92E-08 |
| GALNT6 | 0.770978376 | 1.17E-15 |
| CLEC19A | 0.771545343 | 2.27E-06 |
| ADCY2 | 0.771674394 | 1.54E-13 |
| MEF2B | 0.772222359 | 1.85E-13 |
| SLC9C2 | 0.772363201 | 0.000941564 |
| DYNC2H1 | 0.772932088 | 2.65E-10 |
| FOXD4L6 | 0.774046092 | 0.014535616 |
| ABCC3 | 0.774229808 | 1.48E-17 |
| C11orf53 | 0.774307609 | 0.000640095 |
| SMIM32 | 0.774323884 | 0.013268201 |
| INTU | 0.774481349 | 7.66E-10 |
| TMEM210 | 0.774693431 | 0.0013259 |
| PPP1R14B | 0.774775725 | 1.68E-12 |
| HEATR1 | 0.775068545 | 2.91E-07 |
| FOXA1 | 0.775519086 | 1.80E-06 |
| ADAMTS3 | 0.775716462 | 0.0013259 |
| SERTAD4 | 0.776407238 | 1.72E-07 |
| PDGFD | 0.776603258 | 1.20E-05 |
| RHPN1 | 0.776696239 | 2.41E-13 |
| VAV3 | 0.778418462 | 1.34E-09 |
| ROPN1 | 0.77857282 | 0.000307441 |
| MTRNR2L8 | 0.77919506 | 0.000862486 |
| XBP1 | 0.779316743 | 9.63E-22 |
| BDH1 | 0.78015809 | 1.05E-09 |
| FKBP10 | 0.780866007 | 3.75E-15 |
| LYNX1-SLURP2 | 0.781075402 | 1.84E-11 |
| PSG3 | 0.781551285 | 0.019931028 |
| SNCAIP | 0.7816574 | 2.27E-18 |
| COX8C | 0.781967908 | 0.008825125 |
| ENC1 | 0.782334272 | 6.15E-23 |
| PIWIL3 | 0.783252839 | 0.007891643 |
| BCO2 | 0.78335953 | 2.36E-09 |
| EGLN3 | 0.784036854 | 3.56E-08 |
| NEK2 | 0.784923998 | 1.22E-09 |
| C15orf48 | 0.785147408 | 7.65E-05 |
| HIST3H2A | 0.785522811 | 1.29E-10 |
| EDN2 | 0.785602631 | 0.038548684 |
| EGR4 | 0.786032318 | 0.009821086 |
| DNAAF4 | 0.786393813 | 3.01E-10 |
| COL24A1 | 0.786443929 | 3.44E-05 |
| MISP | 0.786470885 | 0.00231676 |
| HTRA1 | 0.787175438 | 5.53E-16 |
| GLT8D2 | 0.787862052 | 2.17E-17 |
| MSC | 0.788641455 | 2.75E-05 |
| C10orf53 | 0.788721871 | 8.13E-05 |
| C22orf42 | 0.789633505 | 3.55E-09 |
| TSPAN8 | 0.790521427 | 3.39E-06 |
| LPAR3 | 0.791255308 | 1.23E-06 |
| C3 | 0.791274598 | 1.31E-11 |
| TSPAN6 | 0.791409775 | 5.29E-18 |
| EXTL1 | 0.791675807 | 3.42E-10 |
| SMYD3 | 0.791770829 | 3.43E-15 |
| C19orf67 | 0.791920556 | 2.25E-06 |
| IL17RD | 0.792316748 | 0.008632828 |
| CCDC136 | 0.792734053 | 2.77E-15 |
| TRPA1 | 0.792949078 | 1.47E-06 |
| CLMN | 0.793832167 | 2.30E-14 |
| FLG2 | 0.794245963 | 6.27E-06 |
| ARHGEF37 | 0.795131019 | 1.14E-05 |
| IER5L | 0.795391445 | 1.70E-10 |
| TGFBI | 0.795586615 | 2.22E-13 |
| ARMH4 | 0.796569427 | 3.14E-15 |
| ELK3 | 0.796586949 | 3.11E-09 |
| TEX46 | 0.798381121 | 0.017230593 |
| HEPH | 0.799166024 | 2.62E-18 |
| AC008687.8 | 0.800038981 | 1.69E-08 |
| MYBPC2 | 0.800378226 | 0.018311863 |
| CBLN1 | 0.800388109 | 0.04474928 |
| CCDC30 | 0.800404092 | 5.08E-09 |
| DUSP5 | 0.801070315 | 0.001512132 |
| CXXC4 | 0.801099666 | 0.002338171 |
| STMN2 | 0.801156721 | 2.02E-05 |
| ZBTB7C | 0.801286987 | 6.98E-14 |
| GADD45A | 0.802658327 | 1.19E-06 |
| GH1 | 0.803938996 | 0.026144942 |
| SORD | 0.804542873 | 2.18E-12 |
| IL4 | 0.804935713 | 0.000487595 |
| PCBP3 | 0.805416793 | 3.05E-17 |
| ST18 | 0.805734305 | 4.57E-06 |
| ADGRF1 | 0.8058881 | 7.79E-07 |
| THSD7B | 0.80608045 | 0.008526685 |
| COL6A1 | 0.806348369 | 3.21E-15 |
| SLC38A8 | 0.807292926 | 0.008162679 |
| THEGL | 0.808110639 | 0.000713915 |
| CDK14 | 0.809886943 | 1.39E-11 |
| SLC29A1 | 0.810307931 | 0.022154527 |
| KLHDC7B | 0.810665518 | 0.001270575 |
| KIAA1211 | 0.810783183 | 7.23E-06 |
| SMPDL3B | 0.810795851 | 3.19E-11 |
| CADM4 | 0.810909242 | 9.19E-11 |
| HLA-DOB | 0.811088063 | 3.44E-09 |
| PCSK1 | 0.811320533 | 0.004107659 |
| C4orf48 | 0.811601648 | 9.84E-15 |
| TRIM63 | 0.812080435 | 2.03E-06 |
| HORMAD1 | 0.812225144 | 6.23E-06 |
| ADH6 | 0.812289672 | 1.12E-06 |
| FRMD6 | 0.812784103 | 2.29E-16 |
| DDIT4L | 0.812811366 | 1.54E-06 |
| NTF3 | 0.81342267 | 9.43E-08 |
| PLEK2 | 0.81364753 | 1.10E-06 |
| CYP2S1 | 0.813875455 | 0.000333673 |
| FCMR | 0.814079065 | 1.19E-14 |
| SAP30 | 0.814132513 | 1.32E-08 |
| RHBDL3 | 0.814510681 | 1.33E-08 |
| SEMA6B | 0.815144952 | 0.000132737 |
| AC087632.1 | 0.815567524 | 0.000155606 |
| ZDHHC1 | 0.815799797 | 2.49E-12 |
| NME7 | 0.815918222 | 7.89E-12 |
| MYRIP | 0.817313494 | 5.13E-09 |
| CCDC197 | 0.817895956 | 0.000886211 |
| ESRRG | 0.818378888 | 2.55E-08 |
| KCNMB2 | 0.818602048 | 4.12E-06 |
| CALR3 | 0.818984879 | 0.010315992 |
| TBX1 | 0.819778144 | 1.60E-12 |
| OR8H1 | 0.820036781 | 0.000496218 |
| LIF | 0.820385852 | 0.003805889 |
| CERS1 | 0.821279029 | 3.34E-07 |
| CCL18 | 0.821519789 | 0.000118772 |
| AC079594.2 | 0.821728631 | 0.000362529 |
| PIF1 | 0.821826933 | 2.00E-11 |
| NNMT | 0.822046451 | 6.40E-09 |
| TUBB8P12 | 0.822825619 | 0.001382754 |
| TRIM2 | 0.825776923 | 2.30E-13 |
| BHMT2 | 0.825817839 | 5.98E-08 |
| TTC21A | 0.827132601 | 9.13E-10 |
| GFRA1 | 0.827439546 | 1.29E-08 |
| METTL27 | 0.827842494 | 9.89E-06 |
| SIRPG | 0.829566029 | 2.40E-09 |
| OGN | 0.829585293 | 1.80E-05 |
| FAM237B | 0.829755541 | 2.10E-08 |
| RBFOX1 | 0.829995129 | 3.27E-06 |
| CHODL | 0.831096202 | 1.37E-13 |
| EDARADD | 0.831175364 | 1.50E-10 |
| FRMPD4 | 0.831772725 | 0.00017472 |
| SYTL1 | 0.831916771 | 4.20E-20 |
| PDLIM3 | 0.834395377 | 1.16E-14 |
| IL2RA | 0.834530796 | 3.40E-07 |
| CPT1C | 0.834721583 | 5.98E-16 |
| IQCH | 0.834747748 | 7.00E-08 |
| TMC1 | 0.834919788 | 5.40E-09 |
| GLB1L | 0.835193944 | 4.21E-16 |
| SEC14L2 | 0.835680447 | 2.67E-21 |
| KCNN1 | 0.83573138 | 1.31E-08 |
| SLC12A3 | 0.836326544 | 4.04E-05 |
| NBPF4 | 0.83680343 | 0.029746662 |
| EPPK1 | 0.837610786 | 7.35E-11 |
| AL360181.3 | 0.83762039 | 2.32E-07 |
| TDGF1 | 0.838925663 | 0.000274038 |
| UGT3A2 | 0.839202794 | 0.009650556 |
| CPA5 | 0.839326195 | 0.000399428 |
| PSG4 | 0.839658887 | 0.000608597 |
| RIMBP2 | 0.839956835 | 0.000131871 |
| SLC35E4 | 0.840165335 | 1.44E-07 |
| ENO4 | 0.840815959 | 0.00501044 |
| OXT | 0.840900826 | 2.83E-05 |
| TNFSF14 | 0.841543899 | 2.43E-08 |
| ENTPD1 | 0.841619173 | 2.11E-39 |
| GCLC | 0.841775354 | 1.04E-20 |
| LRRC23 | 0.843369562 | 1.80E-09 |
| ASTN2 | 0.843374045 | 5.80E-18 |
| TSPAN11 | 0.843389222 | 3.45E-09 |
| LEF1 | 0.845047912 | 3.95E-20 |
| ST14 | 0.845895678 | 1.48E-06 |
| ITM2C | 0.846519379 | 5.03E-11 |
| TMEM239 | 0.846738683 | 0.000122069 |
| BTBD11 | 0.846857136 | 1.27E-13 |
| TUBAL3 | 0.847919785 | 0.0441792 |
| ANKRD65 | 0.848959401 | 2.06E-18 |
| DENND6B | 0.849079606 | 1.40E-11 |
| FAM169A | 0.849262205 | 4.93E-05 |
| FAM163B | 0.850041195 | 7.83E-08 |
| ATG9B | 0.850877143 | 3.32E-10 |
| SORCS2 | 0.852199655 | 1.20E-16 |
| NECTIN1 | 0.852872239 | 1.46E-09 |
| NOVA1 | 0.85352374 | 5.24E-11 |
| CADPS | 0.854154608 | 3.08E-12 |
| CCDC86 | 0.854327036 | 7.75E-05 |
| PANX3 | 0.85545445 | 0.036675595 |
| FAM155A | 0.855617199 | 3.43E-13 |
| GRIP2 | 0.856162164 | 2.26E-08 |
| PFN2 | 0.856789856 | 4.62E-19 |
| CLCNKA | 0.857222689 | 9.10E-07 |
| UNC5CL | 0.857693737 | 5.80E-08 |
| OSBPL6 | 0.857960099 | 3.44E-09 |
| FAT1 | 0.858471827 | 6.41E-32 |
| CCDC178 | 0.858477621 | 0.000431178 |
| GZMK | 0.858579825 | 1.23E-08 |
| URB1 | 0.859050679 | 0.001065252 |
| CEP126 | 0.859172137 | 6.42E-11 |
| RAB7B | 0.859320716 | 5.17E-13 |
| SCN3A | 0.859573848 | 8.74E-05 |
| LEKR1 | 0.859619275 | 3.32E-08 |
| SERPINB2 | 0.8596552 | 0.049271518 |
| MKRN2OS | 0.860194176 | 7.38E-17 |
| HAO2 | 0.860332007 | 0.00065362 |
| UNC79 | 0.860592924 | 9.83E-11 |
| IAPP | 0.860862779 | 0.016671442 |
| CYP11A1 | 0.861235493 | 1.42E-09 |
| ACTA2 | 0.861275362 | 2.17E-10 |
| BBOF1 | 0.86241214 | 1.59E-10 |
| SYNGAP1 | 0.863002227 | 3.35E-14 |
| FAM177B | 0.863568339 | 1.09E-06 |
| WDR78 | 0.863892177 | 5.39E-09 |
| AP002748.4 | 0.863932202 | 0.006090934 |
| EDA2R | 0.864037376 | 3.89E-09 |
| PLAU | 0.864329612 | 3.30E-09 |
| CCL21 | 0.864554344 | 2.75E-07 |
| NMU | 0.864648139 | 0.000408606 |
| C1QTNF9B | 0.864872128 | 7.37E-10 |
| COL9A1 | 0.865023166 | 3.99E-06 |
| VIPR2 | 0.866234079 | 9.34E-05 |
| SELE | 0.867638384 | 0.018328926 |
| PROS1 | 0.868301417 | 0.003542542 |
| HIF1A | 0.868732195 | 3.33E-12 |
| CCR6 | 0.869054134 | 1.26E-08 |
| BLOC1S5-TXNDC5 | 0.870812115 | 1.28E-05 |
| PLIN1 | 0.871135178 | 0.000340131 |
| COL18A1 | 0.871616275 | 2.47E-19 |
| GLYATL1B | 0.871989253 | 0.032791091 |
| GRIK5 | 0.872229404 | 4.94E-12 |
| ALDH18A1 | 0.872364015 | 9.40E-23 |
| PPFIA2 | 0.872437139 | 1.98E-14 |
| EPHA3 | 0.872626055 | 9.78E-06 |
| ANLN | 0.872663178 | 6.31E-07 |
| SPRY1 | 0.874029776 | 1.87E-16 |
| CD180 | 0.87408913 | 6.20E-09 |
| KCNK9 | 0.874758545 | 0.004678438 |
| TMEM132C | 0.874864814 | 7.79E-08 |
| CPA6 | 0.875135461 | 9.07E-05 |
| KCNK4 | 0.875529799 | 0.000288409 |
| PLA2G7 | 0.875925533 | 2.05E-06 |
| NMNAT2 | 0.876148554 | 2.07E-11 |
| LRTOMT | 0.87644632 | 5.66E-11 |
| TPPP3 | 0.876648355 | 1.20E-07 |
| KIF5A | 0.876819915 | 0.032619885 |
| RHOD | 0.877094274 | 1.10E-10 |
| UBE2U | 0.877946085 | 0.005306979 |
| CCDC198 | 0.878081119 | 4.44E-05 |
| PLPP5 | 0.878707638 | 5.03E-60 |
| COL16A1 | 0.878864702 | 4.58E-24 |
| FOXD4L3 | 0.879342716 | 0.00288341 |
| SEC14L3 | 0.879497577 | 0.000867603 |
| FBXW9 | 0.879893406 | 2.05E-09 |
| NMBR | 0.880048259 | 2.63E-05 |
| DMBX1 | 0.880509095 | 3.62E-05 |
| SEL1L3 | 0.880730895 | 3.51E-37 |
| CLCN1 | 0.881833128 | 7.33E-09 |
| TTC16 | 0.884085184 | 3.11E-06 |
| IQCA1 | 0.885512501 | 9.48E-11 |
| MAJIN | 0.885593274 | 2.77E-07 |
| UTS2 | 0.885905304 | 8.22E-05 |
| ETNPPL | 0.886615348 | 0.002305331 |
| IL1RN | 0.886673984 | 9.38E-06 |
| F5 | 0.887197033 | 1.02E-09 |
| PTGES3L | 0.888253562 | 7.40E-22 |
| MRO | 0.888578297 | 7.84E-09 |
| CCN4 | 0.888956771 | 0.007142299 |
| KIF6 | 0.889495734 | 4.56E-09 |
| ADRA2A | 0.889630546 | 1.87E-10 |
| TMEM63C | 0.889648855 | 0.000274038 |
| TTC26 | 0.890323799 | 5.82E-12 |
| VCAN | 0.891976392 | 1.30E-12 |
| SYK | 0.892814984 | 1.71E-09 |
| TEX55 | 0.892901954 | 0.026921801 |
| GPR37 | 0.893653877 | 3.57E-09 |
| TF | 0.894766878 | 2.58E-10 |
| MYH11 | 0.894769342 | 3.78E-11 |
| KIF7 | 0.894973803 | 1.43E-09 |
| CLECL1 | 0.895305503 | 1.71E-09 |
| UCHL1 | 0.896046431 | 9.05E-09 |
| CCL14 | 0.896771673 | 3.12E-07 |
| AMY1B | 0.897437972 | 7.81E-05 |
| ANKDD1B | 0.897825204 | 6.69E-12 |
| PGM2L1 | 0.898130689 | 0.008294032 |
| AC093512.2 | 0.898199409 | 5.90E-07 |
| C1QTNF9 | 0.898364525 | 5.87E-07 |
| MESP1 | 0.898802709 | 4.57E-12 |
| SDR42E2 | 0.898805933 | 6.06E-06 |
| NPHP1 | 0.898988072 | 2.83E-13 |
| DRP2 | 0.899078856 | 8.14E-11 |
| CCKBR | 0.900455094 | 0.015886746 |
| WDR54 | 0.900673097 | 1.07E-12 |
| NCS1 | 0.900680557 | 1.32E-06 |
| P2RY6 | 0.901265928 | 7.17E-13 |
| SCARA3 | 0.90139492 | 1.62E-15 |
| MYT1 | 0.901497715 | 0.000257042 |
| NTS | 0.902579277 | 0.015557381 |
| MYOCD | 0.902671363 | 1.30E-10 |
| MAEL | 0.903326076 | 6.43E-09 |
| PERM1 | 0.90341109 | 9.21E-22 |
| CFHR1 | 0.903855043 | 4.76E-10 |
| OR2AG2 | 0.903863204 | 5.39E-09 |
| TLL2 | 0.904627068 | 9.21E-07 |
| PKP1 | 0.904877659 | 1.01E-07 |
| CPZ | 0.904902501 | 2.56E-08 |
| LOXL1 | 0.905181292 | 3.89E-18 |
| HTR4 | 0.905184702 | 3.43E-10 |
| MIA | 0.905322556 | 2.49E-07 |
| CCDC157 | 0.905560696 | 9.62E-12 |
| FSHR | 0.906120061 | 8.80E-05 |
| PLD5 | 0.906799535 | 2.75E-07 |
| RGS1 | 0.907098146 | 3.62E-05 |
| CYGB | 0.907147383 | 1.06E-20 |
| SLC35F2 | 0.908935409 | 0.000240202 |
| AC053503.7 | 0.909972858 | 0.000174702 |
| CFAP54 | 0.910251829 | 8.59E-14 |
| SYT6 | 0.910681101 | 8.33E-07 |
| SPEG | 0.911672846 | 8.49E-13 |
| BGN | 0.912746249 | 2.30E-07 |
| GPX8 | 0.913288003 | 2.97E-20 |
| RTL1 | 0.913818174 | 0.005323922 |
| RABL2A | 0.913928158 | 6.51E-13 |
| CALML4 | 0.914497726 | 4.96E-15 |
| LRRC4 | 0.914868771 | 4.86E-07 |
| SLAMF1 | 0.916502272 | 4.54E-06 |
| KATNAL2 | 0.916852964 | 5.37E-17 |
| CNIH2 | 0.919236574 | 8.07E-10 |
| ATP2B3 | 0.919450325 | 2.32E-05 |
| SLC29A4 | 0.91964765 | 5.18E-19 |
| CEACAM6 | 0.91991143 | 3.42E-08 |
| TTLL6 | 0.920504313 | 1.56E-06 |
| NAT8L | 0.920636079 | 2.47E-08 |
| CCL17 | 0.920681939 | 2.03E-06 |
| KIRREL2 | 0.920687132 | 4.82E-05 |
| SYNJ2 | 0.922048858 | 7.68E-15 |
| BICDL1 | 0.922285074 | 1.49E-14 |
| TMEM155 | 0.922353336 | 3.39E-08 |
| GSC | 0.922672212 | 5.53E-07 |
| NPTX2 | 0.923275328 | 0.037690947 |
| AL139260.3 | 0.92424885 | 3.91E-09 |
| MYLPF | 0.925160686 | 2.76E-14 |
| LYPD6B | 0.926696494 | 7.31E-07 |
| CEP128 | 0.926931974 | 5.75E-19 |
| AL353579.1 | 0.927272263 | 0.003392981 |
| MINAR1 | 0.927555954 | 6.00E-27 |
| PRRT3 | 0.927599265 | 1.86E-10 |
| PROC | 0.927693624 | 2.60E-22 |
| SPSB1 | 0.928084581 | 0.000875088 |
| C7 | 0.928449407 | 3.43E-13 |
| CCDC24 | 0.928505585 | 1.18E-15 |
| CXorf58 | 0.928665279 | 1.55E-07 |
| SLC22A4 | 0.929251372 | 1.28E-07 |
| VWA5B2 | 0.929288634 | 3.49E-10 |
| FAM20C | 0.929581493 | 3.38E-14 |
| SLAMF9 | 0.929874495 | 0.000256315 |
| FBN1 | 0.929905939 | 1.46E-17 |
| F12 | 0.93057998 | 1.44E-25 |
| FNDC11 | 0.930605471 | 3.74E-09 |
| ALLC | 0.930734467 | 0.036590496 |
| C16orf74 | 0.930746027 | 8.00E-24 |
| F2 | 0.931330061 | 0.001178456 |
| COL9A2 | 0.935806958 | 1.40E-08 |
| NFATC2 | 0.935990599 | 4.65E-08 |
| SVOPL | 0.936226403 | 7.85E-09 |
| HPCA | 0.9371053 | 6.25E-07 |
| LINGO1 | 0.937663616 | 2.32E-09 |
| OSCP1 | 0.937836651 | 2.22E-11 |
| PHF24 | 0.938368143 | 4.54E-12 |
| CLGN | 0.938743378 | 2.09E-05 |
| KRT80 | 0.938910014 | 4.37E-09 |
| DPF1 | 0.941110789 | 4.97E-19 |
| CCDC138 | 0.941236814 | 1.16E-11 |
| AK7 | 0.941243269 | 2.63E-09 |
| AC034228.4 | 0.942159081 | 0.009515406 |
| PNMA8C | 0.942283561 | 2.10E-06 |
| MKX | 0.942596176 | 0.000466313 |
| ADAMTSL1 | 0.943489313 | 2.23E-11 |
| KLHL32 | 0.94360454 | 0.000771173 |
| LMOD1 | 0.943836676 | 0.003130246 |
| GRM5 | 0.944755883 | 0.001407316 |
| ATP10B | 0.945059119 | 2.87E-10 |
| SNTG2 | 0.945159679 | 2.08E-09 |
| ISLR2 | 0.945294457 | 1.37E-11 |
| TEKT5 | 0.945600223 | 1.12E-11 |
| SRGAP3 | 0.945951833 | 6.73E-18 |
| ZMYND12 | 0.946709399 | 1.88E-10 |
| TEX36 | 0.947075517 | 0.015049041 |
| PHF21B | 0.949420608 | 6.06E-09 |
| PGF | 0.950344175 | 2.50E-11 |
| HAGHL | 0.950863255 | 1.02E-15 |
| EGFLAM | 0.951941423 | 1.61E-11 |
| NR0B1 | 0.95230385 | 0.01883686 |
| PRG4 | 0.952518662 | 1.12E-06 |
| FLRT2 | 0.953062922 | 3.09E-15 |
| C16orf46 | 0.953244622 | 1.62E-10 |
| FAM71F1 | 0.953784511 | 8.49E-05 |
| GPR156 | 0.954067111 | 9.75E-09 |
| GADL1 | 0.954260898 | 1.28E-21 |
| MAP6 | 0.955188041 | 3.52E-11 |
| B9D1 | 0.955234361 | 1.80E-15 |
| REEP6 | 0.955351462 | 0.036228314 |
| TNFRSF21 | 0.955524184 | 3.05E-24 |
| INAVA | 0.955532773 | 7.97E-20 |
| SHOC1 | 0.95677423 | 3.63E-13 |
| HOXD8 | 0.956932826 | 7.50E-10 |
| LRRC15 | 0.957669021 | 3.92E-06 |
| POU3F1 | 0.958259527 | 5.25E-07 |
| CEACAM5 | 0.958598104 | 5.14E-05 |
| LARGE2 | 0.958895448 | 9.44E-22 |
| CLDN3 | 0.959234816 | 2.53E-11 |
| ALG1L | 0.959446094 | 1.38E-05 |
| CETN2 | 0.95956609 | 2.87E-14 |
| TCTEX1D2 | 0.959689751 | 2.41E-14 |
| ALDH1L2 | 0.959908818 | 2.14E-16 |
| CYP2C8 | 0.959926457 | 1.20E-09 |
| CYP2A6 | 0.960436215 | 3.22E-06 |
| HOXD10 | 0.960759288 | 7.06E-05 |
| AL353588.1 | 0.960885205 | 2.07E-10 |
| P3H3 | 0.963376421 | 1.54E-21 |
| CHRNA6 | 0.96373573 | 2.26E-05 |
| ELL3 | 0.963894758 | 9.11E-17 |
| PPP1R36 | 0.963986748 | 5.82E-07 |
| CTNNA2 | 0.964775744 | 4.06E-07 |
| GASK1A | 0.964908805 | 7.56E-14 |
| SLC28A1 | 0.965804038 | 3.22E-06 |
| RAB30 | 0.969775984 | 5.12E-17 |
| BAIAP3 | 0.970885001 | 1.62E-08 |
| PDE1A | 0.970977418 | 1.47E-20 |
| GRIA2 | 0.973197261 | 0.002299247 |
| TGFB3 | 0.973275746 | 1.79E-16 |
| SNAP91 | 0.973863939 | 5.61E-06 |
| HDC | 0.974419805 | 4.24E-10 |
| PRR29 | 0.974972913 | 2.21E-08 |
| KRT39 | 0.975106527 | 6.32E-07 |
| OR2I1P | 0.975221672 | 5.85E-07 |
| HMCN2 | 0.975476637 | 1.91E-09 |
| CLDN10 | 0.978282581 | 0.000477567 |
| SSR4 | 0.97927394 | 2.10E-30 |
| ZBTB9 | 0.979755983 | 0.001803722 |
| C6orf52 | 0.98086768 | 6.16E-10 |
| PRR7 | 0.980926175 | 1.48E-12 |
| NHS | 0.98138898 | 7.50E-23 |
| HIST2H3C | 0.982048219 | 8.19E-07 |
| HIST2H3A | 0.982048219 | 8.19E-07 |
| SLC17A9 | 0.982320588 | 2.62E-21 |
| CFAP69 | 0.982540347 | 9.53E-17 |
| HSPA4L | 0.982606006 | 1.06E-17 |
| ZBTB32 | 0.982783431 | 7.92E-13 |
| PRSS22 | 0.982967484 | 8.37E-14 |
| TMEM213 | 0.983202203 | 0.002957461 |
| CERCAM | 0.983674069 | 1.58E-18 |
| SLITRK3 | 0.986132471 | 5.35E-05 |
| CALB2 | 0.986410021 | 0.000749767 |
| TMEM52 | 0.98803468 | 8.48E-07 |
| CXCL12 | 0.98823944 | 1.64E-17 |
| ENPP6 | 0.989634934 | 0.001236221 |
| RFX8 | 0.989683459 | 4.48E-12 |
| IL31RA | 0.990288622 | 1.58E-08 |
| C2CD4A | 0.991374319 | 6.11E-05 |
| GGT6 | 0.991446868 | 4.30E-13 |
| CASKIN1 | 0.992782789 | 0.001703143 |
| SMIM22 | 0.993226086 | 1.65E-13 |
| PRDX4 | 0.993237577 | 6.14E-09 |
| NTRK1 | 0.994430541 | 6.09E-15 |
| DZIP1L | 0.994813415 | 3.33E-13 |
| NXF2 | 0.995446332 | 0.000208092 |
| SMIM35 | 0.995897792 | 1.25E-10 |
| HES2 | 0.996184161 | 1.66E-12 |
| ATP2B2 | 0.997174306 | 0.042746443 |
| ATP1A2 | 0.999527107 | 6.11E-10 |
| CGB7 | 0.999606406 | 6.29E-11 |
| PRMT8 | 0.999625655 | 4.40E-07 |
| RIMS2 | 1.000148286 | 1.59E-15 |
| OPCML | 1.000254825 | 4.28E-05 |
| RNF207 | 1.000274457 | 1.54E-20 |
| SPRED3 | 1.001729566 | 8.59E-09 |
| UBE3D | 1.002727238 | 1.80E-11 |
| HIST1H3G | 1.003039766 | 5.81E-08 |
| COL28A1 | 1.003154218 | 4.29E-09 |
| TNC | 1.003265567 | 2.93E-09 |
| GGT5 | 1.003640451 | 3.44E-16 |
| LY6H | 1.003907671 | 2.69E-06 |
| LCA5L | 1.005242175 | 6.72E-12 |
| PNMA8A | 1.005724451 | 0.022226892 |
| MT3 | 1.006164661 | 0.000805109 |
| CCDC81 | 1.006216069 | 6.20E-11 |
| SMOC2 | 1.007520053 | 2.48E-12 |
| OR1Q1 | 1.007747268 | 0.018301368 |
| NFATC4 | 1.008848955 | 2.44E-31 |
| CRYM | 1.009332697 | 6.82E-14 |
| S100A11 | 1.00988553 | 7.02E-09 |
| FEV | 1.010750866 | 0.006730154 |
| KLHDC7A | 1.012429596 | 1.54E-12 |
| GJB6 | 1.012627027 | 0.000918435 |
| CDX1 | 1.012844377 | 9.49E-13 |
| TERT | 1.013210262 | 0.00325554 |
| LXN | 1.014194448 | 3.14E-19 |
| ADAMTS9 | 1.014336892 | 2.16E-09 |
| OAF | 1.014944985 | 8.44E-13 |
| C12orf75 | 1.015185486 | 3.53E-13 |
| KLK8 | 1.015890542 | 0.019989702 |
| PDE7B | 1.016112337 | 8.79E-17 |
| GSTA3 | 1.016131137 | 0.003733466 |
| LRRC6 | 1.017299055 | 3.18E-14 |
| STEAP3 | 1.017422241 | 1.25E-21 |
| VCX3B | 1.017504761 | 0.008274408 |
| MDK | 1.018937429 | 8.50E-20 |
| GREB1 | 1.019056519 | 4.12E-17 |
| AHNAK2 | 1.019245517 | 1.08E-13 |
| HEPN1 | 1.019525152 | 0.040478307 |
| CAPN5 | 1.019691864 | 1.34E-13 |
| HECW1 | 1.020399288 | 4.87E-11 |
| PCDH19 | 1.020530059 | 1.23E-07 |
| VWA7 | 1.021028611 | 2.03E-14 |
| EFCAB12 | 1.021474026 | 3.20E-09 |
| ELFN1 | 1.022386871 | 1.15E-13 |
| TAT | 1.022457412 | 0.004839349 |
| MAK | 1.02290651 | 1.44E-10 |
| C10orf95 | 1.023420042 | 3.30E-09 |
| LGALS9B | 1.023782099 | 1.08E-06 |
| LRRC34 | 1.024649962 | 2.93E-12 |
| UGT2A1 | 1.024991685 | 0.000604286 |
| NEK11 | 1.02583557 | 1.03E-14 |
| C9orf50 | 1.026012378 | 2.41E-15 |
| DPP10 | 1.026491221 | 2.03E-14 |
| ZSCAN1 | 1.02651173 | 9.22E-08 |
| CFI | 1.026888702 | 8.73E-42 |
| WDR86 | 1.027106932 | 1.24E-16 |
| FKBP6 | 1.027417245 | 5.26E-11 |
| ITLN1 | 1.027577243 | 0.014382465 |
| KRT16 | 1.031688984 | 0.002023214 |
| AC005041.1 | 1.032259828 | 7.94E-13 |
| IQANK1 | 1.032691887 | 2.34E-13 |
| PODNL1 | 1.033463141 | 1.57E-35 |
| SCN4A | 1.034734439 | 1.32E-08 |
| OSTN | 1.035291453 | 7.04E-06 |
| USP2 | 1.035625138 | 6.21E-16 |
| GRIN2A | 1.037439381 | 4.97E-07 |
| CRISPLD1 | 1.037632406 | 8.52E-14 |
| PRSS3 | 1.038098381 | 7.95E-06 |
| GLB1L2 | 1.038231342 | 7.74E-18 |
| GPR162 | 1.038739712 | 1.35E-16 |
| TIGD4 | 1.039486262 | 1.22E-09 |
| MEX3A | 1.039733167 | 0.021482593 |
| ZNF273 | 1.039758884 | 1.49E-11 |
| KPNA7 | 1.041359798 | 1.40E-10 |
| TXNDC5 | 1.041549021 | 8.87E-34 |
| SLC2A4 | 1.041591189 | 2.57E-12 |
| NME5 | 1.043693909 | 2.20E-09 |
| SYNPO2 | 1.044122812 | 5.95E-16 |
| PKP2 | 1.044493124 | 1.76E-10 |
| MAATS1 | 1.044812594 | 8.31E-10 |
| CCDC170 | 1.04632282 | 0.001748165 |
| SYT5 | 1.047508259 | 6.80E-05 |
| GNB3 | 1.048303373 | 5.38E-21 |
| NBEA | 1.048655451 | 2.26E-18 |
| PPIL6 | 1.049385874 | 1.20E-09 |
| PTGES3L-AARSD1 | 1.049458207 | 2.09E-14 |
| ZFHX2 | 1.049509674 | 8.36E-11 |
| SULF2 | 1.050168706 | 8.47E-29 |
| SLC2A7 | 1.050316078 | 0.002653998 |
| SBK2 | 1.050797271 | 0.01915759 |
| WIPF3 | 1.051986083 | 1.04E-11 |
| TMPRSS7 | 1.052586386 | 5.86E-09 |
| TEX9 | 1.053333015 | 1.51E-18 |
| PPP1R14D | 1.053338431 | 2.82E-13 |
| LDB3 | 1.053486067 | 1.85E-14 |
| DPYSL4 | 1.053711297 | 6.68E-05 |
| ELMOD1 | 1.054387812 | 8.23E-08 |
| SHISA6 | 1.055212291 | 3.21E-09 |
| ZPBP | 1.055315312 | 0.000413165 |
| DSP | 1.055449113 | 3.03E-16 |
| TMEM232 | 1.056048499 | 1.84E-09 |
| KLHL14 | 1.056050609 | 1.31E-14 |
| PC | 1.056546987 | 2.05E-08 |
| TRIM17 | 1.056683697 | 7.93E-19 |
| TMEM238L | 1.057374424 | 0.000148154 |
| CCDC180 | 1.05764346 | 4.04E-12 |
| TIMP1 | 1.058666855 | 6.06E-08 |
| FGF14 | 1.058978906 | 7.10E-13 |
| CERKL | 1.059214463 | 8.47E-07 |
| GEM | 1.059231351 | 4.64E-14 |
| KIAA1211L | 1.060343002 | 2.16E-08 |
| OR6C6 | 1.06140783 | 1.69E-05 |
| MND1 | 1.063519665 | 7.26E-17 |
| AGBL4 | 1.063795671 | 6.11E-06 |
| CDKN2A | 1.064373234 | 3.75E-18 |
| DNALI1 | 1.06450506 | 8.72E-15 |
| PLK1 | 1.064852691 | 1.18E-16 |
| CCDC184 | 1.064909574 | 4.13E-15 |
| WFDC2 | 1.065195096 | 7.76E-13 |
| FAM229B | 1.066594681 | 3.42E-17 |
| DIO1 | 1.067716167 | 1.11E-11 |
| RTL3 | 1.068097406 | 2.32E-10 |
| WDR64 | 1.069123451 | 1.99E-08 |
| MAGEC3 | 1.069238813 | 1.79E-07 |
| FP565260.4 | 1.06961255 | 0.002321037 |
| GRIK2 | 1.070231199 | 8.22E-13 |
| CABYR | 1.071234226 | 1.94E-30 |
| FGFBP1 | 1.071378581 | 4.74E-06 |
| CACNA1B | 1.071398387 | 3.43E-06 |
| C1orf185 | 1.07152733 | 0.03690613 |
| AC131097.2 | 1.073138981 | 2.44E-07 |
| ADH4 | 1.073634528 | 1.28E-12 |
| TM4SF19-TCTEX1D2 | 1.074114415 | 0.000179067 |
| WHRN | 1.074189532 | 9.88E-19 |
| TMEM40 | 1.074589988 | 3.55E-10 |
| PRSS54 | 1.076497946 | 0.003145365 |
| ALPK2 | 1.076994066 | 1.48E-08 |
| ABCB4 | 1.079558548 | 6.45E-11 |
| CDCA7 | 1.080577433 | 2.70E-16 |
| GNAO1 | 1.081162921 | 9.49E-26 |
| LRRIQ3 | 1.081216344 | 6.24E-09 |
| CCDC148 | 1.081788599 | 6.91E-11 |
| JPT2 | 1.082660398 | 1.07E-12 |
| GSDME | 1.083724639 | 5.50E-21 |
| AC073508.2 | 1.084706245 | 3.68E-05 |
| NRG4 | 1.0851689 | 3.85E-11 |
| CES4A | 1.085796274 | 2.09E-14 |
| TDRD5 | 1.085999377 | 2.79E-07 |
| VSTM2L | 1.087903196 | 1.33E-11 |
| PKIB | 1.088576373 | 1.55E-14 |
| ARNTL2 | 1.089612448 | 2.77E-15 |
| FCER2 | 1.091320343 | 7.30E-09 |
| NPHS1 | 1.091517109 | 2.26E-05 |
| SDS | 1.092504931 | 2.13E-09 |
| LRIT2 | 1.092617603 | 0.006616327 |
| LRRC9 | 1.092726541 | 1.56E-05 |
| FBXL22 | 1.092970657 | 4.65E-14 |
| NELL2 | 1.094759336 | 8.00E-10 |
| NR4A3 | 1.095575964 | 5.89E-06 |
| ARMH2 | 1.095961379 | 1.40E-05 |
| MUC21 | 1.096407909 | 7.20E-07 |
| RGMA | 1.096590687 | 7.09E-16 |
| MOXD1 | 1.098379357 | 4.15E-09 |
| SMO | 1.098596055 | 7.19E-13 |
| REEP2 | 1.09871645 | 2.04E-22 |
| CCL15-CCL14 | 1.098988611 | 1.84E-12 |
| GLB1L3 | 1.100879139 | 4.45E-26 |
| TCTEX1D1 | 1.100884266 | 3.02E-08 |
| SMIM6 | 1.101001859 | 4.31E-12 |
| ALS2CR12 | 1.101386071 | 5.49E-10 |
| C4orf54 | 1.102391637 | 2.47E-05 |
| TNFRSF18 | 1.10247457 | 3.89E-16 |
| EN1 | 1.103848532 | 0.005862369 |
| TRPV3 | 1.106013786 | 1.04E-18 |
| IL24 | 1.106448676 | 1.92E-12 |
| HRC | 1.107437617 | 5.50E-05 |
| SLC7A5 | 1.10855155 | 3.40E-10 |
| AMY1A | 1.108623093 | 1.42E-06 |
| LAX1 | 1.109244436 | 4.21E-23 |
| NECTIN4 | 1.109943206 | 6.83E-13 |
| RHBDL2 | 1.111645389 | 3.15E-25 |
| TTC25 | 1.113956436 | 1.13E-10 |
| DHRS9 | 1.11462357 | 1.38E-11 |
| CAMKV | 1.114832923 | 8.39E-05 |
| MYEOV | 1.114854962 | 2.17E-17 |
| UBD | 1.117054989 | 3.63E-09 |
| PRAME | 1.117556175 | 0.000258938 |
| STEAP2 | 1.117889704 | 3.14E-19 |
| AL355987.1 | 1.118706917 | 0.000184927 |
| MUC20 | 1.118797107 | 1.02E-23 |
| CYTL1 | 1.120025943 | 0.015652371 |
| LRRC4C | 1.120650244 | 7.71E-22 |
| BRSK1 | 1.120727524 | 7.36E-33 |
| ISM2 | 1.121341044 | 0.001754108 |
| BCL2L15 | 1.122837601 | 1.83E-17 |
| HEPACAM2 | 1.123102035 | 2.39E-06 |
| ZSWIM2 | 1.123663274 | 0.007967797 |
| NAT1 | 1.124330153 | 1.84E-13 |
| MEPE | 1.12722032 | 0.000356282 |
| SLC4A3 | 1.127773351 | 2.50E-29 |
| CCDC40 | 1.129577604 | 4.54E-15 |
| AC009336.2 | 1.129906147 | 1.70E-06 |
| GNAL | 1.129989578 | 3.50E-10 |
| EFCAB6 | 1.130889497 | 3.41E-11 |
| SLAMF7 | 1.132038211 | 7.18E-14 |
| LRRC61 | 1.132168141 | 3.61E-05 |
| ST6GAL1 | 1.132197177 | 6.70E-43 |
| LTBP1 | 1.133172816 | 2.27E-33 |
| ATP1A4 | 1.13376942 | 4.93E-14 |
| AMTN | 1.133836349 | 0.015674567 |
| C8orf89 | 1.134210672 | 7.63E-09 |
| ITGA11 | 1.134854993 | 2.15E-18 |
| NT5C1A | 1.135388356 | 3.47E-06 |
| RALGPS2 | 1.135539237 | 2.04E-30 |
| DLK2 | 1.135584316 | 9.36E-17 |
| STK33 | 1.135929201 | 3.95E-13 |
| BHMG1 | 1.136223534 | 0.001576429 |
| LY9 | 1.13688075 | 1.95E-13 |
| CRLF2 | 1.138756082 | 1.05E-13 |
| NXPE1 | 1.13892775 | 5.97E-07 |
| CER1 | 1.139601582 | 0.000294069 |
| NPBWR1 | 1.140227541 | 0.00031437 |
| CXorf49B | 1.140408462 | 5.39E-11 |
| CXorf49 | 1.140408462 | 5.39E-11 |
| INSL5 | 1.141095627 | 8.49E-06 |
| BRINP3 | 1.143134025 | 6.56E-11 |
| DLX5 | 1.143529458 | 3.58E-05 |
| CFHR3 | 1.144762683 | 9.09E-14 |
| CDH26 | 1.145417397 | 9.63E-20 |
| SRCIN1 | 1.145490689 | 1.26E-12 |
| RBM46 | 1.145512075 | 0.030586783 |
| COL15A1 | 1.146489486 | 3.54E-21 |
| C5orf46 | 1.146913612 | 1.15E-05 |
| TACR1 | 1.147161636 | 2.49E-05 |
| CNGA1 | 1.147744136 | 1.97E-17 |
| SERPINF1 | 1.148735735 | 2.08E-27 |
| APCDD1L | 1.148963303 | 1.82E-07 |
| TMEM184A | 1.150552681 | 1.32E-16 |
| FAM129C | 1.150745324 | 2.33E-14 |
| CCDC13 | 1.152726596 | 1.03E-14 |
| MROH2A | 1.152766793 | 2.33E-05 |
| GAD1 | 1.155915758 | 5.74E-10 |
| GIPR | 1.156279228 | 8.65E-13 |
| AC018512.1 | 1.156596735 | 6.70E-06 |
| SLC13A5 | 1.156848919 | 2.05E-14 |
| KIAA1324 | 1.157483025 | 3.06E-12 |
| ROR2 | 1.157684773 | 8.17E-13 |
| LRRC18 | 1.159847345 | 3.94E-10 |
| CHIT1 | 1.160586168 | 4.46E-05 |
| DHRS7C | 1.161488124 | 0.045196975 |
| CA9 | 1.162914004 | 1.43E-11 |
| MELTF | 1.163613069 | 1.90E-21 |
| AP000812.5 | 1.164256825 | 1.01E-08 |
| GRIA3 | 1.164482304 | 5.04E-13 |
| MTTP | 1.165999356 | 6.91E-11 |
| ADAMTS19 | 1.166837005 | 4.41E-05 |
| PDK1 | 1.167065834 | 5.27E-30 |
| COL9A3 | 1.16852483 | 1.04E-13 |
| LHCGR | 1.169486089 | 6.18E-07 |
| CCDC153 | 1.169702129 | 5.06E-13 |
| MMP2 | 1.170843451 | 4.36E-22 |
| PXDNL | 1.173199002 | 0.000215675 |
| KCTD1 | 1.173655089 | 4.37E-30 |
| NPW | 1.174820561 | 5.84E-06 |
| NKAIN2 | 1.174899866 | 8.85E-08 |
| GPX2 | 1.174992589 | 5.01E-18 |
| SMIM34A | 1.175370273 | 1.82E-10 |
| SMIM34B | 1.175370273 | 1.82E-10 |
| CALCA | 1.175693849 | 2.30E-05 |
| NPHS2 | 1.17578141 | 0.018677132 |
| OSR1 | 1.176732571 | 5.72E-15 |
| CLDN19 | 1.177053577 | 2.67E-06 |
| B4GALNT4 | 1.1776418 | 4.59E-09 |
| MRVI1 | 1.177894328 | 1.07E-29 |
| NSG1 | 1.178068204 | 1.48E-27 |
| FAIM2 | 1.178352385 | 9.75E-09 |
| RUNDC3B | 1.178455142 | 1.55E-11 |
| TLR10 | 1.180661139 | 6.38E-11 |
| KCND3 | 1.181267874 | 3.21E-33 |
| C2orf66 | 1.183224503 | 8.78E-08 |
| GALNT9 | 1.184384897 | 1.30E-10 |
| RHBDL1 | 1.184662003 | 4.51E-44 |
| IL5RA | 1.186026977 | 5.04E-13 |
| TMPRSS11D | 1.18616746 | 0.000357818 |
| FST | 1.186179276 | 1.37E-16 |
| WDR93 | 1.186747288 | 2.40E-10 |
| FABP6 | 1.187390067 | 3.13E-06 |
| TENM4 | 1.187727418 | 4.42E-28 |
| CLU | 1.188519136 | 4.93E-23 |
| CPA4 | 1.189428486 | 1.46E-05 |
| DNMT3L | 1.189796979 | 0.00134317 |
| PZP | 1.190114277 | 2.15E-09 |
| CLEC20A | 1.190283396 | 5.06E-08 |
| SFRP4 | 1.191751097 | 1.16E-11 |
| ROPN1B | 1.193371465 | 2.81E-08 |
| TAC1 | 1.195344785 | 0.002188015 |
| SFRP1 | 1.195810434 | 3.39E-07 |
| C17orf97 | 1.195894108 | 2.15E-07 |
| ELOA2 | 1.196403548 | 0.000471003 |
| CYP21A2 | 1.198358172 | 5.42E-25 |
| PRB2 | 1.200412506 | 0.0076083 |
| HS3ST1 | 1.201221456 | 3.32E-11 |
| EFHC1 | 1.202056628 | 8.38E-18 |
| EML6 | 1.202391196 | 7.19E-27 |
| MFAP2 | 1.203104398 | 3.47E-23 |
| BMPR1B | 1.204594944 | 1.30E-11 |
| ITIH1 | 1.20518016 | 4.36E-23 |
| COL25A1 | 1.206453151 | 1.63E-10 |
| CLDN8 | 1.206705186 | 2.56E-05 |
| TMEM244 | 1.206817046 | 0.000156754 |
| IL20 | 1.207284178 | 0.000166196 |
| CLDN1 | 1.210016613 | 8.88E-07 |
| NRXN2 | 1.210375642 | 1.55E-13 |
| IL19 | 1.211288866 | 6.27E-11 |
| KLHDC8A | 1.211925703 | 7.00E-14 |
| IQUB | 1.212133262 | 1.66E-09 |
| GRIN1 | 1.214548354 | 1.60E-10 |
| MNS1 | 1.214580185 | 4.63E-06 |
| OR8G5 | 1.215364833 | 6.88E-05 |
| GP2 | 1.216168365 | 9.61E-08 |
| NUP62CL | 1.219147098 | 1.28E-13 |
| BACE2 | 1.220850273 | 4.12E-47 |
| HMGB3 | 1.223539548 | 5.26E-25 |
| CCL19 | 1.223859655 | 0.002165071 |
| CDHR1 | 1.224275831 | 7.55E-17 |
| LMO1 | 1.225098138 | 2.83E-05 |
| CCDC181 | 1.225552411 | 2.71E-11 |
| MYO16 | 1.225694284 | 3.62E-06 |
| KRT33B | 1.225703102 | 0.012755182 |
| TFF3 | 1.228428575 | 1.61E-09 |
| PTPRN | 1.228438287 | 4.34E-08 |
| CHI3L1 | 1.228702564 | 2.62E-12 |
| CCDC189 | 1.22877773 | 4.96E-15 |
| PTCHD4 | 1.229902292 | 1.63E-20 |
| KIF1A | 1.230290746 | 8.91E-06 |
| HIST3H2BB | 1.231415883 | 6.38E-10 |
| AC073611.1 | 1.23200286 | 2.87E-05 |
| LNP1 | 1.233738846 | 0.035716967 |
| MS4A18 | 1.234255108 | 0.000352534 |
| WSCD2 | 1.234471159 | 3.09E-14 |
| RP1 | 1.234538573 | 2.87E-08 |
| MUC12 | 1.236166341 | 8.40E-26 |
| TMEM88B | 1.236571428 | 7.41E-06 |
| AMBN | 1.237326763 | 0.002459816 |
| STAR | 1.237412221 | 2.70E-19 |
| C9orf116 | 1.238183868 | 1.37E-15 |
| SUSD4 | 1.238338796 | 2.46E-21 |
| PLPPR4 | 1.239554373 | 3.82E-19 |
| VWC2L | 1.240067056 | 4.97E-05 |
| MORN3 | 1.240309463 | 8.53E-13 |
| SLC44A5 | 1.240829509 | 1.27E-08 |
| CABCOCO1 | 1.241029845 | 6.10E-10 |
| KRTAP13-4 | 1.241285952 | 0.012974551 |
| SOX21 | 1.241645948 | 1.47E-05 |
| FKBP11 | 1.241923855 | 2.03E-31 |
| EPHB2 | 1.242475254 | 1.30E-29 |
| CBLC | 1.242556064 | 1.37E-20 |
| LAMP5 | 1.243554274 | 3.46E-05 |
| IGFBP2 | 1.243739722 | 2.47E-27 |
| FOXC1 | 1.248704275 | 4.05E-19 |
| GAL3ST2 | 1.248993124 | 1.11E-06 |
| MRAP | 1.249127328 | 5.36E-09 |
| FBXO16 | 1.249564218 | 7.30E-33 |
| MEI1 | 1.249586309 | 3.13E-28 |
| PLEKHN1 | 1.250264841 | 1.53E-32 |
| NCBP2L | 1.252681355 | 2.13E-11 |
| CGREF1 | 1.253079449 | 2.97E-12 |
| PRUNE2 | 1.253851665 | 3.40E-26 |
| CAPN11 | 1.254964412 | 1.43E-15 |
| ASCL3 | 1.255653686 | 1.92E-05 |
| AC019117.3 | 1.257487647 | 4.98E-08 |
| IGSF9B | 1.258002441 | 7.75E-25 |
| CD200R1L | 1.258304348 | 0.000240533 |
| PLPP4 | 1.259319211 | 1.20E-09 |
| CCDC151 | 1.260112689 | 3.92E-11 |
| ARHGAP39 | 1.260689225 | 1.24E-16 |
| LCN6 | 1.261671356 | 1.01E-11 |
| CFAP45 | 1.2617159 | 3.68E-09 |
| CFH | 1.261797407 | 1.39E-56 |
| TTC23L | 1.262081525 | 3.00E-11 |
| TMEM212 | 1.262322905 | 1.17E-11 |
| KCNE1B | 1.262356526 | 9.26E-09 |
| TPBG | 1.262932144 | 1.05E-53 |
| BMP7 | 1.26304978 | 7.44E-10 |
| SLC10A6 | 1.263428481 | 2.00E-20 |
| COL4A6 | 1.263602589 | 2.66E-24 |
| CEL | 1.26365568 | 5.74E-28 |
| RAMP1 | 1.264404318 | 8.29E-18 |
| COL5A1 | 1.264464966 | 1.13E-20 |
| C14orf180 | 1.264641758 | 6.10E-14 |
| MORN2 | 1.264653849 | 1.74E-14 |
| VCAM1 | 1.265124043 | 4.99E-10 |
| RIBC1 | 1.266081133 | 3.65E-10 |
| CPXM2 | 1.267331003 | 9.56E-19 |
| P4HA3 | 1.268053362 | 1.63E-20 |
| EPPIN | 1.26909603 | 3.59E-10 |
| SLC2A1 | 1.269611536 | 3.60E-22 |
| KCNG1 | 1.272582004 | 9.87E-13 |
| HOXC5 | 1.273006744 | 2.71E-06 |
| OMG | 1.274402902 | 5.11E-11 |
| ARTN | 1.275035792 | 4.92E-21 |
| KIAA1755 | 1.275805581 | 2.17E-17 |
| TGFBR3L | 1.276446402 | 1.92E-15 |
| MAGEA9B | 1.278424107 | 0.011639631 |
| SPA17 | 1.278500036 | 1.92E-16 |
| PCDH7 | 1.278802154 | 4.15E-23 |
| ARL9 | 1.279852858 | 6.50E-35 |
| C2orf81 | 1.280040622 | 9.39E-14 |
| HTR3D | 1.280589009 | 0.011515425 |
| ZNF473 | 1.280957552 | 8.72E-16 |
| RBM24 | 1.281444011 | 1.73E-12 |
| BDKRB1 | 1.282065357 | 6.19E-11 |
| PAMR1 | 1.283070381 | 1.24E-17 |
| SERINC2 | 1.283182489 | 2.83E-18 |
| DKK1 | 1.283457469 | 2.73E-08 |
| TIMD4 | 1.284693209 | 0.000401049 |
| FHOD3 | 1.28515698 | 1.53E-16 |
| CCL1 | 1.286779068 | 4.27E-05 |
| NGFR | 1.28680299 | 0.010747888 |
| ARX | 1.287809899 | 0.03167094 |
| ASTN1 | 1.288958893 | 5.20E-06 |
| LRRC17 | 1.289199844 | 1.44E-20 |
| DNAH5 | 1.290443332 | 8.40E-10 |
| CRABP2 | 1.291008593 | 1.83E-10 |
| DUPD1 | 1.291585614 | 0.00386712 |
| TUBB8 | 1.293226456 | 6.74E-07 |
| KCNH3 | 1.293397711 | 5.01E-07 |
| GPR26 | 1.294905384 | 6.35E-07 |
| EPPIN-WFDC6 | 1.297040902 | 0.000449987 |
| C2orf70 | 1.297647734 | 1.82E-10 |
| IRF4 | 1.301072947 | 1.77E-07 |
| FA2H | 1.301305008 | 4.29E-08 |
| AC020909.1 | 1.302494094 | 0.00019346 |
| DRC3 | 1.302546619 | 9.97E-13 |
| GPC1 | 1.303090518 | 9.69E-20 |
| MEIG1 | 1.303231472 | 1.49E-13 |
| AL929554.1 | 1.305298649 | 0.009580004 |
| KCNG2 | 1.306356691 | 7.56E-15 |
| ENTHD1 | 1.307150673 | 3.24E-13 |
| ANXA10 | 1.308169759 | 5.77E-11 |
| BHLHE22 | 1.308357264 | 6.80E-21 |
| DNM1 | 1.308957446 | 5.00E-19 |
| TPTE2 | 1.311622927 | 8.24E-22 |
| WFDC6 | 1.311980823 | 0.000156841 |
| SPACA3 | 1.312016761 | 1.04E-19 |
| SERTM2 | 1.312608948 | 1.05E-05 |
| MMP9 | 1.314175235 | 3.06E-07 |
| TP53TG3C | 1.31458418 | 0.006430676 |
| ANK2 | 1.315146615 | 3.44E-24 |
| FBXO15 | 1.318373133 | 3.43E-13 |
| NTN1 | 1.320180707 | 9.99E-05 |
| COLEC11 | 1.323502969 | 1.75E-21 |
| COL5A2 | 1.324195112 | 1.40E-07 |
| TEKT4 | 1.324364295 | 6.63E-11 |
| SERPINE2 | 1.324987572 | 3.59E-15 |
| PLPPR3 | 1.325110503 | 3.88E-11 |
| DEGS2 | 1.325202844 | 2.30E-08 |
| SAA2 | 1.326638028 | 4.81E-05 |
| KIF24 | 1.326839771 | 5.58E-17 |
| CCDC96 | 1.327138346 | 5.40E-14 |
| FBXO32 | 1.329629976 | 1.91E-34 |
| STARD5 | 1.331447172 | 2.16E-37 |
| RFLNA | 1.332546095 | 6.52E-12 |
| ESR2 | 1.333358903 | 8.57E-35 |
| TCP11 | 1.334035295 | 2.03E-11 |
| SMKR1 | 1.335216745 | 7.57E-05 |
| CFAP58 | 1.337668241 | 4.66E-05 |
| GLRA2 | 1.337785638 | 1.18E-05 |
| SLC22A11 | 1.33835885 | 1.46E-15 |
| BOC | 1.338641753 | 2.19E-31 |
| MYCBPAP | 1.339174849 | 7.25E-14 |
| CHRNA9 | 1.34004998 | 0.031345207 |
| FCAMR | 1.341945897 | 4.73E-05 |
| PHYHIP | 1.342099386 | 1.40E-18 |
| C10orf90 | 1.342138889 | 4.07E-05 |
| IQCD | 1.342993296 | 6.59E-12 |
| CPNE7 | 1.343284603 | 1.37E-17 |
| CFAP70 | 1.343313114 | 4.09E-11 |
| CAPNS2 | 1.345299737 | 1.27E-09 |
| HR | 1.346287064 | 4.66E-32 |
| AC007731.4 | 1.347083448 | 0.00049943 |
| NHLRC4 | 1.347837455 | 6.60E-13 |
| FOXI3 | 1.347914548 | 1.44E-05 |
| ELOA3B | 1.348114714 | 0.026074158 |
| NEK5 | 1.350081654 | 1.01E-10 |
| CTSE | 1.351159341 | 1.77E-16 |
| FCRL2 | 1.352043084 | 5.69E-22 |
| TOGARAM2 | 1.353360781 | 7.22E-12 |
| GOLM1 | 1.353789165 | 4.09E-57 |
| ITGA7 | 1.354633998 | 3.77E-25 |
| ACY3 | 1.355270132 | 0.006409136 |
| DAPL1 | 1.355529106 | 1.81E-09 |
| PAEP | 1.355862049 | 2.17E-08 |
| LMX1B | 1.356318341 | 4.12E-15 |
| ADM2 | 1.356916157 | 5.66E-22 |
| RET | 1.357123758 | 7.97E-26 |
| SOX30 | 1.358068472 | 3.09E-11 |
| RXFP3 | 1.359240045 | 9.25E-05 |
| MRGPRF | 1.359628988 | 4.01E-08 |
| SPATA18 | 1.362797208 | 1.60E-12 |
| TNFRSF17 | 1.362886494 | 1.97E-05 |
| SHOX2 | 1.363010038 | 1.64E-15 |
| FBXW10 | 1.363046682 | 8.19E-12 |
| TSHZ2 | 1.363338019 | 1.31E-23 |
| DNAJC12 | 1.364032195 | 3.45E-19 |
| TNNI3 | 1.364252863 | 1.35E-09 |
| TAGLN3 | 1.36442518 | 3.91E-07 |
| ASPHD1 | 1.366929376 | 2.27E-19 |
| KERA | 1.368634532 | 1.07E-13 |
| WDR49 | 1.369730389 | 6.37E-12 |
| MC5R | 1.370396379 | 1.85E-11 |
| CLEC2L | 1.370587648 | 2.21E-05 |
| IQCG | 1.373636898 | 2.24E-18 |
| RGS22 | 1.373800027 | 6.57E-16 |
| UGT1A1 | 1.376643386 | 4.76E-27 |
| E2F8 | 1.379476865 | 3.44E-12 |
| NXF2B | 1.380639252 | 9.36E-08 |
| KRT78 | 1.381844688 | 2.24E-05 |
| CCR8 | 1.382327556 | 3.36E-11 |
| WDR66 | 1.38282787 | 1.71E-17 |
| CCDC173 | 1.383668027 | 2.27E-05 |
| CITED1 | 1.38377134 | 2.29E-11 |
| CMA1 | 1.38426376 | 1.23E-07 |
| CCDC146 | 1.384304871 | 4.37E-17 |
| CHST8 | 1.386144922 | 1.96E-22 |
| FJX1 | 1.38655312 | 8.57E-24 |
| DNAH10 | 1.387474685 | 2.76E-12 |
| FMO1 | 1.387747461 | 3.86E-12 |
| LEMD1 | 1.387819724 | 1.02E-13 |
| ABCA4 | 1.388274832 | 5.90E-26 |
| FRMD5 | 1.388540752 | 1.04E-15 |
| TSNAXIP1 | 1.388759301 | 6.07E-13 |
| TNN | 1.388781274 | 2.64E-18 |
| SIX4 | 1.389230636 | 3.80E-06 |
| C4B | 1.389338064 | 6.88E-29 |
| AGR2 | 1.390023355 | 1.85E-15 |
| FBXO39 | 1.390591581 | 1.97E-20 |
| RAET1L | 1.390709187 | 3.57E-08 |
| ASB2 | 1.390864165 | 1.51E-25 |
| MCHR1 | 1.391468371 | 1.42E-12 |
| CD24 | 1.391861486 | 3.93E-17 |
| MAPK15 | 1.393697188 | 2.23E-11 |
| TCP11X1 | 1.393852229 | 0.001278945 |
| ZSCAN4 | 1.394120313 | 1.16E-23 |
| COX6B2 | 1.394566387 | 1.48E-17 |
| ACSBG1 | 1.395668764 | 1.88E-08 |
| TAF7L | 1.39595347 | 4.69E-24 |
| NLGN4Y | 1.396791946 | 0.000496773 |
| SSUH2 | 1.400239946 | 1.43E-14 |
| EFHC2 | 1.400382182 | 1.82E-10 |
| TEX44 | 1.400544025 | 2.83E-06 |
| NKAIN4 | 1.400712387 | 3.46E-16 |
| CCR7 | 1.400748296 | 3.53E-26 |
| UTS2R | 1.401120588 | 8.94E-10 |
| PAPPA | 1.401620547 | 1.90E-06 |
| DPYS | 1.402161695 | 5.83E-15 |
| SCRG1 | 1.402568505 | 9.09E-21 |
| MAGED4 | 1.402884371 | 1.95E-45 |
| PACRG | 1.403843562 | 1.61E-12 |
| F2RL2 | 1.403962647 | 0.020519949 |
| CGB2 | 1.404439928 | 0.007981276 |
| SAGE1 | 1.405777246 | 0.001582643 |
| VWA3A | 1.40606425 | 2.95E-10 |
| ENAM | 1.407252389 | 1.39E-21 |
| AIPL1 | 1.408426066 | 2.62E-21 |
| MOBP | 1.408437105 | 1.95E-10 |
| KCNN4 | 1.408597089 | 6.05E-31 |
| LRGUK | 1.409527987 | 8.12E-15 |
| ANKRD45 | 1.410652918 | 4.52E-13 |
| INSYN2 | 1.411010577 | 0.006242031 |
| AIRE | 1.413452644 | 1.49E-16 |
| UGT2B17 | 1.413964924 | 2.26E-07 |
| PPP2R2C | 1.415011717 | 6.02E-23 |
| IRX4 | 1.415202934 | 0.03152746 |
| DCLK1 | 1.41559108 | 1.44E-41 |
| KLHL4 | 1.416901432 | 1.10E-11 |
| C4A | 1.416946478 | 2.12E-30 |
| RARRES1 | 1.420993041 | 3.26E-20 |
| CNTN3 | 1.421508613 | 6.47E-26 |
| SCGB1A1 | 1.421756476 | 1.46E-06 |
| FAM187A | 1.422383419 | 2.19E-16 |
| DTHD1 | 1.424178362 | 8.44E-11 |
| TMEM231 | 1.424850221 | 2.21E-16 |
| CFAP299 | 1.426587607 | 2.63E-07 |
| CFAP61 | 1.42779545 | 2.70E-15 |
| TRIP13 | 1.429000434 | 3.06E-08 |
| CCL25 | 1.429844226 | 3.85E-08 |
| ENTPD8 | 1.43050353 | 1.11E-15 |
| SPINK1 | 1.430720447 | 1.79E-07 |
| CLEC4G | 1.433310293 | 3.24E-13 |
| C12orf42 | 1.433324178 | 6.81E-12 |
| OPN5 | 1.433544055 | 3.46E-05 |
| FBLN2 | 1.434171062 | 8.01E-25 |
| SSC5D | 1.436239858 | 1.17E-28 |
| COL6A3 | 1.436275598 | 9.29E-32 |
| CYP2J2 | 1.436727631 | 1.02E-07 |
| AXDND1 | 1.437075814 | 2.07E-11 |
| HRK | 1.437106059 | 2.93E-14 |
| HYDIN | 1.43720702 | 1.41E-13 |
| P2RX5 | 1.43737805 | 2.22E-18 |
| HS6ST3 | 1.438303114 | 6.35E-12 |
| GRM4 | 1.439216258 | 2.47E-17 |
| ERICH5 | 1.440400076 | 0.000871359 |
| FHAD1 | 1.440498083 | 1.81E-13 |
| BLK | 1.442427303 | 6.10E-22 |
| PNLDC1 | 1.443857815 | 0.000459107 |
| MAGED4B | 1.445026068 | 5.39E-37 |
| CLEC17A | 1.446265869 | 9.42E-16 |
| ZNF474 | 1.446520383 | 4.36E-07 |
| CD38 | 1.448559946 | 1.53E-20 |
| SERPINI2 | 1.4491769 | 7.92E-13 |
| OR1J4 | 1.449278108 | 0.001267695 |
| AK8 | 1.451972325 | 1.55E-12 |
| DYNLRB2 | 1.452965139 | 2.82E-09 |
| ASPN | 1.453858199 | 1.87E-21 |
| RGS4 | 1.455244055 | 7.45E-14 |
| TM4SF19 | 1.455622938 | 6.49E-14 |
| HCAR1 | 1.459379823 | 2.40E-27 |
| ISLR | 1.460624076 | 2.86E-24 |
| ETDB | 1.461674691 | 0.002112679 |
| ADAM23 | 1.46178326 | 2.28E-26 |
| TEKT2 | 1.461795929 | 4.64E-11 |
| PIH1D2 | 1.462052137 | 1.23E-15 |
| SRRM3 | 1.462925181 | 2.49E-33 |
| ANKK1 | 1.464759462 | 3.26E-15 |
| CFAP206 | 1.465152363 | 3.31E-09 |
| ADGB | 1.466320381 | 5.34E-11 |
| FGF11 | 1.466801082 | 2.29E-31 |
| SBSN | 1.466871555 | 0.000146594 |
| CENPM | 1.467885351 | 2.49E-20 |
| RAB36 | 1.469016825 | 7.31E-19 |
| DCDC1 | 1.469134848 | 5.67E-10 |
| PLA2G2D | 1.469532937 | 4.75E-09 |
| CCL13 | 1.469756133 | 5.00E-06 |
| THRSP | 1.47121348 | 1.33E-08 |
| WNT4 | 1.471711168 | 4.80E-18 |
| LRRN1 | 1.471782446 | 1.02E-36 |
| DNAH6 | 1.472463437 | 6.94E-13 |
| C4orf47 | 1.474713207 | 4.40E-13 |
| 43534 | 1.475748969 | 2.01E-09 |
| MYO3B | 1.475796694 | 1.10E-50 |
| CFB | 1.47715909 | 1.10E-30 |
| RIIAD1 | 1.477268331 | 1.32E-10 |
| OR2D2 | 1.477578962 | 0.002686879 |
| PTPRZ1 | 1.477732459 | 1.97E-18 |
| ARSH | 1.478778304 | 2.71E-06 |
| C11orf88 | 1.47937471 | 1.28E-07 |
| OR51E1 | 1.479711185 | 0.000926801 |
| TCTE1 | 1.479915309 | 3.06E-05 |
| CCDC80 | 1.47992224 | 2.63E-17 |
| OXTR | 1.480452991 | 1.36E-13 |
| C21orf58 | 1.481699598 | 3.56E-15 |
| C16orf71 | 1.481730793 | 4.80E-14 |
| LMNTD1 | 1.482304655 | 4.84E-09 |
| TRPV6 | 1.482348221 | 3.03E-20 |
| AQP6 | 1.482705123 | 2.61E-16 |
| AOC1 | 1.484278968 | 1.43E-33 |
| SPAG8 | 1.484971059 | 3.92E-14 |
| SLC27A2 | 1.485197951 | 4.55E-08 |
| IL36B | 1.485920365 | 0.00139608 |
| CIDEA | 1.487165616 | 3.73E-05 |
| LYPD1 | 1.487650208 | 8.73E-19 |
| SAXO2 | 1.488231773 | 8.60E-12 |
| ADAM28 | 1.489063222 | 4.59E-31 |
| CCDC74A | 1.48931679 | 8.28E-16 |
| TCTEX1D4 | 1.490140975 | 7.94E-13 |
| TNFSF11 | 1.490414526 | 1.85E-13 |
| ARMC4 | 1.492899121 | 1.69E-11 |
| ITGB4 | 1.493250169 | 3.58E-41 |
| CCDC74B | 1.496766303 | 1.10E-11 |
| PIWIL1 | 1.497329624 | 4.72E-05 |
| TMEM200C | 1.498740337 | 2.80E-24 |
| CDHR2 | 1.499630816 | 2.16E-22 |
| GNG4 | 1.500175705 | 2.39E-17 |
| CBARP | 1.500434165 | 3.90E-12 |
| OR10A4 | 1.50062574 | 0.007153518 |
| EFCAB10 | 1.500715202 | 1.10E-14 |
| B3GALT5 | 1.500734889 | 2.29E-11 |
| COL1A2 | 1.501984641 | 1.49E-26 |
| PTPRH | 1.504977764 | 9.32E-16 |
| LRRC73 | 1.504984744 | 2.11E-14 |
| CCDC103 | 1.505109883 | 1.38E-13 |
| KNCN | 1.506612684 | 3.42E-05 |
| DEUP1 | 1.506808014 | 4.25E-11 |
| KLHL13 | 1.507236904 | 2.03E-31 |
| CROCC2 | 1.50918696 | 2.82E-12 |
| PRR15 | 1.511934126 | 3.57E-12 |
| CRISP2 | 1.512977882 | 4.06E-07 |
| SCAMP5 | 1.513027146 | 1.29E-32 |
| C2orf50 | 1.513220722 | 6.80E-11 |
| NKX6-3 | 1.513270309 | 7.96E-10 |
| C7orf57 | 1.514527857 | 1.96E-10 |
| SMOC1 | 1.514528679 | 2.56E-32 |
| CCND2 | 1.515518801 | 2.43E-17 |
| AKR1B10 | 1.518092271 | 2.18E-05 |
| ADAMDEC1 | 1.518664105 | 2.15E-09 |
| DYDC1 | 1.519988714 | 9.33E-06 |
| FOXN4 | 1.52214155 | 4.18E-08 |
| CD79B | 1.524435031 | 9.33E-32 |
| CASC1 | 1.525066837 | 8.06E-13 |
| KCNJ16 | 1.525779232 | 1.15E-14 |
| RNF183 | 1.525827824 | 4.89E-16 |
| TMPRSS11A | 1.52611893 | 5.77E-05 |
| CALML5 | 1.527556336 | 0.004130497 |
| BTG4 | 1.529203703 | 1.04E-06 |
| FAM19A3 | 1.529237201 | 1.81E-09 |
| GRIN3B | 1.530762688 | 3.01E-15 |
| TTLL10 | 1.531561025 | 2.49E-12 |
| POU3F2 | 1.531602949 | 4.10E-07 |
| AQP2 | 1.531711316 | 1.17E-16 |
| SERPINB3 | 1.53215497 | 0.031649411 |
| LRRC43 | 1.533147711 | 8.71E-13 |
| CHRDL2 | 1.534785161 | 5.61E-37 |
| WNT10A | 1.536595119 | 2.80E-17 |
| MSGN1 | 1.53728754 | 0.015272931 |
| KIF19 | 1.538159976 | 5.56E-17 |
| THEG | 1.53963745 | 5.07E-09 |
| UGT2A2 | 1.540014949 | 0.033810313 |
| DOC2A | 1.540348468 | 4.70E-14 |
| DUSP14 | 1.543083141 | 9.47E-18 |
| ELOA3 | 1.543110624 | 0.001420173 |
| CPXCR1 | 1.546139921 | 1.53E-07 |
| DEFB4A | 1.549052005 | 0.000274389 |
| CHST4 | 1.549465942 | 5.76E-18 |
| FOXI2 | 1.549901172 | 1.31E-19 |
| HOXC4 | 1.550316031 | 5.07E-14 |
| CLDN16 | 1.55195344 | 1.05E-11 |
| GYG2 | 1.552183519 | 2.93E-36 |
| SLC23A1 | 1.552762942 | 8.96E-10 |
| SPOCK3 | 1.553507363 | 3.21E-12 |
| CAPS | 1.555140092 | 2.64E-09 |
| RAB26 | 1.557062472 | 6.71E-38 |
| IL37 | 1.557240284 | 3.05E-11 |
| GLIS1 | 1.55861788 | 9.13E-27 |
| CFAP74 | 1.56016114 | 1.03E-14 |
| AADACL2 | 1.560623593 | 1.30E-06 |
| ANKFN1 | 1.562041641 | 1.77E-13 |
| CACNA1G | 1.562180976 | 6.35E-30 |
| CNTN5 | 1.562447529 | 4.49E-11 |
| C1orf141 | 1.563156128 | 4.15E-08 |
| C2orf73 | 1.56589226 | 9.30E-12 |
| CDHR3 | 1.565949943 | 5.42E-13 |
| FAM227A | 1.566519633 | 3.47E-16 |
| SPTBN2 | 1.567146699 | 9.30E-47 |
| RASAL1 | 1.567888952 | 1.69E-20 |
| CATSPERD | 1.568555838 | 2.97E-07 |
| SNX31 | 1.571093564 | 8.89E-10 |
| SAMD15 | 1.573695956 | 1.50E-18 |
| ITPRID1 | 1.573873088 | 8.29E-24 |
| AC235565.2 | 1.574721669 | 2.84E-05 |
| IZUMO1R | 1.575275004 | 5.57E-06 |
| CCL24 | 1.577623149 | 2.00E-12 |
| CLIC6 | 1.578492537 | 1.17E-30 |
| C3orf67 | 1.581441462 | 8.63E-24 |
| C1orf87 | 1.582209499 | 2.39E-09 |
| CFAP57 | 1.582392437 | 2.03E-12 |
| CTXND2 | 1.583823785 | 2.37E-09 |
| RAET1E | 1.585073799 | 1.71E-24 |
| ABCB11 | 1.585643781 | 0.005169255 |
| FAM71E2 | 1.586372274 | 0.006372085 |
| DGKI | 1.587385093 | 4.64E-26 |
| OSR2 | 1.588473275 | 9.21E-22 |
| HEPACAM | 1.58913606 | 0.040085273 |
| TCP11X2 | 1.589392964 | 5.25E-05 |
| NEK10 | 1.589706157 | 1.64E-12 |
| NME9 | 1.590860781 | 5.15E-14 |
| PRB3 | 1.591031073 | 1.34E-10 |
| ERVV-1 | 1.591037816 | 1.53E-08 |
| ZNF215 | 1.592603462 | 1.18E-38 |
| VWA3B | 1.594874321 | 1.55E-09 |
| C20orf141 | 1.595105138 | 0.007573961 |
| SERPINA5 | 1.595585506 | 8.06E-17 |
| ST6GALNAC1 | 1.5957843 | 1.25E-25 |
| FERMT1 | 1.596883628 | 1.10E-40 |
| MROH9 | 1.597101432 | 6.20E-10 |
| BCAS1 | 1.597598057 | 1.65E-16 |
| DBX2 | 1.601619082 | 9.60E-05 |
| COL11A1 | 1.602523635 | 2.79E-11 |
| CFAP97D2 | 1.603399657 | 1.83E-07 |
| DNAH7 | 1.603969946 | 1.10E-14 |
| CCDC113 | 1.605994543 | 1.98E-14 |
| PRR4 | 1.606125647 | 1.57E-11 |
| ADGRB2 | 1.609107327 | 1.49E-42 |
| AC013470.2 | 1.609522339 | 7.07E-09 |
| NRAP | 1.611326674 | 1.55E-09 |
| SOWAHA | 1.611660942 | 1.75E-14 |
| LCN2 | 1.612240985 | 9.04E-14 |
| FAM166B | 1.612765793 | 3.36E-10 |
| CFAP52 | 1.615126133 | 4.57E-10 |
| MESP2 | 1.618588759 | 3.07E-15 |
| TP73 | 1.620718598 | 3.52E-12 |
| LRRIQ1 | 1.622003943 | 4.80E-11 |
| FAP | 1.622541393 | 2.96E-25 |
| AVPR1A | 1.622951756 | 5.75E-19 |
| ADGRA1 | 1.623611855 | 7.53E-12 |
| WDR63 | 1.623807703 | 4.22E-11 |
| RSPH9 | 1.624320536 | 1.69E-14 |
| SSTR2 | 1.625642641 | 0.024538529 |
| COCH | 1.626405431 | 2.87E-10 |
| MGAT4D | 1.627078582 | 5.71E-07 |
| PIFO | 1.627923137 | 1.01E-13 |
| TRPV4 | 1.628600582 | 2.26E-27 |
| CDK5R2 | 1.629462101 | 1.32E-05 |
| ATP6V0A4 | 1.629645268 | 6.71E-11 |
| GDF15 | 1.629898211 | 1.63E-23 |
| XPNPEP2 | 1.632477354 | 6.41E-09 |
| AGBL2 | 1.63311247 | 2.69E-15 |
| FEZF1 | 1.633142861 | 1.75E-09 |
| GSDMC | 1.633167348 | 7.75E-25 |
| WNK2 | 1.634175175 | 2.94E-21 |
| KCNH8 | 1.634436559 | 1.62E-13 |
| DNAJB13 | 1.635127026 | 1.15E-13 |
| HABP2 | 1.636008185 | 7.48E-07 |
| GBP6 | 1.638277723 | 4.58E-15 |
| PSCA | 1.638325238 | 4.23E-13 |
| GPR12 | 1.638794043 | 0.000112643 |
| NPSR1 | 1.639212532 | 5.92E-06 |
| LCN10 | 1.641154916 | 1.29E-19 |
| DES | 1.642197327 | 1.30E-22 |
| KCNRG | 1.642928271 | 8.30E-11 |
| SPATA17 | 1.643260978 | 1.02E-15 |
| ZG16B | 1.645229905 | 3.88E-15 |
| TTLL9 | 1.64564983 | 1.20E-12 |
| RSPH10B2 | 1.647350802 | 3.64E-11 |
| THBS4 | 1.647579775 | 5.87E-31 |
| SPDEF | 1.647739739 | 3.82E-09 |
| RSPH4A | 1.651534091 | 3.18E-13 |
| DLL3 | 1.651833405 | 5.68E-11 |
| LRRC71 | 1.652157522 | 2.48E-13 |
| ZMYND10 | 1.65233844 | 1.26E-11 |
| SYCE3 | 1.652669415 | 0.02984845 |
| TRIM29 | 1.653078865 | 1.70E-18 |
| HMGA2 | 1.653665555 | 2.89E-24 |
| MAP1A | 1.654459373 | 1.69E-07 |
| FLNC | 1.658426069 | 2.05E-22 |
| TMEM179 | 1.659988864 | 1.10E-12 |
| FAM95C | 1.660662779 | 2.33E-21 |
| NGEF | 1.661934207 | 1.70E-17 |
| SALL4 | 1.664247325 | 1.98E-11 |
| TRDN | 1.666143439 | 4.43E-07 |
| CFAP65 | 1.666471828 | 2.16E-10 |
| TTC29 | 1.666535159 | 9.51E-10 |
| ANKRD66 | 1.666594378 | 6.61E-09 |
| CWH43 | 1.667017116 | 4.96E-09 |
| SPAG6 | 1.667814444 | 3.85E-10 |
| MDH1B | 1.668597264 | 1.46E-14 |
| SAMD11 | 1.668908245 | 3.66E-34 |
| ROPN1L | 1.669378469 | 9.06E-13 |
| CRHR1 | 1.669716479 | 8.51E-09 |
| ERICH3 | 1.670579717 | 2.41E-09 |
| CEACAM7 | 1.670701098 | 1.51E-16 |
| TCERG1L | 1.671533789 | 1.84E-15 |
| MYL2 | 1.672069423 | 9.84E-10 |
| FAM216B | 1.674056594 | 2.61E-07 |
| SLC6A2 | 1.675319415 | 2.52E-13 |
| PSG11 | 1.6770628 | 0.000125851 |
| TRIM55 | 1.677432077 | 2.61E-20 |
| NKAIN3 | 1.677737342 | 8.76E-07 |
| ALDH3A1 | 1.679971694 | 2.24E-14 |
| C1orf158 | 1.680552532 | 5.56E-09 |
| PPP1R42 | 1.681146253 | 6.52E-11 |
| MEDAG | 1.682249693 | 0.01262357 |
| SYNDIG1 | 1.682256025 | 0.016754884 |
| FNDC4 | 1.682529315 | 6.09E-18 |
| SPEF1 | 1.683220161 | 3.32E-11 |
| DNAH2 | 1.684787368 | 2.01E-14 |
| C11orf97 | 1.685084288 | 1.76E-08 |
| NWD1 | 1.686693945 | 2.45E-10 |
| MAP3K19 | 1.687090644 | 3.21E-10 |
| FAM81B | 1.687091831 | 3.78E-12 |
| STOX1 | 1.687201317 | 1.68E-11 |
| TMEM156 | 1.687601041 | 3.32E-19 |
| AC117457.1 | 1.689136757 | 0.009068619 |
| CCDC114 | 1.691393317 | 6.04E-12 |
| PIM2 | 1.691430204 | 2.69E-42 |
| DLEC1 | 1.691475591 | 8.87E-15 |
| CCDC78 | 1.692459023 | 2.10E-12 |
| C6 | 1.692911811 | 1.01E-15 |
| GAS2L2 | 1.693534956 | 2.05E-12 |
| CFAP73 | 1.694103585 | 2.73E-11 |
| NKX6-1 | 1.694744581 | 1.17E-14 |
| FAM183A | 1.695543962 | 3.23E-10 |
| ARL14 | 1.696861422 | 4.25E-14 |
| PLXNA4 | 1.697321928 | 4.30E-45 |
| C11orf16 | 1.699688936 | 3.26E-10 |
| GATA4 | 1.700886337 | 7.04E-09 |
| LRRC74B | 1.702029561 | 6.87E-11 |
| VAT1L | 1.703112256 | 0.000367798 |
| BCL2L10 | 1.704415785 | 1.51E-16 |
| HTR3A | 1.705298694 | 4.55E-14 |
| HNF4A | 1.705647681 | 9.26E-25 |
| MS4A8 | 1.7072691 | 2.68E-08 |
| STOML3 | 1.707359444 | 7.02E-09 |
| C5orf49 | 1.707648796 | 2.29E-11 |
| IL1RAPL2 | 1.708910114 | 0.000221103 |
| HAS1 | 1.7116639 | 8.72E-07 |
| ALOX15 | 1.711768509 | 8.10E-10 |
| EFCAB1 | 1.711770719 | 1.36E-11 |
| CFAP47 | 1.712112985 | 5.14E-10 |
| DCDC2B | 1.712864455 | 8.94E-13 |
| HTR2A | 1.713651333 | 4.33E-32 |
| ALKAL1 | 1.71426378 | 4.73E-06 |
| RIBC2 | 1.715868497 | 1.09E-15 |
| ZNF469 | 1.71754101 | 6.05E-40 |
| CFAP300 | 1.718862278 | 4.72E-19 |
| TMCO5A | 1.719352766 | 4.73E-08 |
| SPINK4 | 1.720527018 | 0.045682324 |
| LHX8 | 1.721196802 | 1.60E-05 |
| CNGA3 | 1.722209555 | 1.06E-11 |
| CFAP46 | 1.724516261 | 6.65E-12 |
| SLC44A4 | 1.724897993 | 2.17E-22 |
| CCDC185 | 1.725456362 | 3.06E-09 |
| STAP1 | 1.725888074 | 7.77E-24 |
| SPIB | 1.728158191 | 1.44E-20 |
| STMND1 | 1.730931042 | 3.02E-12 |
| SPAG4 | 1.732341329 | 2.40E-40 |
| ASIC1 | 1.733039846 | 8.45E-37 |
| ARMC3 | 1.73418017 | 2.96E-11 |
| FANK1 | 1.735122888 | 9.14E-20 |
| ABCA13 | 1.735282069 | 8.29E-11 |
| SLC38A11 | 1.736089413 | 7.59E-31 |
| CCDC160 | 1.739701323 | 3.55E-18 |
| TMEM45A | 1.739815052 | 1.27E-49 |
| PRR18 | 1.741646873 | 0.000286631 |
| KRT32 | 1.742494712 | 1.42E-12 |
| NDST4 | 1.743239503 | 3.49E-08 |
| RSPH14 | 1.743861536 | 1.54E-12 |
| DNAH9 | 1.74442841 | 4.89E-12 |
| VWCE | 1.745686661 | 7.08E-29 |
| IGDCC4 | 1.746798299 | 4.21E-24 |
| SPATA4 | 1.752373738 | 4.15E-12 |
| C9orf24 | 1.755163031 | 8.36E-11 |
| RAD51AP2 | 1.755598761 | 3.16E-19 |
| CHRNA4 | 1.755954306 | 2.09E-06 |
| DYDC2 | 1.756376609 | 5.02E-10 |
| PI15 | 1.756995852 | 9.99E-05 |
| DMP1 | 1.757777954 | 5.98E-08 |
| RSPH10B | 1.757939676 | 1.74E-13 |
| TEKT1 | 1.759876515 | 4.44E-12 |
| CFAP43 | 1.760414628 | 5.60E-12 |
| HS3ST6 | 1.76273515 | 9.16E-10 |
| LDLRAD1 | 1.76334611 | 3.30E-11 |
| PTHLH | 1.764062822 | 8.89E-29 |
| MORN5 | 1.764142501 | 1.44E-10 |
| MAT1A | 1.765989287 | 2.53E-10 |
| GUCY2F | 1.76630621 | 1.06E-08 |
| CPNE6 | 1.768640814 | 2.20E-15 |
| EEF1AKMT4-ECE2 | 1.769654283 | 0.000291916 |
| DIPK1C | 1.769924388 | 4.13E-16 |
| DNAAF3 | 1.779907691 | 2.97E-13 |
| NPY2R | 1.779915277 | 4.83E-05 |
| TMEM95 | 1.780416728 | 1.51E-11 |
| CFAP77 | 1.780812687 | 7.02E-11 |
| OR6A2 | 1.781194528 | 1.46E-08 |
| C9orf135 | 1.783767462 | 3.94E-09 |
| KRTAP13-2 | 1.784275762 | 0.004983199 |
| BFSP2 | 1.784405719 | 3.31E-21 |
| ANKUB1 | 1.786323992 | 1.59E-14 |
| ENKUR | 1.789522479 | 7.75E-12 |
| C1orf194 | 1.790704378 | 5.28E-10 |
| AC009163.2 | 1.794337255 | 1.37E-06 |
| KRT37 | 1.795640344 | 0.000873824 |
| AKAP14 | 1.79616333 | 4.50E-10 |
| SNAP25 | 1.799470993 | 2.65E-38 |
| PTPRT | 1.799651786 | 5.72E-13 |
| DQX1 | 1.799897498 | 8.95E-27 |
| GDA | 1.800663951 | 6.45E-27 |
| TMEM158 | 1.802053968 | 1.39E-34 |
| ZPLD1 | 1.805206261 | 5.11E-14 |
| LGSN | 1.805328145 | 3.76E-17 |
| THBS2 | 1.806526578 | 1.19E-27 |
| CCDC33 | 1.8084282 | 9.55E-11 |
| SNTN | 1.80961994 | 4.51E-11 |
| C12orf74 | 1.810894792 | 8.66E-15 |
| LRRC46 | 1.811057505 | 1.60E-12 |
| WDR38 | 1.813515006 | 2.48E-10 |
| CFAP157 | 1.81564147 | 2.80E-13 |
| CFAP126 | 1.817711852 | 5.70E-11 |
| SPAG17 | 1.817810675 | 6.99E-13 |
| CCDC17 | 1.818261747 | 6.06E-15 |
| CHST9 | 1.81921596 | 1.59E-09 |
| ECT2L | 1.820089877 | 2.81E-12 |
| EPN3 | 1.82080144 | 3.17E-24 |
| JSRP1 | 1.82103191 | 4.74E-34 |
| PTGFRN | 1.822148713 | 5.48E-44 |
| OR2D3 | 1.822369281 | 0.000560312 |
| MMP12 | 1.822790526 | 4.06E-09 |
| SLC28A3 | 1.825183217 | 4.44E-27 |
| SERPINA4 | 1.827130786 | 0.000564032 |
| CCDC60 | 1.8271328 | 1.35E-16 |
| GABRG3 | 1.827259898 | 1.27E-09 |
| CACNG7 | 1.827489294 | 0.01600048 |
| TUBA4B | 1.829020114 | 8.22E-12 |
| AC008397.1 | 1.829514899 | 1.02E-09 |
| CNGA4 | 1.830428299 | 6.47E-14 |
| SERPINA6 | 1.831893239 | 2.17E-05 |
| CCDC187 | 1.834945432 | 2.79E-11 |
| C6orf118 | 1.836735596 | 7.29E-10 |
| KRT34 | 1.83794238 | 6.31E-05 |
| POU2AF1 | 1.838494681 | 5.00E-16 |
| CNTNAP4 | 1.840817937 | 2.05E-07 |
| CACNG6 | 1.841327838 | 7.50E-12 |
| SMPX | 1.841355931 | 0.023189216 |
| ACKR1 | 1.843440514 | 1.19E-19 |
| TMEM132E | 1.846231466 | 3.03E-30 |
| ENTPD2 | 1.849134975 | 1.16E-21 |
| KRT74 | 1.851144479 | 1.53E-06 |
| DRC7 | 1.851209905 | 1.05E-13 |
| DNAH3 | 1.855011855 | 4.70E-15 |
| KCNS2 | 1.855988376 | 2.79E-24 |
| CCDC190 | 1.857693743 | 5.63E-11 |
| CRLF1 | 1.858091312 | 9.65E-28 |
| C22orf15 | 1.858459244 | 1.99E-14 |
| CFAP99 | 1.862022167 | 3.44E-15 |
| PRH2 | 1.862449967 | 4.89E-11 |
| KRT40 | 1.863681735 | 9.39E-13 |
| SYT16 | 1.864600579 | 5.15E-35 |
| DRC1 | 1.864982963 | 1.47E-08 |
| ACAN | 1.865336787 | 1.08E-19 |
| PAPPA2 | 1.865557819 | 2.43E-06 |
| HNF4G | 1.866727602 | 6.44E-18 |
| LCE1B | 1.869244809 | 0.010633264 |
| C20orf85 | 1.870569331 | 6.47E-11 |
| ACTG2 | 1.871279679 | 1.09E-25 |
| STEAP1 | 1.873378113 | 2.34E-30 |
| DNAAF1 | 1.87468064 | 2.76E-16 |
| AC011604.2 | 1.875945857 | 0.007226158 |
| PCSK1N | 1.876109234 | 0.004399376 |
| FAM92B | 1.877160662 | 3.32E-12 |
| MRLN | 1.878076629 | 4.46E-19 |
| RSPH1 | 1.879104233 | 4.94E-12 |
| ARHGAP40 | 1.879118031 | 8.18E-27 |
| HHATL | 1.884868441 | 8.53E-16 |
| TFAP2B | 1.88698056 | 0.000196213 |
| MIXL1 | 1.887188061 | 4.05E-30 |
| PIH1D3 | 1.889745226 | 3.69E-11 |
| HPSE2 | 1.89016048 | 9.70E-34 |
| TRIM49C | 1.891637019 | 0.035312347 |
| CBLN4 | 1.896864518 | 1.88E-14 |
| APOBEC4 | 1.896988041 | 2.55E-08 |
| AC007906.2 | 1.897367098 | 3.66E-16 |
| ZBBX | 1.89912903 | 1.62E-11 |
| PNCK | 1.904289978 | 7.65E-29 |
| BHLHA15 | 1.904736711 | 2.31E-30 |
| CAPSL | 1.905147794 | 9.04E-11 |
| ALDH3B2 | 1.905327722 | 1.28E-24 |
| EBF3 | 1.906300966 | 5.31E-31 |
| AC092143.1 | 1.907528018 | 0.009460181 |
| DNAH12 | 1.909749942 | 4.37E-14 |
| EYA1 | 1.911680697 | 6.81E-10 |
| MMP7 | 1.913069313 | 5.87E-31 |
| TSPAN19 | 1.915167065 | 1.93E-13 |
| TNFRSF13B | 1.919217646 | 1.28E-24 |
| SLC52A1 | 1.919354984 | 1.12E-17 |
| GJB3 | 1.920147809 | 1.47E-27 |
| CYP2A13 | 1.924286137 | 2.03E-09 |
| IGF1 | 1.924477397 | 4.50E-36 |
| WDR87 | 1.925355048 | 4.07E-08 |
| RORB | 1.925483643 | 1.30E-08 |
| CDHR4 | 1.928074082 | 8.33E-13 |
| MTNR1A | 1.929659375 | 2.52E-09 |
| SPATS1 | 1.929681968 | 2.71E-08 |
| FNDC1 | 1.929757539 | 1.81E-17 |
| DNAI2 | 1.932213091 | 9.03E-14 |
| ESPN | 1.932466062 | 1.89E-19 |
| CD164L2 | 1.935782104 | 4.27E-18 |
| TTLL8 | 1.939192637 | 0.000237652 |
| CLCA2 | 1.940864804 | 2.87E-05 |
| SULF1 | 1.94117333 | 5.99E-55 |
| KRT23 | 1.944473728 | 4.22E-18 |
| EYA2 | 1.947432127 | 3.99E-31 |
| LTF | 1.947692841 | 1.73E-15 |
| CD27 | 1.947694314 | 3.78E-41 |
| XIRP1 | 1.948156804 | 5.35E-09 |
| KLK12 | 1.951739322 | 5.30E-10 |
| CKMT1A | 1.957618719 | 2.87E-27 |
| SERPINB5 | 1.959176173 | 2.47E-11 |
| CDC20B | 1.959501848 | 1.27E-13 |
| DRD2 | 1.959619268 | 5.23E-22 |
| CAPN13 | 1.969234963 | 4.75E-34 |
| DNAI1 | 1.970918093 | 1.11E-13 |
| MLIP | 1.971902105 | 5.52E-28 |
| KLK13 | 1.972261898 | 6.31E-18 |
| JCHAIN | 1.975823878 | 1.21E-22 |
| SUGCT | 1.976964968 | 1.62E-62 |
| LKAAEAR1 | 1.978781419 | 5.11E-12 |
| CR2 | 1.979458922 | 5.42E-30 |
| C1QTNF8 | 1.983378252 | 9.69E-10 |
| GRHL3 | 1.985373904 | 1.12E-32 |
| BPIFB4 | 1.987410724 | 5.15E-08 |
| DMRTA2 | 1.991135119 | 4.60E-06 |
| PLPP2 | 1.991455108 | 4.35E-32 |
| SERPINB7 | 1.992505224 | 4.91E-09 |
| CFAP100 | 1.994340466 | 1.45E-14 |
| KIAA2012 | 1.997218748 | 1.65E-16 |
| ITIH6 | 1.997920029 | 8.42E-06 |
| FAM3D | 1.997926929 | 6.04E-18 |
| CDH2 | 1.998779603 | 8.94E-28 |
| CPNE5 | 1.998793496 | 2.13E-59 |
| ADCYAP1 | 1.998874369 | 7.56E-14 |
| IL13 | 1.998900845 | 7.56E-12 |
| ZNF385D | 2.004716512 | 2.50E-22 |
| BMP15 | 2.007944355 | 2.34E-10 |
| FAM163A | 2.010897945 | 9.71E-22 |
| AMPD1 | 2.013490921 | 3.92E-20 |
| PLCH2 | 2.014171567 | 1.35E-35 |
| FUT2 | 2.014199821 | 4.41E-37 |
| AC010255.3 | 2.01594755 | 1.08E-11 |
| FAM155B | 2.019428591 | 1.55E-27 |
| ZNF648 | 2.024190433 | 1.82E-07 |
| UNC93A | 2.025106198 | 0.004837303 |
| NELL1 | 2.025249115 | 9.78E-20 |
| BBOX1 | 2.029601756 | 5.92E-22 |
| TDO2 | 2.030693688 | 1.83E-16 |
| PRSS1 | 2.038177912 | 8.29E-13 |
| ACBD7 | 2.043340725 | 0.002580105 |
| COL22A1 | 2.047162606 | 2.17E-43 |
| CP | 2.048036015 | 1.60E-36 |
| DUSP9 | 2.052814957 | 1.95E-19 |
| TSPAN1 | 2.056734718 | 2.21E-25 |
| TMEM270 | 2.057510848 | 9.97E-07 |
| ST8SIA2 | 2.058544888 | 3.10E-06 |
| SLC2A5 | 2.062672617 | 4.79E-44 |
| MCIDAS | 2.063560091 | 3.36E-12 |
| CXCL14 | 2.063996655 | 5.98E-24 |
| PDLIM4 | 2.064063668 | 2.77E-44 |
| PAX5 | 2.064270986 | 2.14E-09 |
| TXNDC8 | 2.065855618 | 9.24E-05 |
| CKMT1B | 2.068212718 | 6.04E-30 |
| FRMPD2 | 2.069072738 | 1.21E-13 |
| ADH7 | 2.07195375 | 9.02E-10 |
| IL17REL | 2.072435575 | 7.55E-24 |
| HHLA2 | 2.074549772 | 1.39E-22 |
| FHL2 | 2.075530028 | 1.16E-76 |
| KRTAP29-1 | 2.078970366 | 2.54E-08 |
| KIF26B | 2.079980598 | 3.30E-45 |
| TEX11 | 2.081493885 | 2.33E-18 |
| FER1L6 | 2.082127952 | 7.27E-14 |
| CYP2F1 | 2.087268636 | 2.77E-14 |
| DOK5 | 2.093159782 | 1.48E-34 |
| LCN8 | 2.098112983 | 4.68E-05 |
| KLHL40 | 2.099358093 | 0.000132737 |
| MB | 2.09987459 | 2.16E-14 |
| GTSF1L | 2.101275728 | 6.70E-10 |
| PCP4 | 2.107573178 | 4.96E-06 |
| FAM83D | 2.108723488 | 1.53E-07 |
| RAB3C | 2.112508521 | 1.30E-32 |
| KRT13 | 2.112945964 | 1.08E-21 |
| BDKRB2 | 2.115371428 | 2.44E-40 |
| AL355102.2 | 2.122831351 | 4.23E-20 |
| ADAM12 | 2.123385129 | 2.27E-37 |
| SIX1 | 2.124455345 | 2.23E-22 |
| CPNE4 | 2.125039526 | 6.09E-46 |
| PITX1 | 2.127563286 | 3.10E-06 |
| ATP6V1G3 | 2.133448102 | 0.007079605 |
| CCNO | 2.136445931 | 2.30E-14 |
| IGSF9 | 2.141440156 | 9.78E-44 |
| EPHA10 | 2.143876853 | 4.61E-27 |
| CCNA1 | 2.146479566 | 1.52E-19 |
| TAS2R1 | 2.151176165 | 0.000721949 |
| KCNN3 | 2.155150501 | 9.31E-35 |
| ASB5 | 2.161423737 | 5.26E-21 |
| GSTA1 | 2.162986044 | 2.88E-08 |
| BARX2 | 2.167117052 | 4.56E-28 |
| SAA2-SAA4 | 2.167369858 | 1.57E-10 |
| GLYATL2 | 2.171758861 | 3.54E-13 |
| GPR142 | 2.175167346 | 1.01E-20 |
| KRTAP3-2 | 2.178116532 | 4.13E-05 |
| DEFB124 | 2.18063657 | 1.16E-25 |
| POSTN | 2.186373997 | 4.80E-42 |
| VTCN1 | 2.187880838 | 4.77E-16 |
| LRRTM1 | 2.189835085 | 6.32E-18 |
| SERPINB11 | 2.195963696 | 1.71E-09 |
| DAZL | 2.204669303 | 1.04E-11 |
| THY1 | 2.205118112 | 1.15E-33 |
| C1orf232 | 2.205237407 | 0.0065694 |
| USH1C | 2.208579818 | 2.24E-19 |
| ALDH1A3 | 2.20870557 | 4.01E-48 |
| KRTAP3-3 | 2.213973673 | 0.008592845 |
| CD19 | 2.218757224 | 4.56E-30 |
| TUBB3 | 2.21940641 | 6.05E-40 |
| COL3A1 | 2.225264268 | 8.93E-35 |
| SPP1 | 2.227818272 | 6.13E-14 |
| NETO1 | 2.229963602 | 9.07E-15 |
| CLDN14 | 2.236925659 | 3.02E-28 |
| AQP5 | 2.245797694 | 4.64E-26 |
| SYT8 | 2.252652438 | 2.87E-33 |
| RNF225 | 2.254693017 | 8.37E-23 |
| AKR1D1 | 2.26155365 | 6.53E-35 |
| COL14A1 | 2.263150422 | 1.16E-50 |
| NPFFR2 | 2.265461432 | 1.15E-22 |
| FUT6 | 2.269708295 | 1.05E-25 |
| PROM2 | 2.272556975 | 1.18E-50 |
| KCNK2 | 2.28204215 | 4.17E-27 |
| CRB2 | 2.285432591 | 3.02E-06 |
| TWIST1 | 2.286988476 | 5.46E-10 |
| CYP27C1 | 2.291920955 | 9.18E-61 |
| B3GNT6 | 2.294463924 | 2.89E-18 |
| AFP | 2.296842534 | 3.83E-12 |
| COL7A1 | 2.298294958 | 6.90E-69 |
| AC020922.1 | 2.308840845 | 3.83E-09 |
| CLDN25 | 2.309362475 | 0.004215976 |
| SCG5 | 2.319123099 | 5.88E-39 |
| COL1A1 | 2.320833676 | 3.17E-37 |
| TEX26 | 2.336943094 | 7.42E-13 |
| KRT20 | 2.342606582 | 5.39E-07 |
| ADGRF4 | 2.360601915 | 6.47E-40 |
| SPRR5 | 2.365425345 | 9.87E-12 |
| DNAJC22 | 2.365587119 | 2.86E-24 |
| MUCL1 | 2.366558423 | 1.17E-08 |
| WT1 | 2.379599514 | 2.50E-11 |
| OPN4 | 2.395400443 | 1.25E-06 |
| PLA2G2A | 2.398170678 | 1.01E-17 |
| TP63 | 2.398207903 | 1.18E-25 |
| MS4A1 | 2.398534924 | 8.03E-30 |
| MMP11 | 2.403439529 | 2.98E-37 |
| METTL11B | 2.405435496 | 0.00024483 |
| CXCL6 | 2.40554433 | 1.99E-08 |
| SCGB3A1 | 2.419177213 | 4.21E-27 |
| LIPF | 2.427607023 | 2.29E-10 |
| SLC4A11 | 2.431169711 | 8.39E-54 |
| CCL7 | 2.431214893 | 5.60E-14 |
| DPEP1 | 2.432556215 | 2.69E-44 |
| FCRL4 | 2.438352449 | 7.43E-09 |
| PAX9 | 2.445808541 | 5.85E-30 |
| VSIG10L2 | 2.452884027 | 5.89E-19 |
| CTXN3 | 2.474034984 | 1.50E-08 |
| VSIG1 | 2.47454553 | 1.35E-25 |
| CTHRC1 | 2.475034001 | 9.75E-65 |
| GUCA1A | 2.478296054 | 1.13E-17 |
| C11orf86 | 2.48324808 | 7.48E-09 |
| HS6ST2 | 2.495280752 | 1.27E-49 |
| CCKAR | 2.497375499 | 5.31E-09 |
| NXPE4 | 2.507053831 | 8.48E-33 |
| OTX1 | 2.508886892 | 1.07E-23 |
| PROM1 | 2.509677324 | 1.33E-20 |
| OR4N2 | 2.51784204 | 0.006694818 |
| SSTR5 | 2.520661768 | 9.02E-09 |
| LRRC26 | 2.523635207 | 9.86E-16 |
| GIP | 2.52498358 | 4.53E-05 |
| CDH3 | 2.525014473 | 1.49E-64 |
| SMIM31 | 2.527909285 | 1.01E-28 |
| TMEM59L | 2.533454118 | 3.25E-29 |
| TSHR | 2.544981886 | 2.10E-40 |
| GABRA5 | 2.545419572 | 2.71E-06 |
| IL36RN | 2.545557355 | 2.74E-06 |
| MMP3 | 2.546722251 | 6.35E-07 |
| DIO2 | 2.546736289 | 7.69E-38 |
| OGDHL | 2.549617923 | 5.32E-27 |
| UGT1A5 | 2.550126281 | 0.000891663 |
| CGB8 | 2.560024134 | 0.000113935 |
| MUC16 | 2.561174633 | 3.33E-15 |
| MEOX1 | 2.563704384 | 9.42E-30 |
| TRIM49 | 2.564179825 | 1.08E-05 |
| CIDEC | 2.567664059 | 2.95E-19 |
| FETUB | 2.571972263 | 1.76E-10 |
| OR4C6 | 2.572515039 | 4.23E-08 |
| GCNT3 | 2.572869664 | 1.03E-42 |
| SLN | 2.573621868 | 1.82E-13 |
| DERL3 | 2.576859939 | 4.35E-50 |
| PGLYRP4 | 2.581688755 | 2.39E-15 |
| TMEM229A | 2.582429434 | 1.73E-27 |
| MUC13 | 2.5856971 | 2.15E-20 |
| SPRR3 | 2.586521753 | 0.027230791 |
| SPRR2B | 2.59349261 | 0.007384737 |
| PNOC | 2.59560404 | 8.58E-49 |
| IL13RA2 | 2.596573998 | 1.65E-24 |
| TFAP2A | 2.596921503 | 3.15E-33 |
| ADAMTS16 | 2.600424819 | 9.39E-37 |
| KRT17 | 2.600888402 | 3.22E-22 |
| SYT12 | 2.603149709 | 2.20E-61 |
| OCSTAMP | 2.612375092 | 1.82E-13 |
| FCRL5 | 2.613911568 | 2.85E-73 |
| PDE10A | 2.616265432 | 1.43E-34 |
| IL11 | 2.618082594 | 8.69E-38 |
| ECEL1 | 2.631101975 | 2.01E-14 |
| TNFRSF13C | 2.659720131 | 6.92E-43 |
| CLCA4 | 2.661877263 | 1.22E-23 |
| TTR | 2.663906391 | 2.75E-14 |
| PLEKHS1 | 2.682007012 | 1.73E-22 |
| CLDN2 | 2.685141834 | 8.70E-35 |
| TP53AIP1 | 2.686743177 | 4.57E-25 |
| GRM7 | 2.700603171 | 1.85E-20 |
| ADAMTS14 | 2.704657999 | 1.31E-18 |
| UGT1A4 | 2.710455522 | 1.92E-13 |
| SHISAL2B | 2.710558106 | 6.84E-20 |
| MUC4 | 2.714332616 | 1.58E-42 |
| DMRT3 | 2.735414876 | 0.000501753 |
| GJB5 | 2.756926454 | 1.12E-37 |
| CD79A | 2.761694423 | 4.18E-44 |
| CST4 | 2.788508519 | 0.000728711 |
| GAP43 | 2.789543942 | 9.61E-56 |
| FAM83F | 2.790668371 | 7.86E-22 |
| MZB1 | 2.792836861 | 1.41E-51 |
| C6orf15 | 2.793446221 | 3.03E-09 |
| FCRLA | 2.807132023 | 6.79E-56 |
| TNS4 | 2.847893537 | 3.33E-37 |
| PADI1 | 2.862071684 | 9.43E-16 |
| IGLL5 | 2.866928692 | 1.16E-42 |
| HOXB1 | 2.869062918 | 7.68E-12 |
| BNC1 | 2.89136138 | 5.43E-32 |
| DRD5 | 2.892579785 | 1.41E-51 |
| SMR3A | 2.902853143 | 0.000190887 |
| IL22RA2 | 2.91474139 | 2.92E-19 |
| KRTAP1-3 | 2.92343553 | 3.89E-06 |
| ATP12A | 2.925870621 | 8.57E-24 |
| UPK1B | 2.949742609 | 4.00E-20 |
| B3GNT3 | 2.966893419 | 9.76E-51 |
| ERN2 | 2.991624764 | 7.56E-41 |
| CILP2 | 2.997433037 | 6.16E-51 |
| UGT1A10 | 3.010299876 | 1.07E-06 |
| UGT1A8 | 3.01186101 | 3.01E-05 |
| BPIFA1 | 3.018219838 | 1.05E-12 |
| SHISA8 | 3.027903921 | 1.42E-33 |
| STRA6 | 3.028248949 | 1.57E-53 |
| SLCO1B3 | 3.050350416 | 2.44E-28 |
| AC136428.1 | 3.053043289 | 2.20E-16 |
| FOXE1 | 3.071250296 | 9.16E-26 |
| KRT6A | 3.102206509 | 2.50E-11 |
| COMP | 3.111302742 | 6.18E-51 |
| KRT15 | 3.133201971 | 7.68E-47 |
| PGLYRP3 | 3.179516885 | 2.36E-19 |
| BAAT | 3.180399802 | 7.70E-57 |
| NGB | 3.182128483 | 3.06E-16 |
| IFNE | 3.220321538 | 4.46E-41 |
| LGALS7 | 3.221764295 | 5.95E-23 |
| MUC5AC | 3.249538492 | 1.38E-15 |
| MUC2 | 3.26154086 | 9.70E-11 |
| SERPINB13 | 3.265786343 | 7.77E-15 |
| COL10A1 | 3.315392146 | 6.40E-56 |
| FAM83A | 3.346678762 | 7.10E-70 |
| COL17A1 | 3.371053907 | 2.16E-46 |
| BPIFB1 | 3.383405766 | 6.19E-32 |
| GREM1 | 3.406160298 | 2.96E-28 |
| SLC5A5 | 3.439790084 | 2.77E-10 |
| MUC5B | 3.457636391 | 1.65E-32 |
| MOGAT2 | 3.461326912 | 9.76E-19 |
| UGT1A6 | 3.482335907 | 1.70E-31 |
| FAT2 | 3.502011962 | 8.30E-56 |
| MSMB | 3.507209425 | 3.38E-10 |
| TMPRSS4 | 3.558698062 | 3.09E-59 |
| SERPINB4 | 3.571501318 | 8.63E-18 |
| UGT1A3 | 3.668181382 | 8.01E-09 |
| DSC3 | 3.707662002 | 4.76E-24 |
| MMP13 | 3.719028834 | 1.68E-28 |
| CALML3 | 3.769776632 | 1.05E-23 |
| ADAMTS18 | 3.793577468 | 9.74E-20 |
| LCE1C | 3.806491122 | 8.83E-09 |
| KRT5 | 3.842560252 | 1.12E-45 |
| S100A2 | 3.844958412 | 4.94E-78 |
| KRT14 | 3.86220687 | 5.51E-39 |
| CLDN22 | 3.873379388 | 0.001511333 |
| KRTAP2-3 | 3.884491971 | 1.08E-18 |
| LGALS7B | 3.970666253 | 3.43E-41 |
| PADI3 | 4.09580994 | 2.41E-23 |
| GPR87 | 4.126632536 | 1.29E-53 |
| CYP24A1 | 4.222457168 | 1.29E-49 |
| KLK6 | 4.251451775 | 1.16E-35 |
| LY6D | 4.282250331 | 1.45E-24 |
| CXCL13 | 4.286871034 | 2.20E-52 |
| IGFL2 | 4.299628226 | 7.58E-106 |
| FDCSP | 4.816816335 | 1.36E-36 |
| PRSS2 | 4.838913197 | 1.29E-49 |
| CST1 | 5.784714465 | 5.50E-37 |
| SPRR1A | 6.614717204 | 9.35E-49 |

Log2FC denotes log2 fold change in expression of proteins in IPF over HS

**Table S4. Differentially expressed genes identified in comparison of transcriptome profiles of lung tissues from IPF and HS**

| **Gene/protein name** | **Log2 Fold change** | |
| --- | --- | --- |
|  | **Plasma EV Protein** | **Lung tissue RNA** |
| ARG1 | -1.224671576 | -2.006920952 |
| MME | -1.811411479 | -1.735272366 |
| LCN1 | -1.53072172 | -1.333886181 |
| SULT2B1 | -1.543917032 | -1.180363312 |
| TFPI | -1.938021551 | -0.729460635 |
| HMCN1 | 3.430668097 | 0.587352244 |
| HSPG2 | 3.335028031 | 0.59745081 |
| CHL1 | 3.065000969 | 0.63931514 |
| SELP | 2.418575182 | 0.640569755 |
| ANGPTL6 | 5.424441222 | 0.681071006 |
| FBLN1 | 1.557436188 | 0.684280729 |
| PFKP | 1.596222092 | 0.753715353 |
| FLG2 | 1.673914277 | 0.794245963 |
| PKP1 | 2.11119167 | 0.904877659 |
| FBN1 | 4.855105716 | 0.929905939 |
| NR0B1 | 4.150251825 | 0.95230385 |
| PRG4 | 1.009141731 | 0.952518662 |
| TNC | 2.137049461 | 1.003265567 |
| DSP | 1.799033411 | 1.055449113 |
| AMY1A | 2.351644796 | 1.108623093 |
| SSC5D | 1.844957099 | 1.436239858 |
| SBSN | 5.211991792 | 1.466871555 |
| DCDC1 | 1.962198005 | 1.469134848 |
| CALML5 | 2.705746557 | 1.527556336 |
| PRR4 | 3.044957398 | 1.606125647 |
| LTF | 1.853745453 | 1.947692841 |
| CKMT1A | 4.084069522 | 1.957618719 |
| FETUB | 1.474320737 | 2.571972263 |
| SPRR3 | 3.051918908 | 2.586521753 |
| TTR | 1.257762323 | 2.663906391 |
| CST4 | 2.393102066 | 2.788508519 |
| COMP | 2.627172139 | 3.111302742 |
| LGALS7 | 1.744686081 | 3.221764295 |
| MUC5B | 2.630800453 | 3.457636391 |
| CALML3 | 1.933699484 | 3.769776632 |
| LGALS7B | 1.744686081 | 3.970666253 |

**Table S5A: Pathways over represented in IPF plasma EV proteome compared to healthy plasma EV proteome**

| **Pathway category** | **Pathway term name** | **Padj** | **term_size** | **Intersections** |
| --- | --- | --- | --- | --- |
| ECM/collagen formation | ECM proteoglycans | 0.001987956 | 75 | ACAN,LAMC1,TNC,ITGA2B,VTN,COMP,TNXB,HSPG2 |
|  | Extracellular matrix organization | 0.002578374 | 298 | ACAN,CD47,PPIB,TTR,FBLN1,LAMC1,TNC,ITGA2B,ACTN1,VTN,COMP,TNXB,CAPN1,HSPG2,FBN1 |
|  | Integrin cell surface interactions | 0.039387346 | 84 | CD47,TNC,ITGA2B,VTN,COMP,HSPG2,FBN1 |
|  | Non-integrin membrane-ECM interactions | 0.037549069 | 58 | TTR,LAMC1,TNC,ACTN1,VTN,HSPG2 |
| keratinization | Formation of the cornified envelope | 3.52E-07 | 128 | DSC3,DSC1,JUP,DSP,PKP1,CSTA,EVPL,CASP14,FLG,IVL,CAPN1,SPRR3,TGM1,LORICRIN |
|  | Keratinization | 0.000262238 | 215 | DSC3,DSC1,JUP,DSP,PKP1,CSTA,EVPL,CASP14,FLG,IVL,CAPN1,SPRR3,TGM1,LORICRIN |
| Pulmonary surfactant metabolism dysfunction | Defective CSF2RB causes SMDP5 | 0.027518006 | 7 | SFTPD,SFTPA1,SFTPB |
|  | Defective CSF2RA causes SMDP4 | 0.027518006 | 7 | SFTPD,SFTPA1,SFTPB |
| Neutrophil degranulation | Neutrophil degranulation | 7.72E-18 | 474 | CAB39,GGH,CD47,LGALS3,PYCARD,MME,ARG1,PRTN3,CAMP,DSC1,FCN1,TTR,PFKL,JUP,FLG2,DSP,LTF,AMPD3,PKP1,BIN2,SERPINA1,CREG1,GSN,RAP1B,CFP,GM2A,CALML5,PTPRJ,ASAH1,CAPN1,CSTB,CAP1,HMGB1,HRNR,VCL,PNP,ANPEP,ALDOA |
|  | Innate Immune System | 5.43E-15 | 1073 | CAB39,GGH,CD47,LGALS3,PYCARD,RAC2,MME,PRKCSH,ARG1,PRTN3,COLEC10,CAMP,DSC1,FCN1,MBL2,TTR,LBP,PFKL,JUP,SFTPD,FLG2,MUC7,DSP,LTF,AMPD3,PKP1,BIN2,FCN2,SERPINA1,CREG1,GSN,RAP1B,CFP,VTN,MUC5B,GM2A,SFTPA1,CALML5,PTPRJ,CFL1,ASAH1,CAPN1,CSTB,CAP1,HMGB1,CARD9,HRNR,VCL,PNP,ANPEP,ALDOA |
|  | Immune System | 5.73E-08 | 2116 | CAB39,GGH,CD47,LGALS3,PYCARD,RAC2,MME,PTPRZ1,PRKCSH,ARG1,PRTN3,COLEC10,CAMP,DSC1,FCN1,MBL2,TTR,LBP,PFKL,AP1B1,JUP,SFTPD,FLG2,MUC7,DSP,LTF,CALR,AMPD3,PKP1,CD226,ITGA2B,BIN2,FCN2,SERPINA1,CREG1,GSN,RAP1B,BLMH,CFP,VTN,MUC5B,GM2A,SFTPA1,CALML5,IL36G,PTPRJ,CFL1,ASAH1,CAPN1,STAT3,CSTB,CAP1,HMGB1,VIM,CARD9,HRNR,VCL,PNP,ANPEP,ALDOA |
| Platelet degranulation | Platelet degranulation | 2.23E-08 | 125 | ANXA5,LGALS3BP,SERPINF2,TTN,PFN1,ITGA2B,SERPINA1,SELP,ACTN1,CFL1,ACTN4,CAP1,VCL,FERMT3,ALDOA |
|  | Response to elevated platelet cytosolic Ca2+ | 3.94E-08 | 130 | ANXA5,LGALS3BP,SERPINF2,TTN,PFN1,ITGA2B,SERPINA1,SELP,ACTN1,CFL1,ACTN4,CAP1,VCL,FERMT3,ALDOA |
|  | Platelet activation, signaling and aggregation | 2.00E-06 | 258 | ANXA5,RAC2,GNAI2,LGALS3BP,SERPINF2,TTN,PFN1,ITGA2B,SERPINA1,RAP1B,SELP,ACTN1,CFL1,ACTN4,CAP1,VCL,FERMT3,ALDOA |
|  | Hemostasis | 0.01271222 | 657 | ANXA5,CD47,RAC2,TFPI,GNAI2,PRTN3,LGALS3BP,SERPINF2,TTN,PFN1,ITGA2B,SERPINA1,RAP1B,SELP,ACTN1,SELPLG,CFL1,ACTN4,CAP1,VCL,FERMT3,ALDOA |
|  | Lectin pathway of complement activation | 0.000350361 | 7 | COLEC10,FCN1,MBL2,FCN2 |
|  | Regulation of TLR by endogenous ligand | 0.034390983 | 19 | LBP,SFTPD,SFTPA1,HMGB1 |

**Table S5B: Pathways over represented in IPF lung transcriptome compared to healthy lung transcriptome**

| **Pathway category** | **Pathway term name** | **padj** | **term_size** | **Intersections** |
| --- | --- | --- | --- | --- |
| ECM/collagen formation | Collagen formation | 4.64E-12 | 89 | COL7A1,COL10A1,COL14A1,COL17A1,COL22A1,ITGB4,COL1A1,COL3A1,LAMA3,COL6A3,MMP7,P3H2,MMP13,COL1A2,COL4A6,COL16A1,P3H3,COL15A1,COL5A1,P4HA3,COL18A1,ADAMTS14,LOXL1,COL4A3,COL6A1,COL4A5,COL9A3,COL6A2,COL23A1,COL11A1,COL25A1,COL28A1,PCOLCE,COL9A2,LOXL2,COL6A6,COL5A2,MMP9,MMP3,TLL2,COL9A1,LOX,CTSL,COL24A1,PCOLCE2,COL20A1,ADAMTS3 |
|  | Degradation of the extracellular matrix | 1.23E-11 | 140 | COL7A1,COL10A1,COL14A1,PRSS2,COL17A1,MMP11,COL1A1,ADAMTS16,COL3A1,CAPN13,LAMA3,COL6A3,MMP7,MMP13,COL1A2,COL4A6,COL16A1,MMP2,COL15A1,COL5A1,ADAMTS18,ACAN,COL18A1,COL4A3,FBN1,HTRA1,CAPN11,COL6A1,COL4A5,SPP1,COL9A3,CAPN5,COL6A2,PRSS1,SPOCK3,COL23A1,COL11A1,COL25A1,CAPNS2,FBN3,ADAMTS9,MMP12,ADAMTS8,COL9A2,HSPG2,TIMP1,COL6A6,CMA1,COL5A2,MMP9,MMP3,TLL2,TPSAB1,MMP25,COL9A1,CTSK,CTSL,CAPN9,MMP8,CTRB1,CTSG,ADAMTS4 |
|  | Assembly of collagen fibrils and other multimeric structures | 2.44E-10 | 60 | COL7A1,COL10A1,COL14A1,COL17A1,ITGB4,COL1A1,COL3A1,LAMA3,COL6A3,MMP7,MMP13,COL1A2,COL4A6,COL15A1,COL5A1,COL18A1,LOXL1,COL4A3,COL6A1,COL4A5,COL9A3,COL6A2,COL11A1,PCOLCE,COL9A2,LOXL2,COL6A6,COL5A2,MMP9,MMP3,TLL2,COL9A1,LOX,CTSL,COL24A1 |
|  | Collagen chain trimerization | 2.95E-10 | 44 | COL7A1,COL10A1,COL14A1,COL17A1,COL22A1,COL1A1,COL3A1,COL6A3,COL1A2,COL4A6,COL16A1,COL15A1,COL5A1,COL18A1,COL4A3,COL6A1,COL4A5,COL9A3,COL6A2,COL23A1,COL11A1,COL25A1,COL28A1,COL9A2,COL6A6,COL5A2,COL9A1,COL24A1,COL20A1 |
|  | Collagen degradation | 5.16E-10 | 64 | COL7A1,COL10A1,COL14A1,PRSS2,COL17A1,MMP11,COL1A1,COL3A1,COL6A3,MMP7,MMP13,COL1A2,COL4A6,COL16A1,MMP2,COL15A1,COL5A1,COL18A1,COL4A3,COL6A1,COL4A5,COL9A3,COL6A2,COL23A1,COL11A1,COL25A1,MMP12,COL9A2,COL6A6,COL5A2,MMP9,MMP3,COL9A1,CTSK,CTSL,MMP8 |
|  | Collagen biosynthesis and modifying enzymes | 5.38E-10 | 67 | COL7A1,COL10A1,COL14A1,COL17A1,COL22A1,COL1A1,COL3A1,COL6A3,P3H2,COL1A2,COL4A6,COL16A1,P3H3,COL15A1,COL5A1,P4HA3,COL18A1,ADAMTS14,COL4A3,COL6A1,COL4A5,COL9A3,COL6A2,COL23A1,COL11A1,COL25A1,COL28A1,PCOLCE,COL9A2,COL6A6,COL5A2,TLL2,COL9A1,COL24A1,PCOLCE2,COL20A1,ADAMTS3 |
|  | Extracellular matrix organization | 1.90E-17 | 298 | COL7A1,COL10A1,COMP,COL14A1,PRSS2,COL17A1,COL22A1,ITGB4,ADAM12,MMP11,COL1A1,ADAMTS16,COL3A1,CAPN13,LTBP1,LAMA3,COL6A3,MMP7,P3H2,MMP13,COL1A2,ITGA7,FBLN2,COL4A6,COL16A1,MFAP2,MMP2,P3H3,ASPN,COL15A1,COL5A1,P4HA3,ICAM2,ADAMTS18,ACAN,COL18A1,ADAMTS14,ITGA11,TNN,LOXL1,COL4A3,SH3PXD2A,FBN1,TGFB3,HTRA1,LAMC3,CAPN11,FBLN5,COL6A1,COL4A5,TTR,SPP1,COL9A3,MATN3,CAPN5,ITGA2B,COL6A2,PDGFB,PRSS1,VCAN,SPOCK3,COL23A1,COL11A1,COL25A1,LRP4,VCAM1,ITGB3,NCAM1,BMP7,CAPNS2,FBN3,ADAMTS9,ICAM1,TNC,MMP12,COL28A1,PCOLCE,ADAMTS8,COL9A2,LUM,LOXL2,ITGAL,CEACAM6,SPARC,HSPG2,ITGA10,DMP1,TIMP1,COL6A6,CMA1,COL5A2,FBLN1,BGN,MMP9,MMP3,TLL2,TPSAB1,MMP25,COL9A1,CTSK,LOX,CTSL,CEACAM8,COL24A1,PCOLCE2,NRXN1,CAPN9,COL20A1,ADAMTS3,MMP8,CTRB1,HAPLN1,CTSG,MFAP1,FGG,ADAMTS4 |
|  | Integrin cell surface interactions | 9.76E-06 | 84 | COL7A1,COL10A1,COMP,COL1A1,COL3A1,COL6A3,COL1A2,ITGA7,COL4A6,COL16A1,COL5A1,ICAM2,COL18A1,ITGA11,COL4A3,FBN1,COL6A1,COL4A5,SPP1,COL9A3,ITGA2B,COL6A2,COL23A1,VCAM1,ITGB3,ICAM1,TNC,COL9A2,LUM,ITGAL,HSPG2,ITGA10,COL6A6,COL5A2,COL9A1,FGG |
|  | ECM proteoglycans | 9.86E-07 | 75 | COMP,COL1A1,COL3A1,LAMA3,COL6A3,COL1A2,ITGA7,COL4A6,ASPN,COL5A1,ACAN,TNN,COL4A3,TGFB3,COL6A1,COL4A5,COL9A3,MATN3,ITGA2B,COL6A2,VCAN,LRP4,ITGB3,NCAM1,TNC,COL9A2,LUM,SPARC,HSPG2,DMP1,COL6A6,COL5A2,BGN,COL9A1,HAPLN1 |
|  | NCAM1 interactions | 0.034847 | 42 | COL3A1,COL6A3,CACNA1G,CACNA1S,ARTN,COL5A1,COL4A3,COL6A1,COL4A5,COL9A3,COL6A2,NCAM1,GFRA1,COL9A2,COL6A6,COL5A2,ST8SIA2,COL9A1 |
|  | Activation of Matrix Metalloproteinases | 0.000478 | 33 | PRSS2,MMP11,MMP7,MMP13,MMP2,COL18A1,PRSS1,SPOCK3,TIMP1,CMA1,MMP9,MMP3,TPSAB1,MMP25,CTSK,MMP8,CTRB1,CTSG |
| Keratinization | Formation of the cornified envelope | 0.003945 | 128 | SPRR1A,KRT15,KRT5,KRT14,DSC3,KRT17,KRT13,KRT23,KLK13,KRT73,DSP,KRT72,KRT40,KRT32,KRT6A,KRT19,PKP2,KLK5,KLK12,KRT80,LCE1C,KRT8,SPINK5,PKP1,TGM1,KRT20,KRT39,ST14,KRT74,KRT85,KRT78,KRT34,KRT79,KLK14,KRT37,KRT16,SPRR2B,LCE1B,KRT33B,KLK8,SPRR3,KRT71 |
| Cell-cell communication | Cell-Cell communication | 0.00123 | 127 | CDH3,COL17A1,KRT5,ITGB4,KRT14,CLDN2,CDH13,LAMA3,CLDN14,CDH2,CDH5,FLNC,LIMS2,SPTBN1,CLDN5,FBLIM1,CADM2,NECTIN4,CLDN16,CLDN3,CADM1,SIRPB1,NECTIN1,PARD6G,SIRPG,SFTPA1,CDH10,SFTPD,SFTPA2,CLDN1,CLDN18,CLDN19,NECTIN3,CADM3,NPHS1,CLDN8,KIRREL2,SKAP2,FYB1,CLDN10,CLDN22,PARD6B,NPHS2 |
|  | Cell junction organization | 0.009738 | 91 | CDH3,COL17A1,KRT5,ITGB4,KRT14,CLDN2,CDH13,LAMA3,CLDN14,CDH2,CDH5,FLNC,LIMS2,CLDN5,FBLIM1,CADM2,NECTIN4,CLDN16,CLDN3,CADM1,NECTIN1,PARD6G,CDH10,CLDN1,CLDN18,CLDN19,NECTIN3,CADM3,CLDN8,CLDN10,CLDN22,PARD6B |
|  | Cell-cell junction organization | 0.032279 | 64 | CDH3,CLDN2,CDH13,CLDN14,CDH2,CDH5,CLDN5,CADM2,NECTIN4,CLDN16,CLDN3,CADM1,NECTIN1,PARD6G,CDH10,CLDN1,CLDN18,CLDN19,NECTIN3,CADM3,CLDN8,CLDN10,CLDN22,PARD6B |
| Chemokine signaling | Chemokine receptors bind chemokines | 4.69E-05 | 57 | CXCL13,ACKR4,CCR7,CXCL12,PF4,CCL7,CX3CR1,CCRL2,CCR8,CCR6,CXCL6,CCL25,CCL4,CCL21,CCL17,CCL13,CXCL2,CXCL3,CCL1,CCR4,CX3CL1,CCL11,CCL19,CXCR2,CXCR1,CXCL11,CCL27 |
|  | Peptide ligand-binding receptors | 3.41E-14 | 195 | CXCL13,PNOC,EDNRB,BDKRB2,ACKR4,RXFP1,CCK,CCR7,NPFFR2,ACKR1,AVPR1A,AGTR1,CXCL12,PF4,CCL7,OXTR,CX3CR1,MCHR1,C3,MC5R,CCRL2,CCR8,BDKRB1,UTS2R,TACR2,GPR37,CCKAR,GPER1,SSTR5,CCR6,F2RL3,RXFP2,EDN3,CXCL6,CCL25,GRPR,CCL4,CCL21,QRFPR,CCL17,SSTR4,CCL13,NPW,NPSR1,RLN3,INSL5,CXCL2,TACR1,NMBR,OXT,CXCL3,CCL1,NPY2R,RXFP4,UTS2,RXFP3,ANXA1,CCR4,NPB,CX3CL1,NPBWR1,NMU,F2,NTSR1,PRLH,CCL11,CCL19,TAC1,MCHR2,CXCR2,CXCR1,PDYN,CXCL11,NPY1R,CCL27,PROK2,NTS,CCKBR,F2RL2,SSTR2,EDN2,TRH |
| GPCR signaling | GPCR ligand binding | 3.71E-21 | 459 | ADRA1A,CXCL13,DRD5,PNOC,ADRB2,EDNRB,TSHR,BDKRB2,GRM8,WNT7A,HTR2A,ACKR4,CHRM3,RXFP1,CCK,PTHLH,HCAR1,ADRB1,CCR7,VIPR1,NPFFR2,HTR1D,RAMP2,DRD2,FZD8,ADM2,GPBAR1,GNB3,GRM7,S1PR5,ACKR1,PLPPR4,AVPR1A,RAMP3,WNT4,RAMP1,AGTR1,S1PR1,CXCL12,GHRHR,GNG4,GRM4,WNT10A,CHRM2,ADORA2A,PF4,GNG11,CCL7,ADCYAP1,ADRA1D,ADGRE1,OXTR,CX3CR1,P2RY6,SMO,GIPR,P2RY14,MCHR1,PLPPR1,C3,SHH,CALCRL,MC5R,CCRL2,CCR8,PLPPR3,BDKRB1,ADRA2A,HTR4,UTS2R,TACR2,ADORA2B,MTNR1A,GPR37,GPR17,HRH1,CCKAR,PTGDR,GPER1,FFAR4,SSTR5,CCR6,F2RL3,RXFP2,EDN3,CXCL6,CCL25,CALCR,GRPR,CCL4,GRM2,CCL21,LHCGR,FZD3,TAS2R40,VIP,LPAR3,OPN4,CHRM4,WNT1,QRFPR,CCL17,SSTR4,CCL13,NPW,NPSR1,RLN3,INSL5,RGR,CXCL2,ADCYAP1R1,ADRA1B,CALCA,TACR1,NMBR,OXT,ADGRE3,CXCL3,OPN5,CCL1,PLPPR5,CHRM1,GIP,NPY2R,PTGER4,RXFP4,UTS2,FSHR,RXFP3,VIPR2,ADM,ANXA1,CCR4,NPB,PTGER3,CX3CL1,FFAR2,NPBWR1,NMU,TAS2R1,F2,GRM5,NTSR1,TAS2R60,PRLH,CCL11,CCL19,TAC1,TAS2R50,MCHR2,CXCR2,CXCR1,PDYN,CXCL11,NPY1R,CCL27,PROK2,NTS,CCKBR,IAPP,F2RL2,SSTR2,WNT10B,EDN2,TRH |
|  | Class A/1 (Rhodopsin-like receptors) | 1.65E-17 | 328 | ADRA1A,CXCL13,DRD5,PNOC,ADRB2,EDNRB,TSHR,BDKRB2,HTR2A,ACKR4,CHRM3,RXFP1,CCK,HCAR1,ADRB1,CCR7,NPFFR2,HTR1D,DRD2,GPBAR1,S1PR5,ACKR1,PLPPR4,AVPR1A,AGTR1,S1PR1,CXCL12,CHRM2,ADORA2A,PF4,CCL7,ADRA1D,OXTR,CX3CR1,P2RY6,P2RY14,MCHR1,PLPPR1,C3,MC5R,CCRL2,CCR8,PLPPR3,BDKRB1,ADRA2A,HTR4,UTS2R,TACR2,ADORA2B,MTNR1A,GPR37,GPR17,HRH1,CCKAR,PTGDR,GPER1,FFAR4,SSTR5,CCR6,F2RL3,RXFP2,EDN3,CXCL6,CCL25,GRPR,CCL4,CCL21,LHCGR,LPAR3,OPN4,CHRM4,QRFPR,CCL17,SSTR4,CCL13,NPW,NPSR1,RLN3,INSL5,RGR,CXCL2,ADRA1B,TACR1,NMBR,OXT,CXCL3,OPN5,CCL1,PLPPR5,CHRM1,NPY2R,PTGER4,RXFP4,UTS2,FSHR,RXFP3,ANXA1,CCR4,NPB,PTGER3,CX3CL1,FFAR2,NPBWR1,NMU,F2,NTSR1,PRLH,CCL11,CCL19,TAC1,MCHR2,CXCR2,CXCR1,PDYN,CXCL11,NPY1R,CCL27,PROK2,NTS,CCKBR,F2RL2,SSTR2,EDN2,TRH |
|  | Signaling by GPCR | 8.77E-05 | 1158 | ADRA1A,ARRB1,STRA6,CXCL13,DRD5,PNOC,RGS9BP,ADRB2,EDNRB,HSD17B6,ARHGEF26,TSHR,BDKRB2,GRM8,PDE10A,WNT7A,HTR2A,GRK5,ACKR4,CHRM3,APOA1,RXFP1,CCK,ADCY8,ARHGEF10,PTHLH,HCAR1,RBP2,ADRB1,CCR7,DGKI,ABCA4,GNAO1,MGLL,RGS9,VIPR1,NPFFR2,REEP2,HTR1D,RAMP2,DRD2,FZD8,ADM2,GPBAR1,GNB3,PDE1A,GRM7,S1PR5,GPC1,ACKR1,PLPPR4,AVPR1A,RAMP3,NBEA,OBSCN,WNT4,RAMP1,AGTR1,S1PR1,CXCL12,NGEF,GHRHR,CNGA1,GNG4,GRM4,WNT10A,PDE7B,GPC3,CHRM2,RGS22,ADORA2A,PRKCE,ARHGEF2,PF4,TTR,GNG11,RGS6,CCL7,FGD4,RGS4,ADCYAP1,ADRA1D,ADGRE1,OXTR,ADCY2,PREX1,FGD3,PRKCQ,CX3CR1,P2RY6,SMO,PDE1C,GIPR,P2RY14,MCHR1,PLXNB1,GPSM1,PLPPR1,TRPC3,C3,DHRS9,SHH,CALCRL,MC5R,CCRL2,CCR8,PLPPR3,BDKRB1,OR6K3,ADRA2A,HTR4,GNAL,ADCY5,UTS2R,PDE6A,VAV3,PDE2A,TACR2,BCO2,ADORA2B,MTNR1A,OR2W3,GPR37,GPR17,HRH1,ARHGEF4,CCKAR,OR2AG2,PTGDR,GPER1,FFAR4,SSTR5,GUCY2F,CCR6,F2RL3,RXFP2,OR6A2,EDN3,CXCL6,CCL25,OR5P3,OR4C6,HSPG2,RGS18,CALCR,GRPR,LPL,OR6N1,CCL4,GRM2,CCL21,DGKE,OR2A7,PDE4C,LHCGR,MMP3,RASGRF2,FZD3,TAS2R40,VIP,LPAR3,OPN4,CHRM4,WNT1,QRFPR,CCL17,SSTR4,OR5P2,LDLR,APOC3,CCL13,NPW,NPSR1,RLN3,INSL5,RGR,CXCL2,ARHGEF37,HBEGF,RGS20,OR6C6,ADCYAP1R1,ADRA1B,AKR1B10,CALCA,TACR1,NMBR,OXT,ADGRE3,CXCL3,OPN5,RGS1,CCL1,PLPPR5,CHRM1,GIP,NPY2R,PTGER4,DGKB,RXFP4,OR10A2,UTS2,FSHR,RXFP3,VIPR2,LRP2,ADM,ANXA1,CCR4,NPB,PTGER3,CX3CL1,FFAR2,ARHGEF33,NPBWR1,NMU,OR52K2,OR8H1,OR2D3,OR2T10,TAS2R1,OR51E1,F2,OR1J4,GRM5,NTSR1,TAS2R60,PRLH,OR6N2,CCL11,CCL19,TAC1,TAS2R50,MCHR2,OR2D2,CXCR2,OR2S2,CXCR1,APOA2,PDYN,CXCL11,NPY1R,RGSL1,OR4N2,PRKACG,CCL27,OR10A4,PROK2,NTS,CCKBR,IAPP,OR1Q1,OR8H2,F2RL2,SSTR2,WNT10B,REEP6,PRKAR2B,EDN2,TRH |
|  | G alpha (q) signalling events | 0.000538 | 212 | ADRA1A,EDNRB,BDKRB2,HTR2A,GRK5,CHRM3,CCK,DGKI,MGLL,NPFFR2,GNB3,AVPR1A,AGTR1,GNG4,PRKCE,GNG11,RGS4,ADRA1D,OXTR,PRKCQ,P2RY6,MCHR1,TRPC3,BDKRB1,UTS2R,TACR2,GPR17,HRH1,CCKAR,FFAR4,F2RL3,EDN3,RGS18,GRPR,DGKE,MMP3,LPAR3,OPN4,QRFPR,NPSR1,HBEGF,ADRA1B,TACR1,NMBR,OXT,RGS1,CHRM1,DGKB,UTS2,ANXA1,FFAR2,NMU,F2,GRM5,NTSR1,TAC1,MCHR2,RGSL1,PROK2,NTS,CCKBR,F2RL2,EDN2,TRH |
|  | GPCR downstream signalling | 0.003721 | 1088 | ADRA1A,ARRB1,STRA6,CXCL13,DRD5,PNOC,RGS9BP,ADRB2,EDNRB,HSD17B6,ARHGEF26,TSHR,BDKRB2,GRM8,PDE10A,HTR2A,GRK5,CHRM3,APOA1,RXFP1,CCK,ADCY8,ARHGEF10,PTHLH,HCAR1,RBP2,ADRB1,CCR7,DGKI,ABCA4,GNAO1,MGLL,RGS9,VIPR1,NPFFR2,REEP2,HTR1D,RAMP2,ADM2,GPBAR1,GNB3,PDE1A,GRM7,S1PR5,GPC1,AVPR1A,RAMP3,NBEA,OBSCN,RAMP1,AGTR1,S1PR1,CXCL12,NGEF,GHRHR,CNGA1,GNG4,GRM4,PDE7B,GPC3,CHRM2,RGS22,ADORA2A,PRKCE,ARHGEF2,PF4,TTR,GNG11,RGS6,FGD4,RGS4,ADCYAP1,ADRA1D,OXTR,ADCY2,PREX1,FGD3,PRKCQ,CX3CR1,P2RY6,PDE1C,GIPR,P2RY14,MCHR1,PLXNB1,GPSM1,TRPC3,C3,DHRS9,CALCRL,MC5R,CCR8,BDKRB1,OR6K3,ADRA2A,HTR4,GNAL,ADCY5,UTS2R,PDE6A,VAV3,PDE2A,TACR2,BCO2,ADORA2B,MTNR1A,OR2W3,GPR37,GPR17,HRH1,ARHGEF4,CCKAR,OR2AG2,PTGDR,GPER1,FFAR4,SSTR5,GUCY2F,CCR6,F2RL3,RXFP2,OR6A2,EDN3,CXCL6,CCL25,OR5P3,OR4C6,HSPG2,RGS18,CALCR,GRPR,LPL,OR6N1,CCL4,GRM2,CCL21,DGKE,OR2A7,PDE4C,LHCGR,MMP3,RASGRF2,TAS2R40,VIP,LPAR3,OPN4,CHRM4,QRFPR,SSTR4,OR5P2,LDLR,APOC3,CCL13,NPW,NPSR1,RLN3,INSL5,RGR,CXCL2,ARHGEF37,HBEGF,RGS20,OR6C6,ADCYAP1R1,ADRA1B,AKR1B10,CALCA,TACR1,NMBR,OXT,CXCL3,OPN5,RGS1,CCL1,CHRM1,GIP,NPY2R,PTGER4,DGKB,RXFP4,OR10A2,UTS2,FSHR,RXFP3,VIPR2,LRP2,ADM,ANXA1,CCR4,NPB,PTGER3,CX3CL1,FFAR2,ARHGEF33,NPBWR1,NMU,OR52K2,OR8H1,OR2D3,OR2T10,TAS2R1,OR51E1,F2,OR1J4,GRM5,NTSR1,TAS2R60,OR6N2,CCL19,TAC1,TAS2R50,MCHR2,OR2D2,CXCR2,OR2S2,CXCR1,APOA2,PDYN,CXCL11,NPY1R,RGSL1,OR4N2,PRKACG,CCL27,OR10A4,PROK2,NTS,CCKBR,IAPP,OR1Q1,OR8H2,F2RL2,SSTR2,REEP6,PRKAR2B,EDN2,TRH |
|  | G alpha (i) signalling events | 7.14E-06 | 401 | STRA6,CXCL13,PNOC,RGS9BP,HSD17B6,BDKRB2,GRM8,APOA1,ADCY8,HCAR1,RBP2,CCR7,ABCA4,GNAO1,RGS9,HTR1D,GNB3,PDE1A,GRM7,S1PR5,GPC1,NBEA,S1PR1,CXCL12,CNGA1,GNG4,GRM4,GPC3,CHRM2,RGS22,PF4,TTR,GNG11,RGS6,RGS4,ADCY2,PRKCQ,CX3CR1,PDE1C,P2RY14,MCHR1,GPSM1,C3,DHRS9,CCR8,BDKRB1,ADRA2A,GNAL,ADCY5,PDE6A,BCO2,MTNR1A,GPR37,GPR17,GPER1,SSTR5,GUCY2F,CCR6,CXCL6,CCL25,HSPG2,RGS18,LPL,CCL4,GRM2,CCL21,PDE4C,TAS2R40,LPAR3,CHRM4,SSTR4,LDLR,APOC3,CCL13,NPW,RLN3,INSL5,RGR,CXCL2,RGS20,AKR1B10,CXCL3,OPN5,RGS1,CCL1,NPY2R,RXFP4,RXFP3,LRP2,ANXA1,CCR4,NPB,PTGER3,CX3CL1,NPBWR1,NMU,TAS2R1,TAS2R60,CCL19,TAS2R50,MCHR2,CXCR2,CXCR1,APOA2,PDYN,CXCL11,NPY1R,RGSL1,PRKACG,CCL27,SSTR2,PRKAR2B |
| Mucin dysregulation | Defective GALNT12 causes colorectal cancer 1 (CRCS1) | 0.028235 | 16 | MUC4,MUC5B,MUC12,MUC20,MUC13,MUC5AC,MUC16,MUCL1,MUC21,MUC3A |
|  | Defective GALNT3 causes familial hyperphosphatemic tumoral calcinosis (HFTC) | 0.028235 | 16 | MUC4,MUC5B,MUC12,MUC20,MUC13,MUC5AC,MUC16,MUCL1,MUC21,MUC3A |
|  | O-linked glycosylation | 0.001272 | 107 | B3GNT3,ST6GAL1,GCNT3,MUC4,ADAMTS16,MUC5B,THBS2,MUC12,MUC20,THSD1,LARGE2,SEMA5A,MUC13,ADAMTS18,ADAMTS14,CHST4,B3GNT8,GALNT13,GALNT6,MUC5AC,MUC16,ADAMTSL3,ADAMTSL1,ADAMTS7,GALNT9,ADAMTS9,ADAMTS8,MUCL1,ST6GALNAC3,CFP,MUC21,GALNT14,GALNT18,MUC3A,B3GNT7,ADAMTS3,THSD7B,ADAMTS4 |
|  | Diseases associated with O-glycosylation of proteins | 0.004203 | 65 | MUC4,ADAMTS16,NOTCH4,MUC5B,THBS2,MUC12,MUC20,THSD1,SEMA5A,MUC13,ADAMTS18,ADAMTS14,MUC5AC,MUC16,ADAMTSL3,ADAMTSL1,ADAMTS7,ADAMTS9,ADAMTS8,MUCL1,CFP,MUC21,MUC3A,ADAMTS3,THSD7B,ADAMTS4 |
| Post translational modifications | Regulation of Insulin-like Growth Factor (IGF) transport and uptake by Insulin-like Growth Factor Binding Proteins (IGFBPs) | 8.41E-06 | 124 | GOLM1,CP,IGF1,LTBP1,APOA1,C4A,CDH2,IGFBP2,APOA5,WFS1,PROC,MMP2,ENAM,MELTF,PCSK9,KLK13,IGFBP7,FBN1,GPC3,FAM20C,SPP1,MATN3,VCAN,STC2,AFP,C3,BMP15,EVA1A,TF,F5,IGFALS,TNC,DMP1,TIMP1,PAPPA,PAPPA2,GZMH,HRC,MEPE,F2,AMBN,APOA2,KLK3,CTSG,AMTN,NOTUM,FGG |
|  | Post-translational protein phosphorylation | 0.009287 | 107 | GOLM1,CP,LTBP1,APOA1,C4A,CDH2,APOA5,WFS1,PROC,ENAM,MELTF,PCSK9,IGFBP7,FBN1,GPC3,FAM20C,SPP1,MATN3,VCAN,STC2,AFP,C3,BMP15,EVA1A,TF,F5,TNC,DMP1,TIMP1,HRC,MEPE,AMBN,APOA2,AMTN,NOTUM,FGG |
| Calcitonin-like ligand receptors | Calcitonin-like ligand receptors | 0.000479 | 10 | RAMP2,ADM2,RAMP3,RAMP1,CALCRL,CALCR,CALCA,ADM,IAPP |
|  | Class B/2 (Secretin family receptors) | 0.016055 | 93 | WNT7A,PTHLH,VIPR1,RAMP2,FZD8,ADM2,GNB3,RAMP3,WNT4,RAMP1,GHRHR,GNG4,WNT10A,GNG11,ADCYAP1,ADGRE1,SMO,GIPR,SHH,CALCRL,CALCR,FZD3,VIP,WNT1,ADCYAP1R1,CALCA,ADGRE3,GIP,VIPR2,ADM,IAPP,WNT10B |
|  | ADORA2B mediated anti-inflammatory cytokines production | 0.004712 | 133 | DRD5,ADRB2,TSHR,RXFP1,ADCY8,PTHLH,ADRB1,VIPR1,RAMP2,ADM2,GPBAR1,GNB3,RAMP3,RAMP1,GHRHR,GNG4,ADORA2A,GNG11,ADCYAP1,ADCY2,GIPR,CALCRL,MC5R,HTR4,ADCY5,ADORA2B,PTGDR,RXFP2,CALCR,LHCGR,VIP,NPSR1,RLN3,ADCYAP1R1,CALCA,GIP,PTGER4,FSHR,VIPR2,ADM,PRKACG,IAPP,PRKAR2B |
|  | Neuronal System | 0.000166 | 400 | EPB41L5,SYT12,GRIA1,TUBB3,LIN7A,SNAP25,KCNN3,KCND3,UNC13B,MAOA,KCNN4,ADCY8,PPFIBP1,SYN2,KIF17,KCNK2,CACNA2D3,HTR3C,KCNS2,KCNS1,SYN3,GNB3,TUBB1,CACNA2D2,EPB41L2,NRGN,SLC6A4,DLG2,NBEA,LRRTM1,GNG4,SLITRK2,KCNMB4,TSPAN7,GRIN3B,KCNG2,GABRB2,KCNJ16,PPFIA2,GNG11,HTR3A,ADCY2,NRXN2,KCNH8,KCNS3,GRIA3,GRIK2,KCNG1,SHANK1,GRIK5,TUBA4B,NLGN4X,KCNMA1,STX1A,GRIN1,KCNH6,GNAL,KCNH7,ADCY5,GAD1,GABRG3,LRRTM4,KCNN1,GRIP2,DLG4,CACNA1A,GRIP1,GRIN2A,KCNH3,KCNK17,TUBB8,RASGRF2,SYT1,CHRNA4,GABRA5,CACNA1B,KCNMB2,SYN1,LRFN2,GLRA2,KCNK16,SLC18A2,KCNN2,CHRNA6,KCNJ11,KCND1,SLITRK3,NRXN1,TUBB2A,KCNG3,SLITRK5,DLGAP2,IL1RAPL2,KCNK4,NLGN4Y,GRM5,KCNH5,GRIA2,KCNK9,TUBB2B,PRKACG,HTR3D,ABCC8,CHRNA9,PRKAR2B,TUBAL3,GABRR1 |
|  | Biological oxidations | 0.001299 | 217 | CYP24A1,DPEP1,AOC1,PAPSS2,UGT1A6,MAOA,CYP21A2,SULT1C4,GGT1,CYP3A5,SULT1A2,GCLC,ADH1B,GGT5,ALDH3A1,CYP2F1,NAT1,UGT1A4,GLYATL2,GGT6,AOC3,ADH4,FMO1,ACSM5,DPEP2,MAT1A,CYP4A22,ADH7,CYP2C8,CYP11A1,FDXR,CYP2A13,NNMT,UGT1A3,GSTA1,NR1H4,CYP2J2,UGT2B17,UGT2B4,FMO2,UGT1A10,ADH6,UGT1A7,CYP51A1,CYP2A6,UGDH,UGT1A8,AOC2,CYP3A7,CYP2S1,UGT2A1,UGT1A5,PTGS1,ARNT2,ADH1C,GSTA3,SULT1B1,ACY3,UGT3A2,PTGIS,UGT2A2,UGT2B11,CYP11B1,CYP2W1 |
|  | Muscle contraction | 0.001615 | 205 | KCND3,FGF11,CAV3,KCNK2,FKBP1B,CACNA2D3,TNNC1,ACTG2,CACNA1S,DES,SCN7A,CACNA2D2,SCN1A,FXYD6,FXYD4,TNNT1,NPR1,MYLPF,ATP1A4,TCAP,SLN,KCNIP1,TTN,FGF14,CACNG6,MYH11,TNNT2,MYLK,ACTA2,KCNE4,ATP1A2,GUCY1A2,MYL2,TNNI3,FGF12,TPM2,GATA4,CLIC2,NPR2,SCN4A,TNNI1,TRDN,KCNK17,MYBPC1,MYBPC3,KCNK16,KCNJ11,ATP2B3,KCND1,KAT2B,MYL1,SCN3A,ANXA1,TNNT3,KCNK4,LMOD1,KCNK9,ACTC1,CACNG7,MYBPC2,ATP2B2 |
|  | Glucuronidation | 0.005269 | 25 | UGT1A6,UGT1A4,UGT1A3,UGT2B17,UGT2B4,UGT1A10,UGT1A7,UGDH,UGT1A8,UGT2A1,UGT1A5,UGT3A2,UGT2A2,UGT2B11 |
|  | TNFs bind their physiological receptors | 0.046655 | 29 | CD27,EDA,TNFRSF13B,TNFRSF18,TNFSF11,EDARADD,EDA2R,TNFRSF8,TNFSF14,LTA,CD70,TNFRSF9,TNFRSF17,TNFSF18 |
